# Integrated care for older people or people living with frailty and waiting times/lists – a mixed methods rapid review

**DOI:** 10.1101/2025.06.04.25328979

**Authors:** Judit Csontos, Elizabeth Gillen, Deborah Edwards, Adil Islam, Jacob Davies, Rhiannon Tudor Edwards, Adrian Edwards, Alison Cooper, Ruth Lewis

## Abstract

Integrated care can be defined as the joining up of different health and/or social services to deliver care that meets individuals’ needs in an efficient way. There is limited clarity about the effectiveness of integrated care interventions in improving the timeliness of health and social care delivery. To address this gap, a rapid review was conducted, incorporating both quantitative and qualitative perspectives that evaluate the impact of integrated care interventions on waiting times and waiting lists.

The review included studies published between 2015 and 2024. Sixty-one studies were identified out of which 30 reported integrated care interventions operating across two or more services. Studies were conducted in a number of different countries. Study population included older people (over the age of 65) with various injuries and diseases, and aged care or palliative care needs. The interventions involved integration across different services, with most covering both health and social care. All interventions were multifaceted.

Weak quantitative evidence from multiple studies suggests that integrated care interventions including multidisciplinary team (MDT) working, pathways/ protocols and/or care coordination as their main element may help reduce various waiting times, for example time to admission and/or time to surgery in older people with hip fracture. Strong quantitative evidence from two studies shows that a multidisciplinary assessment for older people presenting at an emergency department (ED) for various reasons, is effective in reducing time spent in the ED.

Qualitative studies mainly investigated waiting times from healthcare professionals’ perspectives. The findings suggest that integrated care interventions could support early assessment and diagnosis of dementia and complex chronic geriatric conditions; enable more timely symptom management and care planning in nursing homes; reduce processing time of aged care referrals in primary and community care; help streamline inpatient care for ageing associated diseases; and reduce delays for hip fracture care. One study explored older people’s and their relatives’ experiences and findings suggest that an ED avoidance service for older adults with urgent but non-emergency needs may help reduce emergency waiting times.

There is a need for high quality research including studies i) investigating the effect of integrated care on waiting times, ii) evaluating the effectiveness of organisational integration on waiting times, iii) exploring older people’s experiences with waiting times in relation to integrated care.

Policy and Practice Implications: There is some evidence that MDTs, integrated care pathways, and care coordination may improve inpatient waiting times to surgery, and emergency waiting times in an ED. Thus, initiatives supporting the development and implementation of these integrated care interventions is crucial.

**Funding statement:** The authors and their Institutions were funded for this work by the Health and Care Research Wales Evidence Centre, itself funded by Health and Care Research Wales on behalf of Welsh Government]

## EXECUTIVE SUMMARY

### What is a Rapid Review?

Our rapid reviews (RR) use a variation of the systematic review approach, abbreviating or omitting some components to generate the evidence to inform stakeholders promptly whilst maintaining attention to bias.

### Who is this Rapid Review for?

This Rapid Review was conducted on request from the Bevan Commission and Cardiff and Vale University Health Board. It is intended for policy makers but could also be of use for health and social care providers and third sector organisations.

### Background / Aim of Rapid Review

Integrated care can be defined as the joining up of different health and/or social services to deliver care that meets individuals’ needs in an efficient way. Increasing waiting times and an ageing population are well-recognised policy drivers for service integration, although there remains limited clarity about the effectiveness of integrated care interventions in improving the timeliness of health and social care delivery. To address this gap, a rapid review was conducted, incorporating both quantitative and qualitative perspectives that evaluate the impact of integrated care interventions on waiting times and waiting lists. Whilst the review includes a description of all relevant studies, the synthesis of the findings focuses on studies that included integrated care that operated across two or more services (primary, hospital, community or social care).

### Results of the Rapid Review

#### Recency of the evidence base

▪ The review included evidence available up until January 2025. The included studies were published between 2015 and 2024.

#### Extent of the evidence base

▪ Sixty-one studies were identified out of which 30 reported integrated care interventions operating across two or more services (23 reported quantitative data and 7 qualitative data).

▪ Quantitative study designs included: uncontrolled before and after studies (n=12), cohort studies (n=6), controlled before and after studies (n=2), randomised controlled trials (n=2) and non-randomised controlled trials (n=1). Qualitative data was gathered from qualitative descriptive studies (n=4), mixed-methods studies (n=2), and a descriptive survey with open ended questions (n=1).

▪ Studies were conducted in European countries (n=12, including 2 from the UK), USA (n=8), Canada (n=4), Australia (n=3), Japan (n=1), and across multiple countries (n=2).

▪ Study population included older people (over the age of 65) with hip or other fractures (n=15), non-surgical traumatic injuries (n=2), various emergency (n=3) or urgent care needs (n=2), mental health conditions (n=2), dementia (n=2), complex chronic geriatric diseases (n=1), ageing associated diseases and aged care needs (n=2), or palliative care needs (n=1).

▪ The interventions involved integration across two (n=16), three (n=9) or four (n=5) different services, with most covering both health and social care (n=25), although the mechanism of integration varied. All interventions were multifaceted with the most consistently reported elements being multidisciplinary team (MDT) working, development of pathways and protocols, and care coordination.

▪ Waiting times and waiting lists were categorised as inpatient, emergency, and routine care. Inpatient waiting times, such as time to surgery, were the most commonly reported (n=18).

#### Key findings and certainty of the evidence

▪ Weak quantitative evidence from multiple studies suggests that **integrated care interventions** including MDT, pathways/protocols and/ or care coordination as their main element **may help reduce** the following **waiting times**: time to admission and time to surgery for hip or other fractures; time to first goals-of-care assessment for non-surgical traumatic injuries; time until geriatric care review for older people presenting at the emergency department (ED); primary care wait time for older people with urgent needs; time to treatment initiation and time to appointment for older people with mental health conditions; and time to investigation of older people’s palliative care needs and desires (GP self-report). The evidence was rated weak, due to weak study designs, low study quality, and inconsistencies in the findings.

▪ Strong quantitative evidence from two studies shows that a **multidisciplinary assessment** for older people presenting at ED for various reasons, is effective in **reducing time spent in the ED**.

▪ Qualitative studies mainly investigated waiting times from healthcare professionals’ perspectives. The findings suggest that integrated care interventions could support early assessment and diagnosis of dementia and complex chronic geriatric conditions; enable more timely symptom management and care planning in nursing homes; reduce processing time of aged care referrals in primary and community care; help streamline inpatient care for ageing associated diseases; and reduce delays for hip fracture care.

▪ One qualitative study explored older people’s and their relatives’ experiences regarding an integrated ED avoidance service. The findings suggest that the ED avoidance service for older adults with urgent but non-emergency needs may help reduce emergency waiting times.

### Research Implications and Evidence Gaps

▪ There is a need for high quality studies investigating the effect of integrated care on waiting times, particularly on routine care and elective waiting times.

▪ The majority of the identified studies focused on MDTs, integrated pathways/protocols and/or care coordination and their impact on waiting times. There seems to be less focus on organisational integration, such as coordination of governance across providers or joint commissioning. More research with rigorous study designs is necessary to evaluate the effectiveness of organisational integration on waiting times.

▪ There is a need for high quality qualitative research that explores people’s experiences with waiting times in relation to integrated care, particularly from older and frail people’s perspectives.

### Policy and Practice Implications

▪ There is some evidence that MDTs, integrated care pathways, and care coordination may improve inpatient waiting times to surgery, and emergency waiting times in an ED. Thus, initiatives supporting the development and implementation of these integrated care interventions is crucial.

### Economic considerations

▪ Hospital costs increase with length of inpatient waiting time, suggesting initiatives reducing time spent waiting may bring positive economic benefit to the NHS.

▪ An estimated £73 billion in total benefits may be generated between 2023 and 2027 if the NHS meets its waiting list reduction targets.

The certainty of evidence from quantitative studies has been assessed using the Critical Appraisal Tool (CAT) based on the guidance by Public Health Agency of Canada (2014).

## 1. BACKGROUND

### 1.1 Who is this review for?

This Rapid Review was conducted as part of the Health and Care Research Wales Evidence Centre Work Programme. The review question was proposed by the Bevan Commission and Cardiff and Vale University Health Board.

### 1.2 Background and purpose of this review

Integrated care can be defined as the joining up of different health and/or social services to deliver care that meets individuals’ needs in an efficient way (Scobie 2021). In Wales, The Well-being of Future Generations Act (2015) provides a framework for public bodies to work together on preventative and integrated approaches, while the Healthier Wales long-term plan aims to organise integrated care around individuals and communities (Welsh Government 2021). Integrated care has been a long standing policy aim of governments across the UK (Reed et al. 2021), as the fragmented health and social care systems can make it difficult for people to receive timely care (Bevan Commission 2024).

Waiting times in the National Health Service (NHS) have significantly increased over the past decade (Welsh Government 2024b). In Wales, referral to treatment wait lists included 769,000 open care pathways in March 2024 compared to 383,000 in January 2013 (Welsh Government 2024b). Median waiting times to treatment were also twice as long, with people waiting approximately 21.8 weeks in March 2024 (Welsh Government 2024b, StatsWales 2024). Emergency department waiting times have also increased, reaching a peak in March 2022 at three hours and eight minutes (Welsh Government 2024c). These increased waiting times can disproportionately affect older people, who are more likely to live with a health condition (Welsh Parliament 2022, Fisher & Taylor 2024).

Heath and social care services across the UK face growing challenges regarding an aging population. In the UK, approximately 19% of the population was aged 65 or above in 2022 in comparison with 13% in 1972 (Barton et al. 2024). However, aging particularly affects Wales, where it is estimated that 30% of the population will be aged 60 or over by 2026 (Older People’s Commissioner for Wales 2023). Additionally, people over the age of 65 are more likely to become frail, with over a quarter of those aged 85 or more expected to be affected (Turner 2014). Frailty can be defined as a long-term condition whereby the resilience of body systems gradually declines, meaning that those affected are less likely to cope with minor illness, infection, or stress (Welsh Government 2024a). While the likelihood of frailty rises with age, it can also affect younger people, as risks include sociodemographic, clinical, lifestyle, and biological factors (Bai et al. 2023).

While increasing waiting times and an ageing population are well-recognised policy drivers for service integration, there remains limited clarity about the effectiveness of integrated care interventions in improving the timeliness of health and social care delivery (Baxter et al. 2018a; Baxter et al. 2018b). To address this gap, a rapid review was conducted, incorporating both quantitative and qualitative perspectives that evaluate the impact of integrated care interventions on waiting times and waiting lists. Therefore, the aim of this review was twofold: (i) to assess the effectiveness of integrated care in reducing waiting times and/or lists for older people or individuals living with frailty; and (ii) to explore the views of healthcare professionals and older people or individuals living with frailty regarding waiting times in the context of integrated care.

## 2. RESULTS

Following a thorough search of bibliographic databases and the grey literature, 61 studies examining various forms of integrated care met the initial inclusion criteria. Studies were eligible for inclusion if they focused on older people or people living with frailty, investigated integrated care interventions and reported waiting time/list outcomes or experiences. Waiting time outcomes were conceptualised as any period where patients were waiting for an appointment, diagnosis or treatment, whether this was in the emergency department (ED), inpatient or outpatient (routine) setting. Waiting lists could include number of people on a waiting list or the number of people waiting more than a defined period. Including studies investigating integrated care interventions that did not report waiting time or waiting list outcomes were out of the scope of this review. The methods and detailed eligibility criteria used to conduct this review are presented in Section 5.1 and the study selection process is detailed in Section 6.1.

Table 1 provides an overview of the 61 included studies. This table summarises the populations, countries of origin, the service level integration (which could include primary care (general practice), hospital care (secondary and tertiary), community care, or social care), study designs, and waiting time outcomes.

**Table 1:**
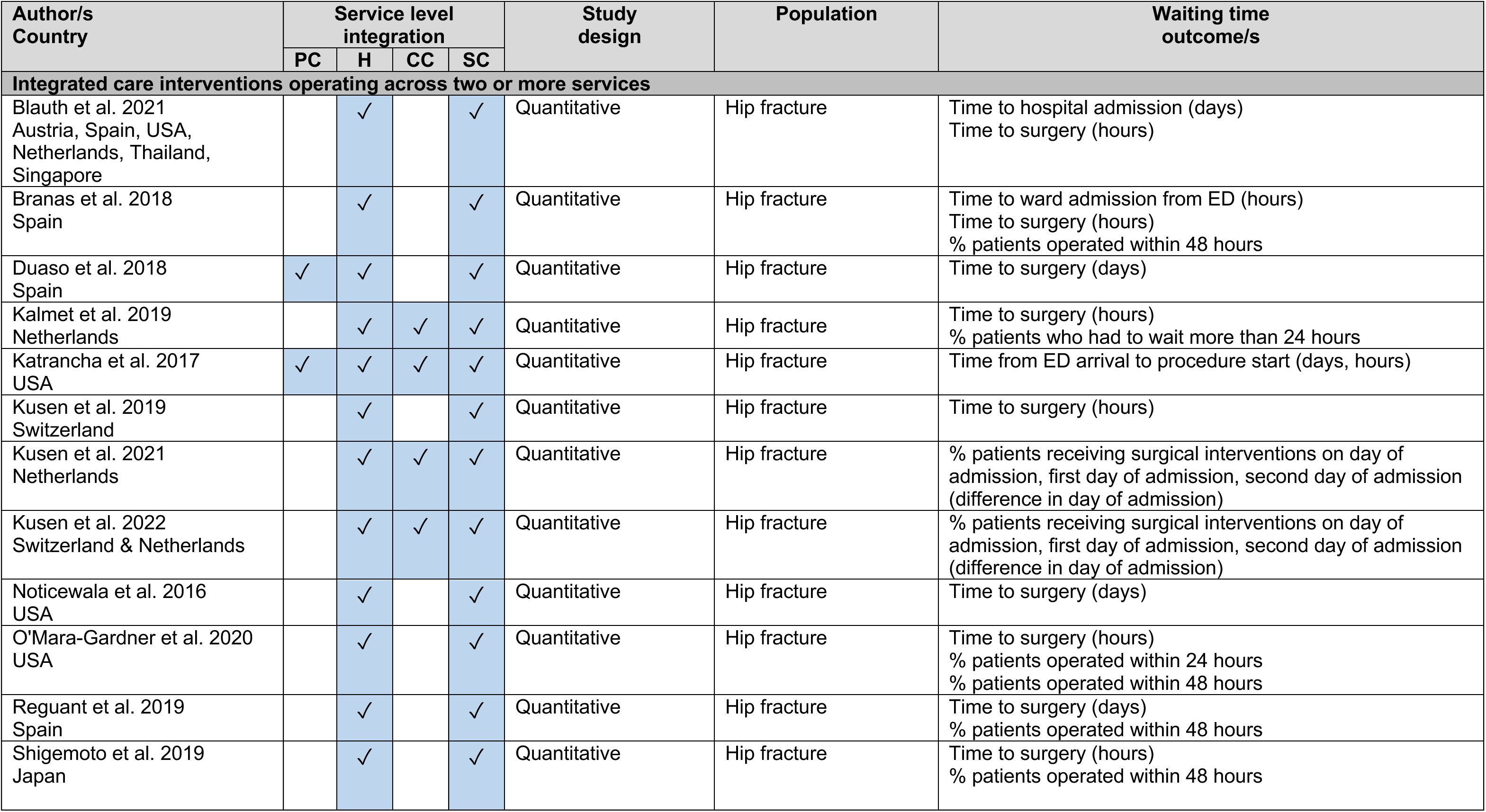
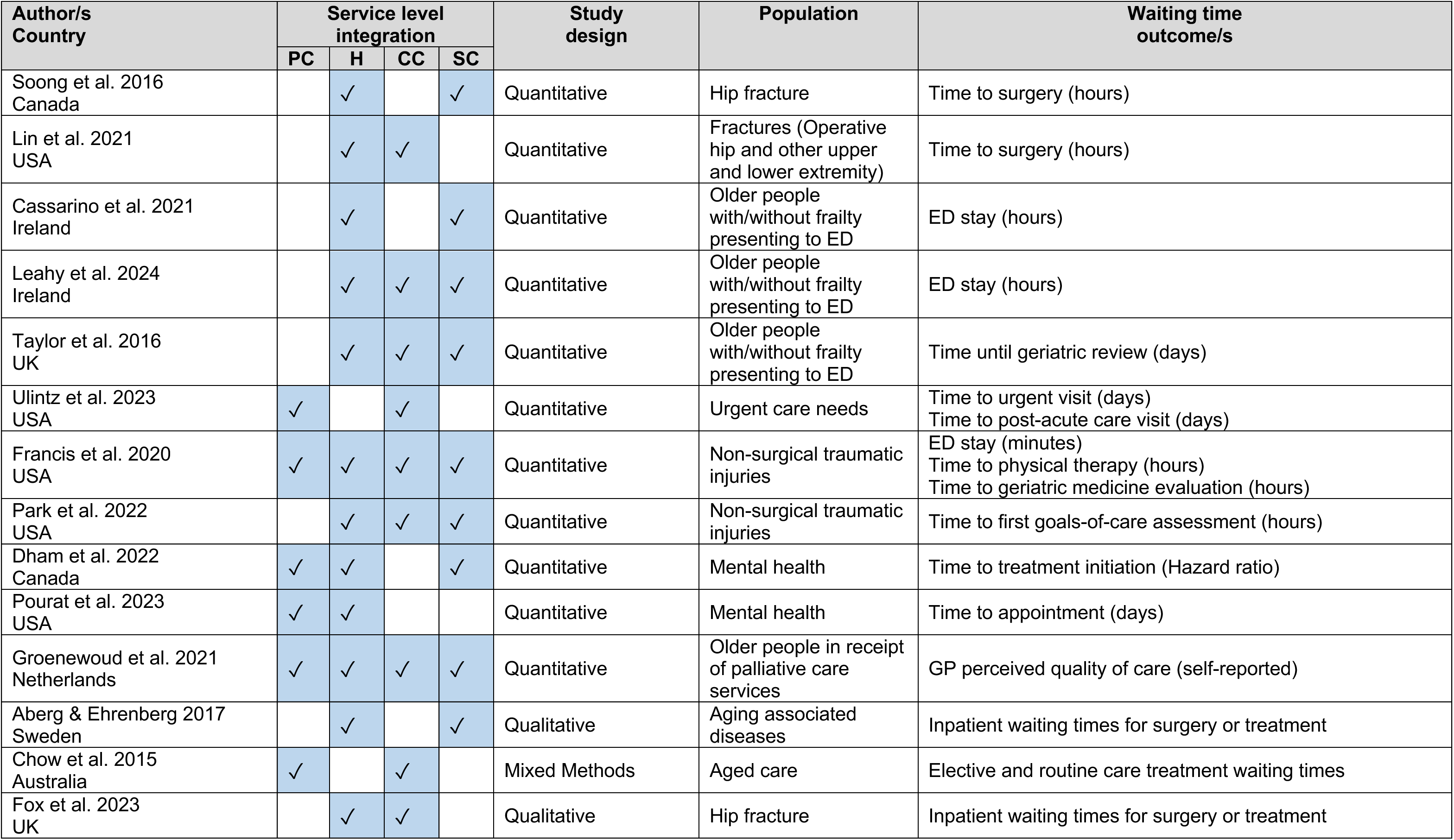
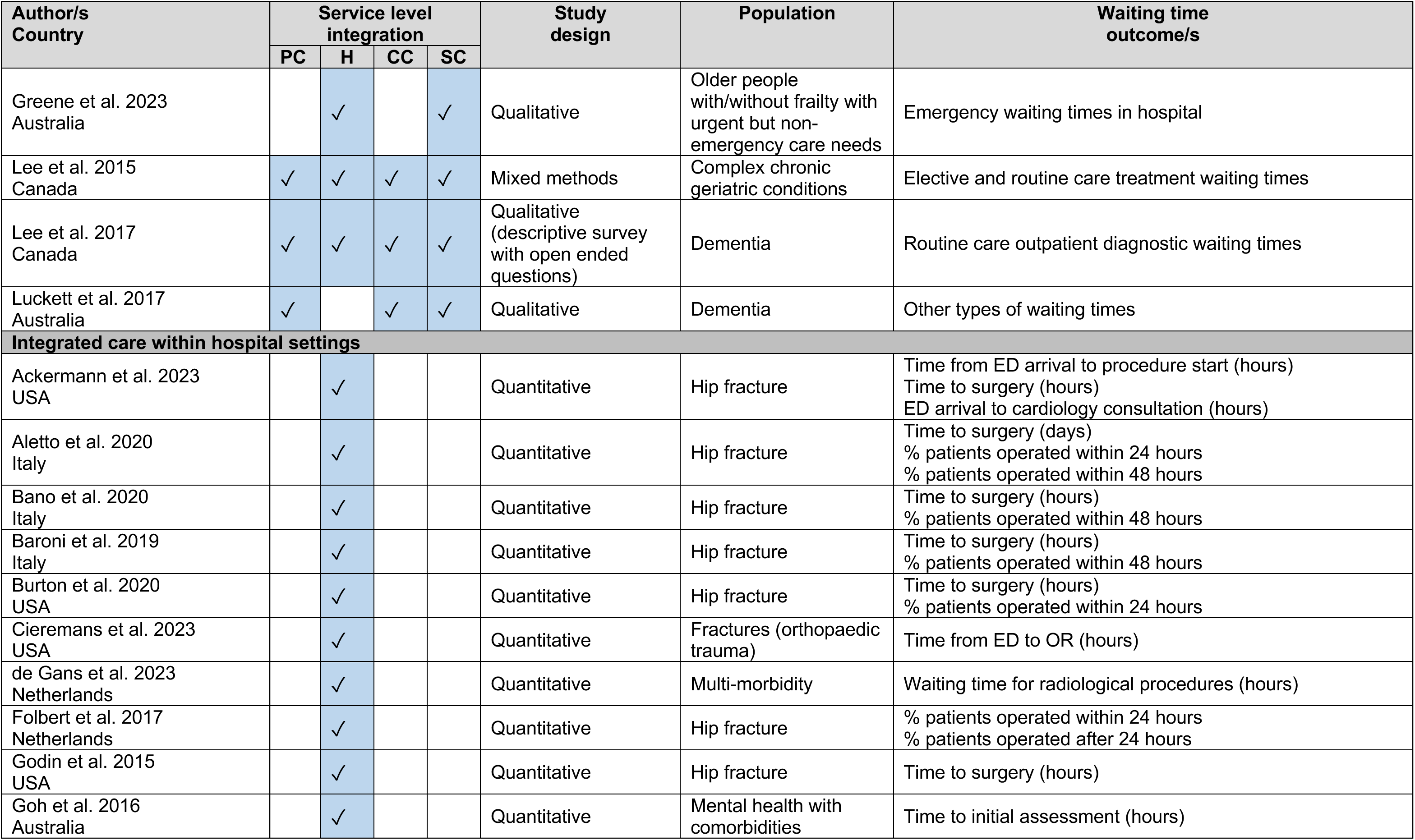
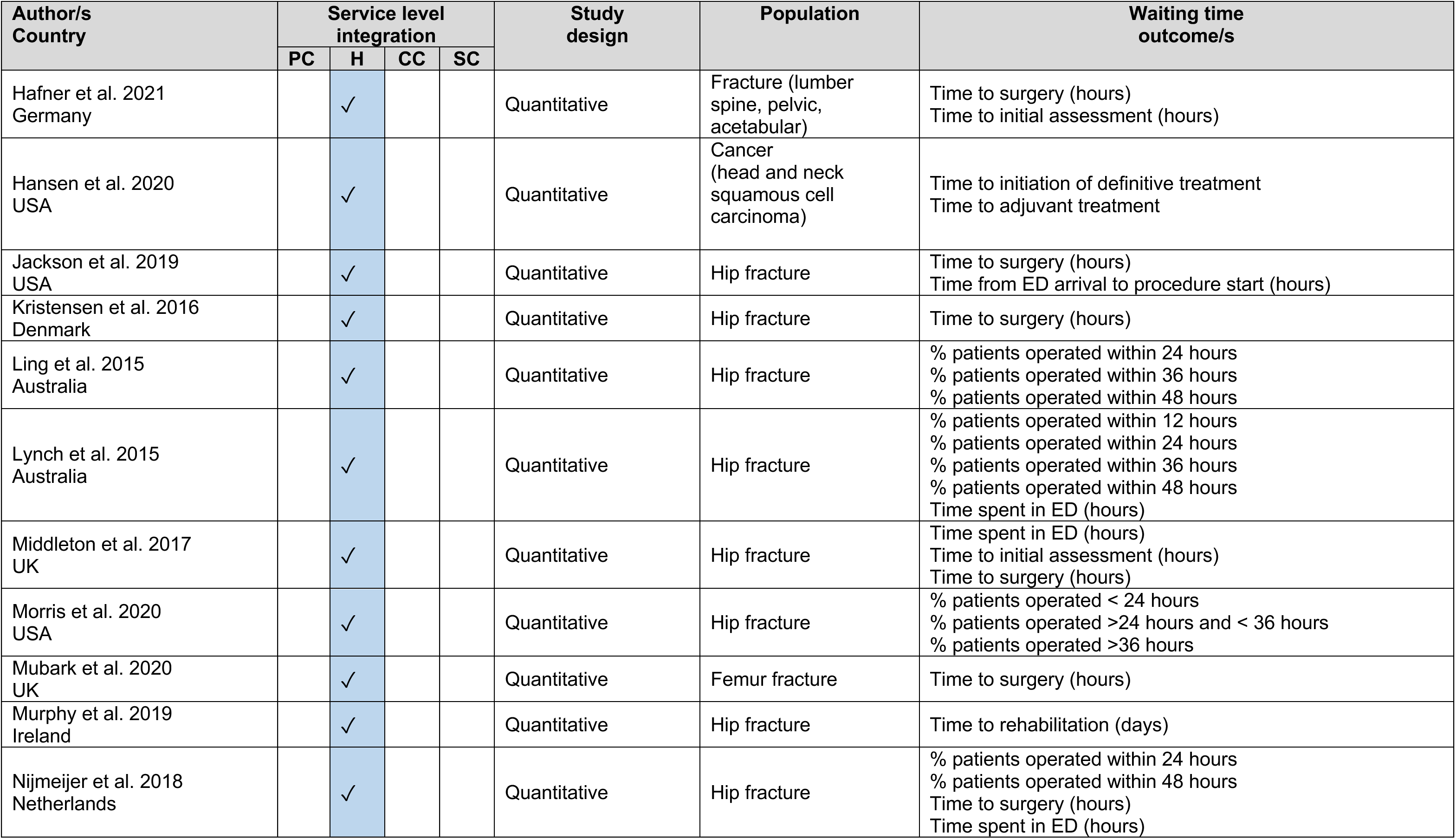
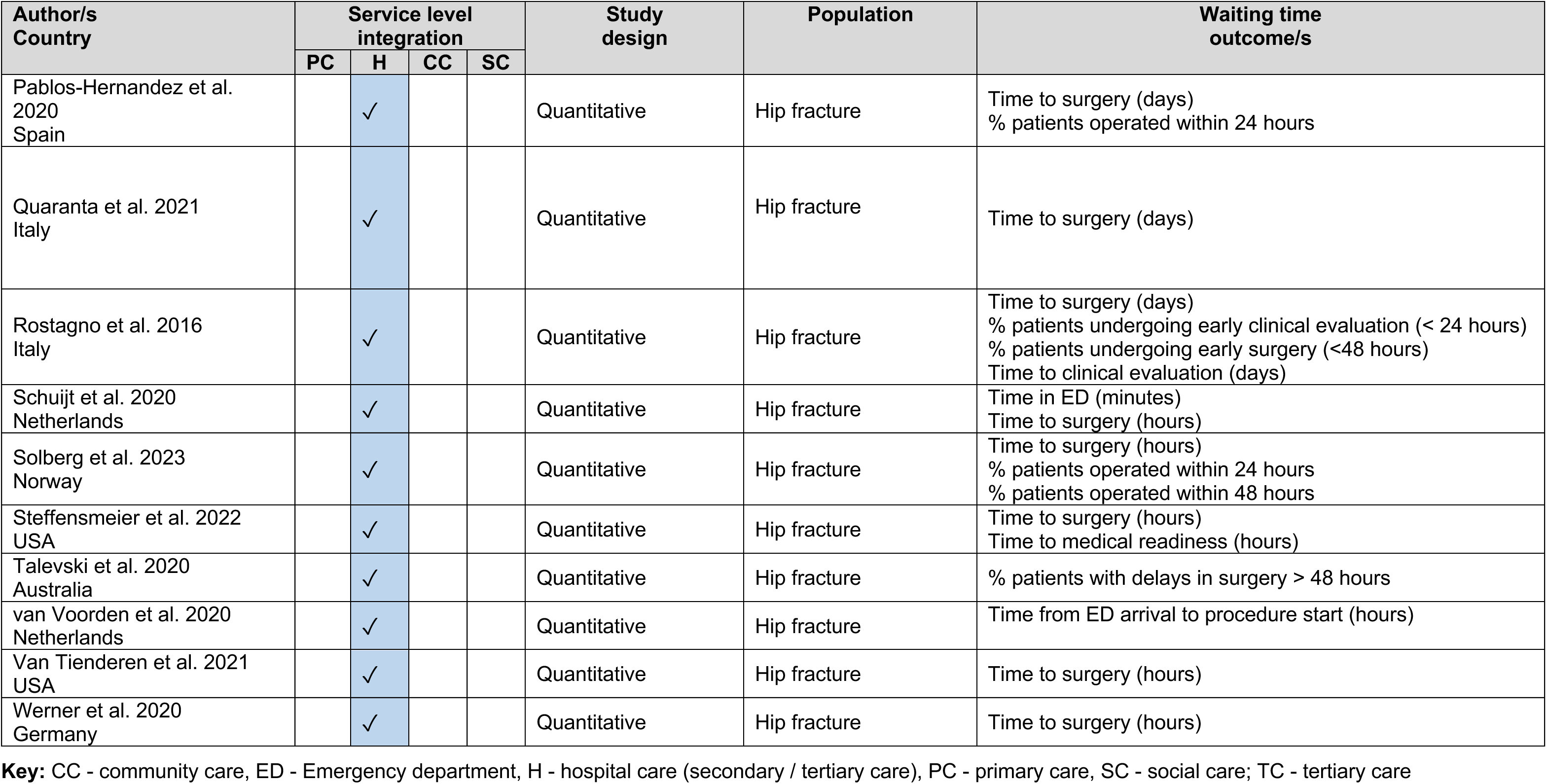
Summary table of included studies.

| Author/s<br>Country | Service level<br>integration |  |  |  | Study<br>design | Population | Waiting time<br>outcome/s |
| --- | --- | --- | --- | --- | --- | --- | --- |
|  | PC | H | CC | SC |  |  |  |
| Integrated care interventions operating across two or more services |  |  |  |  |  |  |  |
| Blauth et al. 2021<br>Austria, Spain, USA,<br>Netherlands, Thailand,<br>Singapore |  | ✓ |  | ✓ | Quantitative | Hip fracture | Time to hospital admission (days)<br>Time to surgery (hours) |
| Branas et al. 2018<br>Spain |  | ✓ |  | ✓ | Quantitative | Hip fracture | Time to ward admission from ED (hours)<br>Time to surgery (hours)<br>% patients operated within 48 hours |
| Duaso et al. 2018<br>Spain | ✓ | ✓ |  | ✓ | Quantitative | Hip fracture | Time to surgery (days) |
| Kalmet et al. 2019<br>Netherlands |  | ✓ | ✓ | ✓ | Quantitative | Hip fracture | Time to surgery (hours)<br>% patients who had to wait more than 24 hours |
| Katrancha et al. 2017<br>USA | ✓ | ✓ | ✓ | ✓ | Quantitative | Hip fracture | Time from ED arrival to procedure start (days, hours) |
| Kusen et al. 2019<br>Switzerland |  | ✓ |  | ✓ | Quantitative | Hip fracture | Time to surgery (hours) |
| Kusen et al. 2021<br>Netherlands |  | ✓ | ✓ | ✓ | Quantitative | Hip fracture | % patients receiving surgical interventions on day of admission, first day of admission, second day of admission (difference in day of admission) |
| Kusen et al. 2022<br>Switzerland & Netherlands |  | ✓ | ✓ | ✓ | Quantitative | Hip fracture | % patients receiving surgical interventions on day of admission, first day of admission, second day of admission (difference in day of admission) |
| Noticewala et al. 2016<br>USA |  | ✓ |  | ✓ | Quantitative | Hip fracture | Time to surgery (days) |
| O'Mara-Gardner et al. 2020<br>USA |  | ✓ |  | ✓ | Quantitative | Hip fracture | Time to surgery (hours)<br>% patients operated within 24 hours<br>% patients operated within 48 hours |
| Reguant et al. 2019<br>Spain |  | ✓ |  | ✓ | Quantitative | Hip fracture | Time to surgery (days)<br>% patients operated within 48 hours |
| Shigemoto et al. 2019<br>Japan |  | ✓ |  | ✓ | Quantitative | Hip fracture | Time to surgery (hours)<br>% patients operated within 48 hours |
|  | PC | H | CC | SC |  |  |  |
| Soong et al. 2016<br>Canada |  | ✓ |  | ✓ | Quantitative | Hip fracture | Time to surgery (hours) |
| Lin et al. 2021<br>USA |  | ✓ | ✓ |  | Quantitative | Fractures (Operative hip and other upper and lower extremity) | Time to surgery (hours) |
| Cassarino et al. 2021<br>Ireland |  | ✓ |  | ✓ | Quantitative | Older people with/without frailty presenting to ED | ED stay (hours) |
| Leahy et al. 2024<br>Ireland |  | ✓ | ✓ | ✓ | Quantitative | Older people with/without frailty presenting to ED | ED stay (hours) |
| Taylor et al. 2016<br>UK |  | ✓ | ✓ | ✓ | Quantitative | Older people with/without frailty presenting to ED | Time until geriatric review (days) |
| Ulintz et al. 2023<br>USA | ✓ |  | ✓ |  | Quantitative | Urgent care needs | Time to urgent visit (days)<br>Time to post-acute care visit (days) |
| Francis et al. 2020<br>USA | ✓ | ✓ | ✓ | ✓ | Quantitative | Non-surgical traumatic injuries | ED stay (minutes)<br>Time to physical therapy (hours)<br>Time to geriatric medicine evaluation (hours) |
| Park et al. 2022<br>USA |  | ✓ | ✓ | ✓ | Quantitative | Non-surgical traumatic injuries | Time to first goals-of-care assessment (hours) |
| Dham et al. 2022<br>Canada | ✓ | ✓ |  | ✓ | Quantitative | Mental health | Time to treatment initiation (Hazard ratio) |
| Pourat et al. 2023<br>USA | ✓ | ✓ |  |  | Quantitative | Mental health | Time to appointment (days) |
| Groenewoud et al. 2021<br>Netherlands | ✓ | ✓ | ✓ | ✓ | Quantitative | Older people in receipt of palliative care services | GP perceived quality of care (self-reported) |
| Aberg & Ehrenberg 2017<br>Sweden |  | ✓ |  | ✓ | Qualitative | Aging associated diseases | Inpatient waiting times for surgery or treatment |
| Chow et al. 2015<br>Australia | ✓ |  | ✓ |  | Mixed Methods | Aged care | Elective and routine care treatment waiting times |
| Fox et al. 2023<br>UK |  | ✓ | ✓ |  | Qualitative | Hip fracture | Inpatient waiting times for surgery or treatment |
|  | PC | H | CC | SC |  |  |  |
| Greene et al. 2023<br>Australia |  | ✓ |  | ✓ | Qualitative | Older people<br>with/without frailty with<br>urgent but non-emergency care needs | Emergency waiting times in hospital |
| Lee et al. 2015<br>Canada | ✓ | ✓ | ✓ | ✓ | Mixed methods | Complex chronic<br>geriatric conditions | Elective and routine care treatment waiting times |
| Lee et al. 2017<br>Canada | ✓ | ✓ | ✓ | ✓ | Qualitative<br>(descriptive survey<br>with open ended<br>questions) | Dementia | Routine care outpatient diagnostic waiting times |
| Luckett et al. 2017<br>Australia | ✓ |  | ✓ | ✓ | Qualitative | Dementia | Other types of waiting times |
| **Integrated care within hospital settings** |  |  |  |  |  |  |  |
| Ackermann et al. 2023<br>USA |  | ✓ |  |  | Quantitative | Hip fracture | Time from ED arrival to procedure start (hours)<br>Time to surgery (hours)<br>ED arrival to cardiology consultation (hours) |
| Aletto et al. 2020<br>Italy |  | ✓ |  |  | Quantitative | Hip fracture | Time to surgery (days)<br>% patients operated within 24 hours<br>% patients operated within 48 hours |
| Bano et al. 2020<br>Italy |  | ✓ |  |  | Quantitative | Hip fracture | Time to surgery (hours)<br>% patients operated within 48 hours |
| Baroni et al. 2019<br>Italy |  | ✓ |  |  | Quantitative | Hip fracture | Time to surgery (hours)<br>% patients operated within 48 hours |
| Burton et al. 2020<br>USA |  | ✓ |  |  | Quantitative | Hip fracture | Time to surgery (hours)<br>% patients operated within 24 hours |
| Cieremans et al. 2023<br>USA |  | ✓ |  |  | Quantitative | Fractures (orthopaedic<br>trauma) | Time from ED to OR (hours) |
| de Gans et al. 2023<br>Netherlands |  | ✓ |  |  | Quantitative | Multi-morbidity | Waiting time for radiological procedures (hours) |
| Folbert et al. 2017<br>Netherlands |  | ✓ |  |  | Quantitative | Hip fracture | % patients operated within 24 hours<br>% patients operated after 24 hours |
| Godin et al. 2015<br>USA |  | ✓ |  |  | Quantitative | Hip fracture | Time to surgery (hours) |
| Goh et al. 2016<br>Australia |  | ✓ |  |  | Quantitative | Mental health with<br>comorbidities | Time to initial assessment (hours) |
|  | PC | H | CC | SC |  |  |  |
| Hafner et al. 2021<br>Germany |  | ✓ |  |  | Quantitative | Fracture (lumber spine, pelvic, acetabular) | Time to surgery (hours)<br>Time to initial assessment (hours) |
| Hansen et al. 2020<br>USA |  | ✓ |  |  | Quantitative | Cancer (head and neck squamous cell carcinoma) | Time to initiation of definitive treatment<br>Time to adjuvant treatment |
| Jackson et al. 2019<br>USA |  | ✓ |  |  | Quantitative | Hip fracture | Time to surgery (hours)<br>Time from ED arrival to procedure start (hours) |
| Kristensen et al. 2016<br>Denmark |  | ✓ |  |  | Quantitative | Hip fracture | Time to surgery (hours) |
| Ling et al. 2015<br>Australia |  | ✓ |  |  | Quantitative | Hip fracture | % patients operated within 24 hours<br>% patients operated within 36 hours<br>% patients operated within 48 hours |
| Lynch et al. 2015<br>Australia |  | ✓ |  |  | Quantitative | Hip fracture | % patients operated within 12 hours<br>% patients operated within 24 hours<br>% patients operated within 36 hours<br>% patients operated within 48 hours<br>Time spent in ED (hours) |
| Middleton et al. 2017<br>UK |  | ✓ |  |  | Quantitative | Hip fracture | Time spent in ED (hours)<br>Time to initial assessment (hours)<br>Time to surgery (hours) |
| Morris et al. 2020<br>USA |  | ✓ |  |  | Quantitative | Hip fracture | % patients operated < 24 hours<br>% patients operated >24 hours and < 36 hours<br>% patients operated >36 hours |
| Mubark et al. 2020<br>UK |  | ✓ |  |  | Quantitative | Femur fracture | Time to surgery (hours) |
| Murphy et al. 2019<br>Ireland |  | ✓ |  |  | Quantitative | Hip fracture | Time to rehabilitation (days) |
| Nijmeijer et al. 2018<br>Netherlands |  | ✓ |  |  | Quantitative | Hip fracture | % patients operated within 24 hours<br>% patients operated within 48 hours<br>Time to surgery (hours)<br>Time spent in ED (hours) |
|  | PC | H | CC | SC |  |  |  |
| Pablos-Hernandez et al. 2020<br>Spain |  | ✓ |  |  | Quantitative | Hip fracture | Time to surgery (days)<br>% patients operated within 24 hours |
| Quaranta et al. 2021<br>Italy |  | ✓ |  |  | Quantitative | Hip fracture | Time to surgery (days) |
| Rostagno et al. 2016<br>Italy |  | ✓ |  |  | Quantitative | Hip fracture | Time to surgery (days)<br>% patients undergoing early clinical evaluation (< 24 hours)<br>% patients undergoing early surgery (<48 hours)<br>Time to clinical evaluation (days) |
| Schuijt et al. 2020<br>Netherlands |  | ✓ |  |  | Quantitative | Hip fracture | Time in ED (minutes)<br>Time to surgery (hours) |
| Solberg et al. 2023<br>Norway |  | ✓ |  |  | Quantitative | Hip fracture | Time to surgery (hours)<br>% patients operated within 24 hours<br>% patients operated within 48 hours |
| Steffensmeier et al. 2022<br>USA |  | ✓ |  |  | Quantitative | Hip fracture | Time to surgery (hours)<br>Time to medical readiness (hours) |
| Talevski et al. 2020<br>Australia |  | ✓ |  |  | Quantitative | Hip fracture | % patients with delays in surgery > 48 hours |
| van Voorden et al. 2020<br>Netherlands |  | ✓ |  |  | Quantitative | Hip fracture | Time from ED arrival to procedure start (hours) |
| Van Tienderen et al. 2021<br>USA |  | ✓ |  |  | Quantitative | Hip fracture | Time to surgery (hours) |
| Werner et al. 2020<br>Germany |  | ✓ |  |  | Quantitative | Hip fracture | Time to surgery (hours) |

**Key:** CC - community care, ED - Emergency department, H - hospital care (secondary / tertiary care), PC - primary care, SC - social care; TC - tertiary care

All the included studies focused on older people with or without frailty, and no studies were identified where the population focused on people living with frailty under the age of 65. The majority of the studies focused on older people who had hip (39 studies) or other fractures (four studies) needing surgery. Other populations included older people presenting to emergency departments (ED) for various reasons (three studies), older people with multimorbidity and/or urgent care needs (three studies), and older people with mental health conditions (with or without comorbidities) (three studies). Fewer studies focused on older people experiencing non-surgical traumatic injuries (from falls, motor vehicle collision or other causes, but injury not specified) (two studies), dementia (two studies), aged care needs or aging associated diseases (two studies), chronic diseases (one study), cancer (one study) and palliative care needs (one study).

There were 30 studies that investigated integrated care interventions operating across two or more services, whilst 31 studies focused on integrated care within hospital settings (secondary / tertiary care). Given the large number of studies and the timeframe of the review, this rapid review focuses on the 30 studies with integrated care interventions operating across two or more services. These studies were further split into two groups based on whether they reported quantitative data on the effectiveness of integrated care in reducing waiting times or qualitative data on people’s waiting time experiences in the context of integrated.

The evidence related to the effectiveness of integrated care interventions in reducing waiting times is reported in Section 2.1 and 2.2, whilst findings based on people’s experiences of waiting times (healthcare professionals, older people and their relatives) in the context of integrated care is presented in Section 2.3 and 2.4. A brief summary of the 31 studies that focused on integrated care within hospital settings is provided in Section 2.5.

### 2.1 Quantitative review of the effectiveness of integrated care

This section addresses the review aim focusing on the effectiveness of integrated care in reducing waiting times for older people. The section starts with an overview of the identified quantitative studies, summarising study designs, country of origin, and the population focus. Then the quality of the studies is presented, followed by the characteristics of the integrated care interventions. The waiting times outcomes covered by the studies is described and finally the results of each study and the effectiveness of the integrated care interventions is reported.

#### 2.1.1 Overview of the quantitative evidence base

From the 30 studies in which the integrated care interventions operated across two or more services, 23 had a quantitative study design. Two RCTs (Cassarino et al. 2021, Leahy et al. 2024), one non-randomised controlled trial (Dham et al. 2022), two controlled before and after studies (Groenewoud et al. 2021, Soong et al. 2016), six cohort studies (three prospective and three retrospective) (Blauth et al. 2021, Kusen et al. 2021, Kusen et al. 2022, Lin et al. 2021, Noticewala et al. 2016, Ulintz et al. 2023), and 12 uncontrolled before and after studies were identified.

Twenty-one studies were conducted within a single country and these included:

- USA (eight studies) (Francis et al. 2020, Katrancha et al. 2017, Lin et al. 2021, Noticewala et al. 2016, O’Mara-Gardner et al. 2020, Park et al. 2022, Pourat et al. 2023, Ulintz et al. 2023)
- Spain (three studies) (Branas et al. 2018, Duaso et al. 2018, Reguant et al. 2019)
- Netherlands (three studies) (Groenewoud et al. 2021, Kalmet et al. 2019, Kusen et al. 2021)
- Canada (two studies) (Dham et al. 2022, Soong et al. 2016)
- Ireland (two studies) (Cassarino et al. 2021, Leahy et al. 2024)
- Switzerland (one study) (Kusen et al. 2019)
- Japan (one study) (Shigemoto et al. 2019)
- UK (one study) (Taylor et al. 2016)

Two further studies were conducted across multiple countries. Blauth et al. (2021) included participants from Austria, Spain, the USA, the Netherlands, Singapore, and Thailand. Similarly, the study by Kusen et al. (2022) as conducted across the Netherlands and Switzerland.

The included studies explored a range of populations with various health conditions, with the majority focusing on hip (13 studies) or other upper and lower extremity fractures needing surgery (Lin et al. 2021). Other populations included older people experiencing non-surgical traumatic injuries (from falls, motor vehicle collision or other causes, but injury not specified) (two studies) (Francis et al. 2020, Park et al. 2022), older people presenting at ED for various reasons (three studies) (Cassarino et al. 2021, Leahy et al. 2024, Taylor et al. 2016) or experiencing urgent care needs (Ulintz et al. 2023), older people with mental health conditions (two studies) (Dham et al. 2022, Pourat et al. 2023) and older people with palliative care needs (one study) (Groenewoud et al. 2021). The detailed characteristics of each included quantitative study can be found in Section 6.2.

#### 2.1.2 Quality of the quantitative studies

The quantitative evidence was critically appraised using the Analytic Study Critical Appraisal Tool (CAT) (see section 5.6 for further details) (Public Health Agency of Canada 2014), and variable quality of evidence was detected. All studies had a focused research question, and participants were representative of the target population with some concerns in three studies regarding participant selection (Leahy et al. 2024, Noticewala et al. 2016, Soong et al. 2016).

Most studies (21 studies) did not select or allocate participants randomly which led to differences between intervention and comparison groups. Many studies showed moderate (10 studies) or weak (nine studies) comparability between control and intervention groups at baseline. These differences could potentially affect the study outcomes. Follow-up was usually (20 studies) completed in the outcome of interest (waiting times), meaning participant had no missing data.

Statistical tests were mostly appropriate for the level of data and hypothesis being tested (22 studies), although most of the studies did not control for differences in baseline data or confounding factors (19 studies). Whilst some confounding variables were identified and managed (4 studies), there remained other factors that were not adequately addressed or for many not discussed or accounted for (19 studies). These unaddressed variables could significantly impact the validity of the results. Fifteen studies either did not calculate statistical power or reported insufficient power to accurately detect statistically significant differences.

When assessing the overall quality of the quantitative studies, a significant variation was observed. Only two studies (Cassarino et al. 2021, Leahy et al. 2024) were rated as high quality, indicating rigorous methodology and more reliable results, whilst nearly half of quantitative studies were assessed as medium quality (11 studies) demonstrating some limitations that could affect validity. Ten studies were rated as low quality showing significant methodological weaknesses and potential biases that could impact the reliability of the results. Detailed results from the critical appraisal can be found in section 6.3.

#### 2.1.3 Characteristics of integrated care interventions

Included studies were initially grouped according to the target population and medical specialty they focused on, namely orthogeriatric care for hip and other upper and lower extremity fractures needing surgery, older people experiencing non-surgical traumatic injury (from falls, motor vehicle collision or other causes, but injury not specified), older people presenting at ED for various reasons, older people with urgent care needs accessing primary care (GPs), older people experiencing mental health conditions, and palliative care services.

The integrated care interventions comprised of a combination of elements, such as multidisciplinary teams (MDTs), pathways or protocols, care coordination, new unit or co-location, joint patient review or discharge, integrated patient records, agreed referral criteria, joint assessment, comprehensive geriatric assessment (CGA), and professional role change. **Multidisciplinary team** refers to a group of healthcare professionals from different disciplines (for example medicine, nursing, physiotherapy, and others) working together to provide specific services for patients (Baxter et al. 2018b). **Pathways or protocols** are outlines of care provision that have set timeframes for anticipated procedures and role allocation for different healthcare professionals to ensure patients move progressively through the health and/or social care system (Baxter et al. 2018b). **Care coordination** describes the process of care organisation that is usually performed by a named point of contact who aims to bring together different healthcare providers and specialists to support the patient (Skills for Care 2018, Baxter et al. 2018b). Care coordination may also include the assessment and regular monitoring of the patient and care delivery (Skills for Care 2018, Baxter et al. 2018b). **New unit or co-location** refers to the development of new health and social care departments that brings healthcare professionals from different specialism or discipline under one roof (Baxter et aal. 2018b). **Joint patient review or discharge** describes the process of patient evaluation and discharge planning that is performed by two or more healthcare professionals and can reduce duplication, resulting in more efficient care provision (Baxter et al. 2018b). **Integrated patient records** refer to the process of documentation that allows different health and social care providers to share patient information efficiently usually by providing access to the same system (Baxter et al. 2018b). **Agreed referral criteria** outline the predetermined conditions under which referral or transfer from one service to another is initiated (Baxter et al. 2018b). **Joint assessment** can be defined as patient examination that is performed by two or more healthcare professionals from different specialisms or disciplines that enables efficient identification of patients’ needs (Baxter et al. 2018b). **Comprehensive geriatric assessment** can be defined as a multidimensional holistic assessment that considers older people’s concerns with the aim to develop a plan that can help meet their needs (British Geriatric Society 2019). The elements of each integrated care intervention are summarised in Table 2. A detailed description of the integrated care interventions is provided below.

**Table 2:**
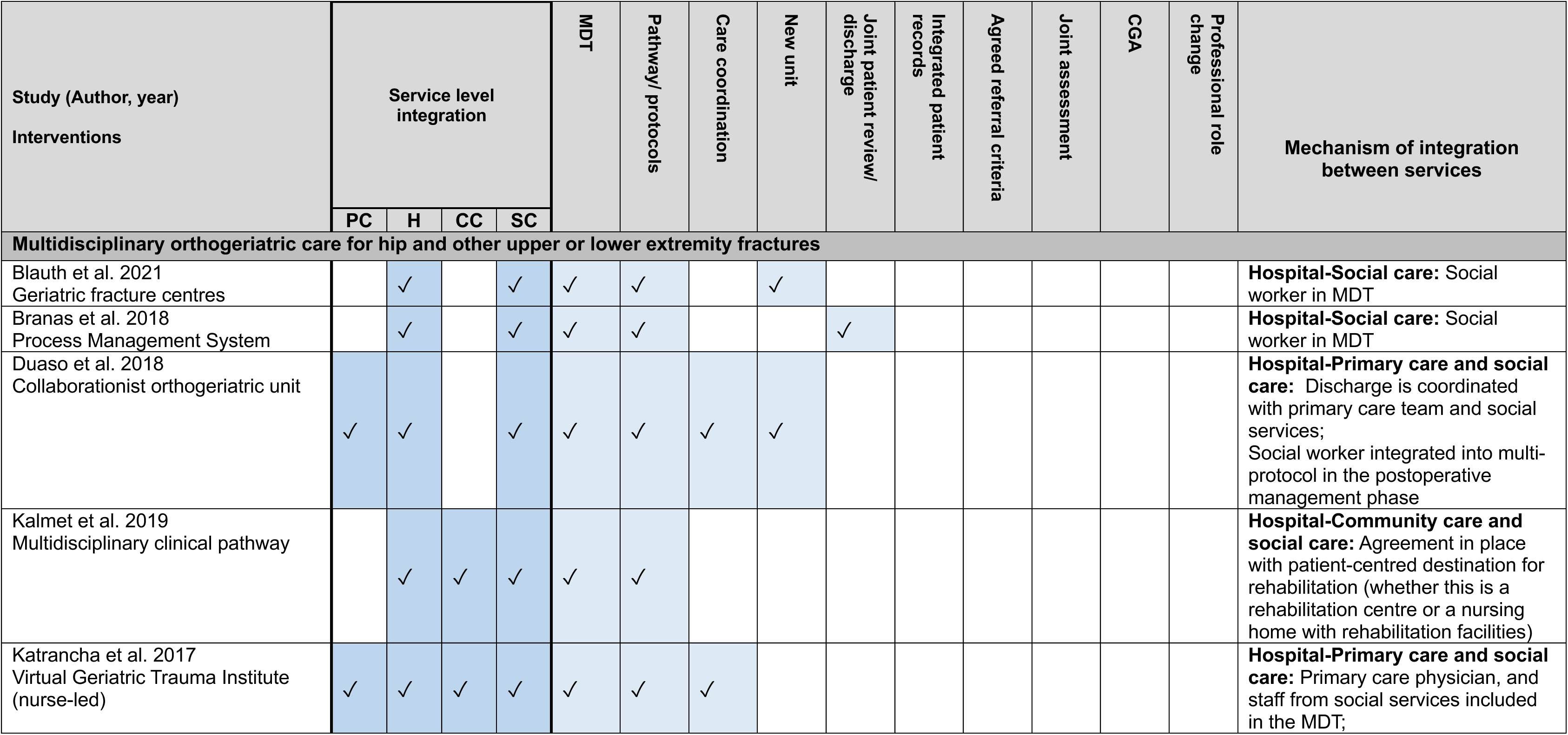
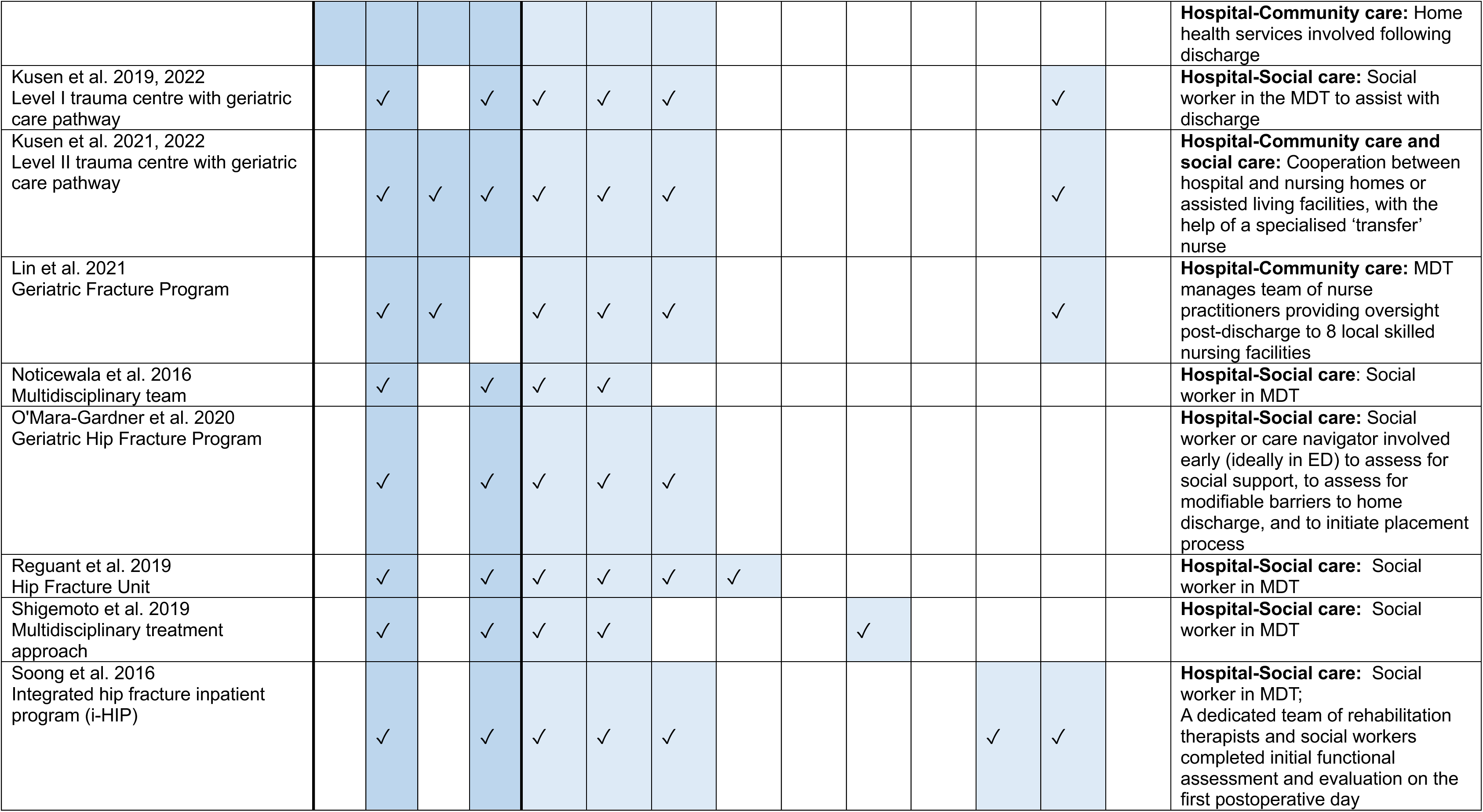
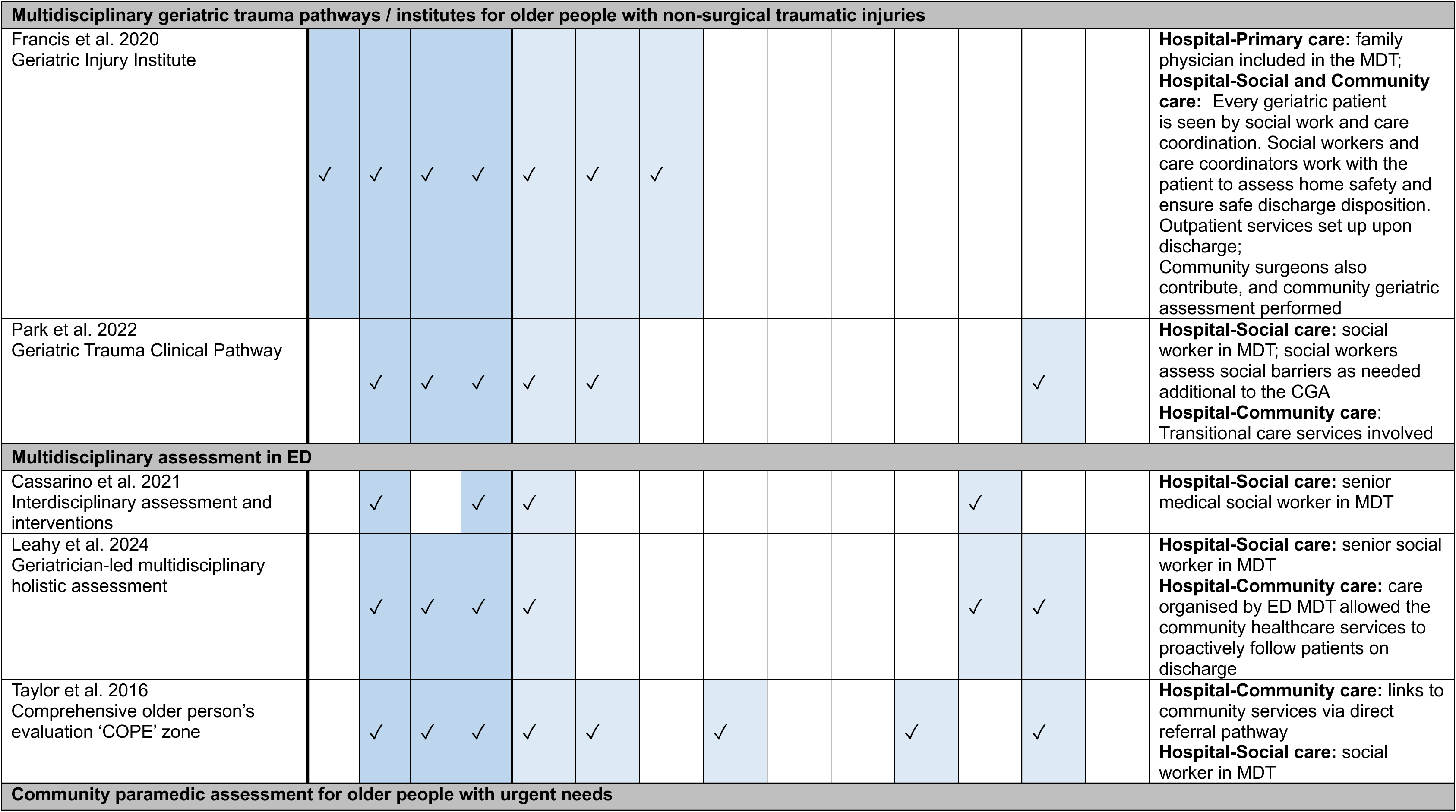
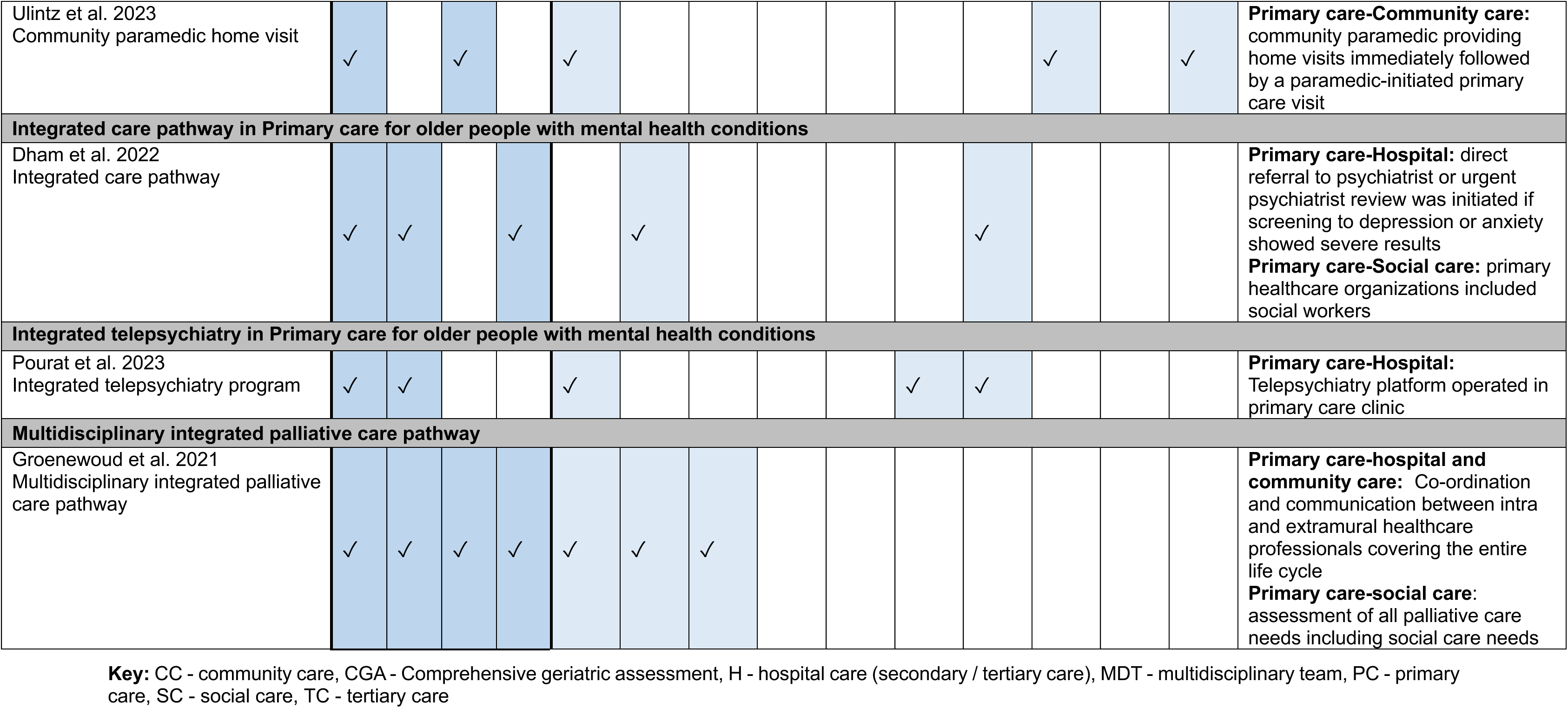
Elements of integrated care interventions from quantitative studies. (ordered by population, medical specialty, and intervention type)

| Study (Author, year)<br>Interventions | Service level integration |  |  |  | MDT | Pathway/ protocols | Care coordination | New unit | Joint patient review/ discharge | Integrated patient records | Agreed referral criteria | Joint assessment | CGA | Professional role change | Mechanism of integration between services |
| --- | --- | --- | --- | --- | --- | --- | --- | --- | --- | --- | --- | --- | --- | --- | --- |
|  | PC | H | CC | SC |  |  |  |  |  |  |  |  |  |  |  |
| Multidisciplinary orthogeriatric care for hip and other upper or lower extremity fractures |  |  |  |  |  |  |  |  |  |  |  |  |  |  |  |
| Blauth et al. 2021<br>Geriatric fracture centres |  | ✓ |  | ✓ | ✓ | ✓ |  | ✓ |  |  |  |  |  |  | **Hospital-Social care:** Social worker in MDT |
| Branas et al. 2018<br>Process Management System |  | ✓ |  | ✓ | ✓ | ✓ |  |  | ✓ |  |  |  |  |  | **Hospital-Social care:** Social worker in MDT |
| Duaso et al. 2018<br>Collaborationist orthogeriatric unit | ✓ | ✓ |  | ✓ | ✓ | ✓ | ✓ | ✓ |  |  |  |  |  |  | **Hospital-Primary care and social care:** Discharge is coordinated with primary care team and social services;<br>Social worker integrated into multi-protocol in the postoperative management phase |
| Kalmet et al. 2019<br>Multidisciplinary clinical pathway |  | ✓ | ✓ | ✓ | ✓ | ✓ |  |  |  |  |  |  |  |  | **Hospital-Community care and social care:** Agreement in place with patient-centred destination for rehabilitation (whether this is a rehabilitation centre or a nursing home with rehabilitation facilities) |
| Katrancha et al. 2017<br>Virtual Geriatric Trauma Institute (nurse-led) | ✓ | ✓ | ✓ | ✓ | ✓ | ✓ | ✓ |  |  |  |  |  |  |  | **Hospital-Primary care and social care:** Primary care physician, and staff from social services included in the MDT; |
|  |  |  |  |  |  |  |  |  |  |  |  |  |  |  | **Hospital-Community care:** Home health services involved following discharge |
| Kusen et al. 2019, 2022<br>Level I trauma centre with geriatric care pathway |  | ✓ |  | ✓ | ✓ | ✓ | ✓ |  |  |  |  |  | ✓ |  | **Hospital-Social care:** Social worker in the MDT to assist with discharge |
| Kusen et al. 2021, 2022<br>Level II trauma centre with geriatric care pathway |  | ✓ | ✓ | ✓ | ✓ | ✓ | ✓ |  |  |  |  |  | ✓ |  | **Hospital-Community care and social care:** Cooperation between hospital and nursing homes or assisted living facilities, with the help of a specialised 'transfer' nurse |
| Lin et al. 2021<br>Geriatric Fracture Program |  | ✓ | ✓ |  | ✓ | ✓ | ✓ |  |  |  |  |  | ✓ |  | **Hospital-Community care:** MDT manages team of nurse practitioners providing oversight post-discharge to 8 local skilled nursing facilities |
| Noticewala et al. 2016<br>Multidisciplinary team |  | ✓ |  | ✓ | ✓ | ✓ |  |  |  |  |  |  |  |  | **Hospital-Social care:** Social worker in MDT |
| O'Mara-Gardner et al. 2020<br>Geriatric Hip Fracture Program |  | ✓ |  | ✓ | ✓ | ✓ | ✓ |  |  |  |  |  |  |  | **Hospital-Social care:** Social worker or care navigator involved early (ideally in ED) to assess for social support, to assess for modifiable barriers to home discharge, and to initiate placement process |
| Reguant et al. 2019<br>Hip Fracture Unit |  | ✓ |  | ✓ | ✓ | ✓ | ✓ | ✓ |  |  |  |  |  |  | **Hospital-Social care:** Social worker in MDT |
| Shigemoto et al. 2019<br>Multidisciplinary treatment approach |  | ✓ |  | ✓ | ✓ | ✓ |  |  |  | ✓ |  |  |  |  | **Hospital-Social care:** Social worker in MDT |
| Soong et al. 2016<br>Integrated hip fracture inpatient program (i-HIP) |  | ✓ |  | ✓ | ✓ | ✓ | ✓ |  |  |  |  | ✓ | ✓ |  | **Hospital-Social care:** Social worker in MDT;<br>A dedicated team of rehabilitation therapists and social workers completed initial functional assessment and evaluation on the first postoperative day |

| Multidisciplinary geriatric trauma pathways / institutes for older people with non-surgical traumatic injuries |  |  |  |  |  |  |  |  |  |  |  |  |  |  |
| --- | --- | --- | --- | --- | --- | --- | --- | --- | --- | --- | --- | --- | --- | --- |
| Francis et al. 2020<br>Geriatric Injury Institute | ✓ | ✓ | ✓ | ✓ | ✓ | ✓ | ✓ |  |  |  |  |  |  | **Hospital-Primary care:** family physician included in the MDT;<br>**Hospital-Social and Community care:** Every geriatric patient is seen by social work and care coordination. Social workers and care coordinators work with the patient to assess home safety and ensure safe discharge disposition. Outpatient services set up upon discharge;<br>Community surgeons also contribute, and community geriatric assessment performed |
| Park et al. 2022<br>Geriatric Trauma Clinical Pathway |  | ✓ | ✓ | ✓ | ✓ | ✓ |  |  |  |  |  |  | ✓ | **Hospital-Social care:** social worker in MDT; social workers assess social barriers as needed additional to the CGA<br>**Hospital-Community care:** Transitional care services involved |
| Multidisciplinary assessment in ED |  |  |  |  |  |  |  |  |  |  |  |  |  |  |
| Cassarino et al. 2021<br>Interdisciplinary assessment and interventions |  | ✓ |  | ✓ | ✓ |  |  |  |  |  |  | ✓ |  | **Hospital-Social care:** senior medical social worker in MDT |
| Leahy et al. 2024<br>Geriatrician-led multidisciplinary holistic assessment |  | ✓ | ✓ | ✓ | ✓ |  |  |  |  |  |  | ✓ | ✓ | **Hospital-Social care:** senior social worker in MDT<br>**Hospital-Community care:** care organised by ED MDT allowed the community healthcare services to proactively follow patients on discharge |
| Taylor et al. 2016<br>Comprehensive older person's evaluation 'COPE' zone |  | ✓ | ✓ | ✓ | ✓ | ✓ |  | ✓ |  |  | ✓ |  | ✓ | **Hospital-Community care:** links to community services via direct referral pathway<br>**Hospital-Social care:** social worker in MDT |
| Community paramedic assessment for older people with urgent needs |  |  |  |  |  |  |  |  |  |  |  |  |  |  |
| Ulintz et al. 2023<br>Community paramedic home visit | ✓ |  | ✓ |  | ✓ |  |  |  |  |  |  | ✓ |  | ✓ | **Primary care-Community care:** community paramedic providing home visits immediately followed by a paramedic-initiated primary care visit |
| **Integrated care pathway in Primary care for older people with mental health conditions** |  |  |  |  |  |  |  |  |  |  |  |  |  |  |  |
| Dham et al. 2022<br>Integrated care pathway | ✓ | ✓ |  | ✓ |  | ✓ |  |  |  |  | ✓ |  |  |  | **Primary care-Hospital:** direct referral to psychiatrist or urgent psychiatrist review was initiated if screening to depression or anxiety showed severe results<br>**Primary care-Social care:** primary healthcare organizations included social workers |
| **Integrated telepsychiatry in Primary care for older people with mental health conditions** |  |  |  |  |  |  |  |  |  |  |  |  |  |  |  |
| Pourat et al. 2023<br>Integrated telepsychiatry program | ✓ | ✓ |  |  | ✓ |  |  |  |  | ✓ | ✓ |  |  |  | **Primary care-Hospital:** Telepsychiatry platform operated in primary care clinic |
| **Multidisciplinary integrated palliative care pathway** |  |  |  |  |  |  |  |  |  |  |  |  |  |  |  |
| Groenewoud et al. 2021<br>Multidisciplinary integrated palliative care pathway | ✓ | ✓ | ✓ | ✓ | ✓ | ✓ | ✓ |  |  |  |  |  |  |  | **Primary care-hospital and community care:** Co-ordination and communication between intra and extramural healthcare professionals covering the entire life cycle<br>**Primary care-social care:** assessment of all palliative care needs including social care needs |

**Key:** CC - community care, CGA - Comprehensive geriatric assessment, H - hospital care (secondary / tertiary care), MDT - multidisciplinary team, PC - primary care, SC - social care, TC - tertiary care

##### Multidisciplinary orthogeriatric care for hip and other upper or lower extremity fractures

The 14 studies focusing on orthogeriatric care for **hip and other upper and lower extremity fractures needing surgeries** reported 13 unique **interventions**. One study (Kusen et al. 2022) compared two interventions from two other included publications (Kusen et al. 2019, Kusen et al. 2021). All of the 13 interventions focused on **orthogeriatric models of care** provision, meaning that a geriatrician was integrated into orthopaedic or trauma specialist teams to improve care for older people with fractures. These interventions were complex, with multiple elements to achieve seamless care. Care provision was **multidisciplinary** often with a team of medical specialists, nurses, occupational therapists, physiotherapists and social workers working together. In eight interventions, **a care coordinator** was employed as part of the MDT. Transfer nurses, whose role included arranging discharge location for patients, often acted as care coordinators (Kusen et al. 2021, Kusen et al. 2022). **Pathways and protocols** determined processes, timeframes, and staff roles usually from hospital admission to the day of discharge following inpatient stay. In six studies, pathways were described to start as early as arrival at ED (Duaso et al. 2018, Kalmet et al. 2019, Kusen et al. 2021, O’Mara-Gardner et al. 2020, Reguant et al. 2019, Shigemoto et al. 2019), while in two studies care continued following discharge (Branas et al. 2018, Lin et al. 2021).

All interventions aimed to integrate different services (for example hospital and social care), although the mechanism and strength of integration varied.

- Eight interventions provided integration across hospital (secondary/tertiary) and social care (Blauth et al. 2021, Branas et al. 2018, Kusen et al. 2019, Noticewala et al. 2016, O’Mara-Gardner et al. 2020, Reguant et al. 2019, Shigemoto et al. 2019, Soong et al. 2016).
- Two interventions aimed to integrate hospital, community and social care (Kalmet et al. 2019, Kusen et al. 2021),
- One intervention was implemented across primary, hospital and social care (Duaso et al. 2018).
- One intervention supported integration across primary, hospital, community and social care (Katrancha et al. 2017).
- One intervention integrated hospital and community care (Lin et al. 2021).

The mechanisms used to achieve integration across hospital and social care varied. In eight interventions, integration between hospital and social care was facilitated by **including a social worker in the MDT,** although details about their specific roles were often lacking (Blauth et al. 2021, Branas et al. 2018, Katrancha et al. 2017, Kusen et al. 2019, Noticewala et al. 2016, Reguant et al. 2019, Shigemoto et al. 2019, Soong et al. 2016). In one intervention, social workers were integrated into the postoperative management phase of the pathway and patient discharge was coordinated with social services (Duaso et al. 2018). In two interventions, agreements were in place between nursing homes and assisted living facilities to enable timely and safe discharge (Kalmet et al. 2019, Kusen et al. 2021, Kusen et al. 2022). In two interventions, social workers were involved early in the care pathway, performing patient assessment to enable identification of social care needs and timely discharge planning (O’Mara-Gardner et al. 2020, Soong et al. 2016).

Integration across hospital and community care was often realised via agreements with community services closer to patient’s homes post hospital discharge (Kalmet et al. 2019, Katrancha et al. 2017, Kusen et al. 2021, Kusen et al. 2022, Lin et al. 2021). Integration across primary and hospital care was managed by the inclusion of primary care physician within the MDT in one intervention (Katrancha et al. 2017) and coordinating discharge with the patients’ primary care team in another (Duaso et al. 2018).

Other less commonly reported elements of the integrated care interventions for hip and other fractures included the development of **new orthogeriatric fracture units** (3 studies) (Blauth et al. 2021, Duaso et al. 2018, Reguant et al. 2019). **Joint patient discharge reports by the orthopaedic surgeon and the geriatrician** were reported in relation to one intervention (Branas et al. 2018). One intervention included the implementation of **integrated patient records** alongside a multidisciplinary care approach (Shigemoto et al. 2019). Finally, the use of **comprehensive geriatric assessments** (CGA) was reported in four interventions (Kusen et al. 2019, Kusen et al. 2021, Kusen et al. 2022, Lin et al. 2021, Soong et al. 2016).

##### Multidisciplinary geriatric trauma pathways / institutes for older people with non-surgical traumatic injuries

Two studies reported two unique integrated care interventions for **older people experiencing non-surgical traumatic injuries** (from falls, motor vehicle collision or other causes, but injury not specified) (Francis et al. 2020, Park et al. 2022). The main element of both interventions was **MDT** working alongside the implementation of **standardised pathways and protocols**. The pathways spanned care from ED arrival to discharge from hospital following inpatient stay. Additionally, other services were also integrated with the in-hospital trauma pathways. One of the interventions aimed to integrate primary, hospital, community and social care (Francis et al. 2020). This was achieved by including family physicians and community surgeons in the MDT, while a social worker acted as **care coordinator** to ensure safe home discharge and the setting up of outpatient support (Francis et al. 2020). The integrated care intervention by Park et al. (2020) aimed to support collaboration across hospital, community and social care (Park et al. 2022). Integration of hospital and social care was achieved by including a social worker in the MDT who was responsible for the assessment of social barriers, while transitional care services were involved to ensure joining up hospital and community care services (Park et al. 2022).

##### Multidisciplinary assessment in ED

Three studies reported three unique integrated care interventions for **older people presenting to the ED with a range of conditions** such as frailty, falls, dementia, delirium, general unwellness, limb problems, back pain, urinary symptoms, and ear or facial issues (Cassarino et al. 2021, Leahy et al. 2024, Taylor et al. 2016). The integrated care interventions focused on the assessment of patients within the ED which could end either in discharge with recommendations or admission to the hospital. Two interventions focused on the provision of **joint assessment** of older people by an MDT situated in the ED (Cassarino et al. 2021, Leahy et al. 2024), out of which one explicitly reported the use of CGA (Leahy et al. 2024). Both MDTs included physicians, nurses, physiotherapists, occupational therapists as well as a social worker to enable integration across hospital and social care. Additionally, one MDT organised follow-on care via community services (Leahy et al. 2024).

In one intervention a comprehensive older person’s evaluation (COPE) zone in an ED was developed (Taylor et al. 2016). The COPE zone was established as a **new unit** run by an **MDT** integrating a geriatrician into an acute care team within the ED. The COPE zone had an **agreed referral criteria** (falls, delirium, dementia or care home/intermediate care residents) and provided comprehensive geriatric assessment, although it was not mentioned whether assessment was performed jointly by the MDT. The COPE zone had a direct referral pathway to community services and included a social worker within the MDT ensuring integration across hospital, community and social care (Taylor et al. 2016).

##### Community paramedic assessment for older people with urgent needs

One integrated care intervention focused on assessment by a community paramedic followed by a paramedic initiated primary care physician telemedicine visit for older people with urgent needs (Ulintz et al. 2023). **Professional role change** was key element of this interventions, enabling community paramedics to provide assessments that were previously solely provided by primary care physicians, while integrating community and primary care services.

##### Integrated care pathway in primary care for older people with mental health conditions

**Mental health** support for older people with anxiety, depression, mild cognitive impairment was the focus of two integrated care interventions, out of which one focused on integrated care **pathways** starting from primary care. This integrated care pathway aimed to integrate primary, hospital, and social care (Dham et al. 2022). An agreed referral criteria based on screening guided family physicians’ decision making regarding the mental health support necessary for the patient, which included cognitive behaviour or brief psychological therapy. Direct referral path to psychiatrists was also available for people with serious mental health issues integrating primary and hospital care. Social care integration was achieved by including social workers in the primary care team.

##### Integrated telepsychiatry in primary care for older people with mental health conditions

One integrated care intervention focused on the provision of mental health support for older people via the use of telepsychiatry platforms within primary care clinics (Pourat et al. 2023). This enabled integration between primary and hospital care, with primary care providers having direct access to psychiatrist support. Integrated patient records further enabled collaboration between services by enabling direct communication between primary physicians and psychiatrists and improving documentation efficiency (Pourat et al. 2023).

##### Multidisciplinary integrated palliative care pathway

One intervention aimed to provide **integrated palliative care to older people** with advanced or life-limiting illnesses, although the specific conditions were not described (Groenewoud et al. 2021). The pathway enabled integration across primary, hospital, community and social care via a MDT that focused on early identification of palliative care needs and coordination of care across multiple providers.

#### 2.1.4 Overview of waiting time outcomes

The quantitative studies reported a range of different waiting time outcomes which can be categorised based on setting into inpatient, emergency, and routine care. **Inpatient waiting times** were the most frequently reported and within this category time to surgery was reported across 14 studies. Time to surgery was mostly measured in hours or days (12 studies), but some studies also calculated the percentage of patients undergoing surgery within specific timeframes such as 48 hours (4 studies) (Branas et al. 2018; O’Mara-Gardner et al. 2020; Reguant et al. 2019; Shigemoto et al. 2019). Additionally, two studies reported on the timing of surgery by day of admission (Kusen et al. 2021, Kusen et al. 2022). Other inpatient waiting times included time to first goals-of-care discussion (Park et al. 2022), time to geriatric medicine evaluation (Francis et al. 2020), and time to physical therapy (Francis et al. 2020). These outcomes were measured from hospital or ward admission to the time of the assessment or procedure, usually in hours or minutes.

**Emergency waiting times** were reported in six studies. These included time to hospital or ward admission (Blauth et al. 2021, Branas et al. 2018), ED stay (Cassarino et al. 2021, Francis et al. 2020, Leahy et al. 2024), time until geriatric review (Taylor et al. 2016). Time to hospital or ward admission was measured slightly differently in the two studies. One study measured this as time from injury to hospital admission (Blauth et al. 2021), while the other captured time from ED to admission to the ward (Branas et al. 2018). The outcome of ED stay covered the time period spent on ED until admission or discharge (Cassarino et al. 2021, Francis et al. 2020, Leahy et al. 2024), while time until geriatric review measured a specific period spent in an emergency assessment unit waiting for CGA.

**Routine care waiting times** were reported in four studies, namely time to treatment initiation (Dham et al. 2022), time to appointment (Pourat et al. 2023), primary care waiting times (Ulintz et al. 2023) and quality of care reported by GPs (Groenewoud et al. 2021). Time to treatment initiation was calculated as the period between screening and the date of expected intervention (Dham et al. 2022). Time to appointment referred to the number of days to telepsychiatry appointment, which could further be split into waiting time for new and returning patients (Pourat et al. 2023). Primary care wait time was calculated as time between the phone call requesting an appointment and the in-home visit (Ulintz et al. 2023). Quality of care captured GPs’ perspectives on the timeliness of patient needs assessment and subsequent palliative care interventions (Groenewoud et al. 2021).

### 2.2 Effectiveness of integrated care interventions in reducing waiting times/lists

The quantitative component of this review aims to determine the effectiveness of integrated care in reducing waiting times and/or lists for older people. In this section, the effectiveness of different integrated care interventions is reported, with findings grouped according to population, medical specialty, type of intervention, and waiting time outcomes.

#### 2.2.1 Effectiveness of multidisciplinary orthogeriatric care for older people with hip and other upper and lower extremity fractures needing surgery

In this section the effect of 13 integrated care interventions for older people with hip and other upper and lower extremity fractures is presented. All 13 interventions were **orthogeriatric care**, which is a complex intervention that involved a dedicated MDT of geriatricians, orthopaedic or trauma specialist surgeons, nurses, occupational therapists, physiotherapists and social workers working together. In eight interventions, **a care coordinator** was employed as part of the MDT. **Pathways** determined processes, timeframes, and staff roles usually from hospital admission to the day of discharge following inpatient stay. The studies investigated waiting times, such as time to admission (measured in days) and time to surgery (measured in days and hours, percentage of patients undergoing surgery in 48 hours, or the difference in day of surgery).

##### Time to admission (days)

Out of the 12 studies that investigated integrated care interventions for hip fracture, two reported time to hospital admission: one medium quality cohort study (Blauth et al. 2021), and one low quality uncontrolled before and after study (Branas et al. 2018). The results were mixed with the uncontrolled before and after study reported a statistically significant change, while the cohort study found no statistically significant difference. Blauth et al. (2021) investigated the difference in the time from injury to hospital admission between geriatric fracture centres and usual care centres across a range of countries (Austria, Spain, USA, Netherlands, Singapore and Thailand). While the average number of days to admission was slightly less in geriatric fracture centres (Mean 1.0 ± 4.1 days) compared to usual care centres (Mean 1.2 ± 5.5 days), this difference was not statistically significant (p=0.270).

Branas et al. (2018) compared time to admission to the ward from the ED before and after the implementation of an improved process management system in a public university hospital in Spain. The original care model was orthogeriatric co-management, although the process management system helped further specify and streamline the pathway and establish regular MDT meetings. The results show that following the implementation of the process management system, the average hours to ward admission from the ED statistically significantly decreased (Mean 11.8 ± 11.2 days) compared to the original orthogeriatric co-management model (Mean 15.9 ± 17.6 days) (p=0.0001).

##### Time to surgery (days or hours)

Ten studies that focused on hip fracture collected data on time to surgery (measure in days and hours), and seven studies reported statistically significant improvements in this outcome: two medium quality cohort studies (Blauth et al. 2021, Noticewala et al. 2016), one medium quality controlled before and after study (Soong et al. 2016), and four medium to low quality uncontrolled before and after studies (Branas et al. 2018, Duaso et al. 2018, O’Mara-Gardner et al. 2020, Reguant et al. 2019).

The cohort study by Blauth et al. (2021) found that the time from admission to surgery was statistically significantly shorter in geriatric fracture centres (median 28 hours) compared to usual care centres across multiple countries (median 43 hours) (p<0.001). Noticewala et al. (2016) compared care provided by a MDT in a small satellite hospital to usual orthopaedic team care at the wider tertiary medical centre in the USA. The average number of days to surgery was statistically significantly shorter for care provided by the MDT (mean 1.7 ± 1.8 days) in comparison to the orthopaedic team (Mean 2.4 ± 2.2 days) (p=0.0004). One controlled before and after study conducted by Soong et al. (2016) found that the average number of hours to surgery statistically significantly reduced after an inpatient hip fracture program (i-HIP) was implemented in an acute care urban academic health sciences centre in Canada (pre Mean 45.8 ± 66.8 hours vs post mean 29.7 ± 17.9 hours; p<0.001).

An uncontrolled before and after study by Branas et al. (2018) found that the average hours of preoperative stay was statistically significantly shorter following the implementation of a process management system (pre mean 88.1 ± 64 hours vs post mean 66.4 ± 53.9 hours; p=0.0001). Another study of similar design reported that the mean number of days from admission to surgery statistically significantly reduced following the implementation of a collaborationist orthogeriatric unit (mean 1.86 ± 1.19 days) compared to the previously provided traditional trauma ward model in the same hospital in Spain (mean 2.70 ± 1.79 days) (p=0.0001) (Duaso et al. 2018). O’Mara-Gardner et al. (2020) found that time to surgery was statistically significantly shorter following the implementation of a geriatric hip fracture programme and care navigator in a level I trauma centre in the USA (pre mean 30.23 ± 29.5 hours vs post mean 22.79 ± 12 hours, p<0.0001). Finally, a study from Spain evaluated the implementation of a multidisciplinary hip fracture unit and compared it to the previous standard care that was managed by an orthopaedic surgeon (Reguant et al. 2019). Following the implementation of the hip fracture unit, surgical delay was statistically significantly reduced (pre median 3 days vs post median 2 days; p=0.001) (Reguant et al. 2019).

Three medium to low quality uncontrolled before and after studies reported no statistically significant difference or no change in time to surgery (Katrancha et al. 2017, Shigemoto et al. 2019, Kusen et al. 2019). The study by Katrancha et al. (2017) evaluated the implementation of a nurse-led virtual geriatric trauma institute in a level I trauma centre in the USA and found no statistically significant difference neither in mean days to surgery (pre mean 1.2 days ± 0.75 vs post mean 1.1 ± 0.71 days; p=0.3) nor in mean hours to surgery (pre mean 28.6 ± 17.92 hours vs post mean 27.0 ± 17.15 hours; p=0.3). Shigemoto et al. (2019) compared a newly implemented multidisciplinary treatment approach to the previous conventional hip fracture care within the same hospital in Japan. While slight reduction in average hours to surgery was reported, this was not statistically significant (pre mean 36 hours 29 minutes vs post mean 33 hours and 22 minutes; p=0.459). Finally, Kusen et al. (2019) compared time to surgery in a level I trauma centre in Switzerland before and after the implementation of a geriatric care pathway. while median time to surgery prior to the geriatric care pathway being put in place was shorter (median 15 hours 34 minutes) compared to post-implementation (median 18 hours and 51 minutes), this difference was not statistically significant (p=0.32).

##### Time to surgery (surgery percentage within 48 hours)

Four medium to low quality uncontrolled before and after studies measured percentage of patients undergoing surgery within 48 hours from injury or admission, and three reported statistically significant increase following implementation of a multidisciplinary intervention (Branas et al. 2018, O’Mara-Gardner et al. 2020, Reguant et al. 2019). Branas et al. (2018) measured the percentage of patients undergoing operation within 48 hours and found that it increased following implementation of the process management system in Spain (pre 33.7% vs post 50.8%, p=0.0001). The study by O’Mara-Gardner et al. (2020) reported a statistically significant increase in the percentage of patients undergoing surgery within 24 hours (pre: 42.2% vs post: 67.2%) and within 48 hours (pre: 82.3% vs post: 97.0%) following the implementation of a geriatric hip fracture program in the USA (p < 0.0001). Finally, Reguant et al. (2019) measured the percentage of patients who underwent surgery within 48 hours, and operation was performed on a higher percentage of patients following the implementation of the hip fracture unit in Spain (pre 38.3% vs post 55.1%; p<0.001). In contrast, Shigemoto et al. (2019) found a slight reduction in the percentage of patients operated on within 48 hours in Japan, although this difference was not statistically significant (pre=75.2% vs post=72.5%; p=0.485).

##### Time to surgery (difference in day of surgery)

Two low quality cohort study investigated on which day from admission patient were operated on: the day of admission, the first day of admission or the second day of admission (Kusen et al. 2021, Kusen et al. 2022). In the study by Kusen et al. (2021), care in two different level II trauma centres within the Netherlands were compared, one providing geriatric care pathways and the other providing standard care system. Results showed that although the percentage of patients receiving surgical interventions on the first and second day of admission was statistically significantly higher in the geriatric care pathway, a statistically significantly greater proportion of patients underwent surgery on the day of admission under the standard care system (geriatric: 18.5% vs standard: 32.3%; p < 0.0001).

Kusen et al. (2022) also compared geriatric care pathways across Switzerland and the Netherlands, with interventions and settings (level I and level II trauma centres) selected from their previous two publications (Kusen et al. 2019, Kusen et al. 2021). No statistically significant difference in the percentage of patients undergoing surgery on the day of admission, on the first or second day were reported (p=0.15). However, a higher percentage of patients underwent surgery on the day of admission in the level I trauma centre (23.4%) compared to the level II trauma centre (18.5%).

#### 2.2.2 Effectiveness of multidisciplinary geriatric trauma pathways / institutes

This section presents the effectiveness of two integrated care interventions for older people experiencing non-surgical traumatic injuries. The main element of the interventions was MDT working alongside the implementation of standardised pathways which spanned care from ED arrival to discharge from hospital following inpatient stay. Various waiting time outcomes were measured by the studies, including emergency department stay (measure in minutes), time to geriatric medicine evaluation (measured in hours), time to physical therapy (measured in hours), and time to first goals-of-care assessment (measured in hours).

##### Emergency department stay (minutes)

One medium quality uncontrolled before and after study investigated ED stay (time prior to admission) for older patients with trauma ((fall, motor vehicle collision, bicycle, or other) (Francis et al. 2020). Francis et al. (2020) found that while ED stay reduced following the implementation of a multidisciplinary geriatric institute within a tertiary care hospital in the USA, this change was not statistically significant (pre mean 310.7 ± 602.9 minutes vs post mean 219.8 ± 141.6 minutes; p=0.054).

##### Time to geriatric medicine evaluation (hours)

One medium quality uncontrolled before and after study investigated time to geriatric medicine evaluation (Francis et al. 2020). While average hours to geriatric medicine evaluation reduced following the implementation of a multidisciplinary geriatric institute, this change was not statistically significant (pre mean 5.1 ± 5.82 hours vs post mean 4.5 ± 3.83 minutes; p=0.594).

##### Time to physical therapy (hours)

One medium quality uncontrolled before and after study investigated time to physical therapy (Francis et al. 2020). Time to physical therapy was shorter following the implementation of a multidisciplinary geriatric institute, although this change was not statistically significant (pre mean 52.1 ± 50 hours vs post mean 51.6 ± 50.2 minutes; p=0.926).

##### Time to first goals-of-care assessment (hours)

One medium-quality uncontrolled before-and-after study conducted in a Level I trauma centre in the USA found that the time to first goals-of-care assessment statistically significantly decreased following the implementation of a geriatric trauma clinical pathway (pre: 49.6 ± 105.5 hours vs post: 35.7 ± 25.3 hours; p = 0.03) (Park et al. 2022).

#### 2.2.3 Effectiveness of multidisciplinary assessment in an emergency department

This section covers the effectiveness of three unique integrated care interventions for older people presenting to the ED with a range of conditions (Cassarino et al. 2021, Leahy et al. 2024, Taylor et al. 2016). All three interventions focused on the assessment of patients within the ED managed by an MDT. The interventions solely focused on assessment in ED, ending in discharge or admission to hospital. The outcomes investigated included emergency department stay (measured in hours), and time until geriatric review (measured in days).

##### Emergency department stay (hours)

Two high quality randomised controlled studies examined the impact of multidisciplinary assessments within EDs on patient flow metrics and both studies reported statistically significant reductions in the time patients spent in the ED prior to admission or discharge. (referred to as patient experience time or duration of stay in ED). (Cassarino et al. 2021, Leahy et al. 2024). Cassarino et al. (2021) reported a statistically significant reduction in ED stay duration for patients receiving interdisciplinary assessments (median 6.43 hours) compared to those receiving routine care (median 12.1 hours; p < 0.001). Similarly, Leahy et al. (2024) found that patients in the geriatrician-led multidisciplinary assessment group had a statistically significantly shorter ED stay (median 11.5 hours) than those in the usual care group (median 20 hours; p = 0.013).

##### Time until geriatric review (days)

A medium-quality uncontrolled before-and-after study by Taylor et al. (2016) assessed the impact of implementing the Comprehensive Older Person’s Evaluation (COPE) zone within the emergency assessment unit. The study found a statistically significant reduction in the time to geriatric assessment, decreasing from a mean of 0.85 days pre-implementation to 0.48 days post-implementation (p < 0.001). In a subgroup analysis of patients with frailty markers greater than 1, a similar statistically significant reduction was observed, with time to geriatric review decreasing from a mean of 0.88 days to 0.49 days (p = 0.001).

#### 2.2.4 Effectiveness of community paramedic assessment for older people with urgent needs

This section presents the effectiveness of one integrated care intervention focused on assessment by a community paramedic which was followed by a paramedic initiated primary care physician telemedicine visit for older people with urgent needs (Ulintz et al. 2023). The outcome of interest was primary care wait time (measured in days).

##### Primary care wait time

One low quality prospective cohort study found that primary care wait time was statistically significantly shorter in the community paramedic assessment group (median 1 day) compared to the usual in-person primary care provider visit (median 5 days) for older people with urgent needs in the USA (p<0.001) (Ulintz et al. 2023).

#### 2.2.5 Effectiveness of an integrated care pathway in primary care for older people with mental health conditions

The effectiveness of a care pathway aimed to integrate primary, hospital, and social care for older people with mental health conditions is the focus of this section (Dham et al. 2022).

The integrated care pathway had an agreed referral criteria based on screening that guided family physicians decision making regarding the mental health support necessary for the patient, including direct contact with a psychiatrist. The outcome investigated was time to treatment initiation.

##### Time to treatment initiation (Hazard ratio^1^)

One medium quality non-randomised controlled trial investigated integrated care pathways across five primary care practices in Canada and their impact on time to treatment initiation (Dham et al. 2022). Participants in the integrated care pathway were 3.56 times more likely to start treatment early compared to the treatment as usual group (hazard ratio 3.557 (95% ci [2.228, 5.678]) p<0.001). In addition, a subgroup analysis focusing on participants experiencing anxiety and depression, the likelihood of early treatment initiation was 4.35 times higher (hazard ratio 4.353 (95% CI [1.993, 9.506]) p=0.002).

#### 2.2.6 Effectiveness of integrated telepsychiatry in Primary care for older people with mental health conditions

In this section the effectiveness of one integrated care intervention using of telepsychiatry platforms within primary care clinics for older people with mental health conditions (Pourat et al. 2023). The intervention enabled integration between primary and hospital care, with primary care providers having direct access to psychiatrist support. The outcome of interest was days to appointment.

##### Days to appointment

One low quality uncontrolled before and after study presented that the mean number of days to a psychiatry appointment reduced from 75 days to 6 for new patients who accessed care via telepsychiatry in primary care as opposed to the usual in-person care in the USA (Pourat et al. 2023). For returning patients, the average number of days to appointment decreased from 30 days to 5 days following the implementation of telepsychiatry in primary care.

However, no statistical test was performed, thus statistical significance cannot be confirmed.

#### 2.2.7 Effectiveness of a multidisciplinary integrated palliative care pathway

This section covers one intervention aimed to provide **integrated palliative care to older people** with advanced or life-limiting illnesses (Groenewoud et al. 2021). The pathway enabled integration across primary, hospital, community and social care via a MDT that focused on early identification of palliative care needs and coordination of care across multiple providers. The investigated outcome was GP’s perceived quality of care.

##### GP perceived quality of care (self-reported)

One low quality controlled before and after study investigated GPs’ perceived quality of care across 21 primary care facilities in the Netherlands (Groenewoud et al. 2021). Based on GPs’ questionnaire responses, integrated palliative care helped patients to receive statistically significantly more timely investigation of their needs and desires (94.6%) compared to usual primary care (78.9%) (p=0.03). A statistically significantly higher percentage of GPs responded that palliative care was timely given in the multidisciplinary integrated care pathway (91.9%) compared to usual care (77.5%) (p=0.042). Additionally, GPs reported to be more proactive in the integrated care pathway (97.3%) compared to usual care (78.9%) (p=0.005).

#### 2.2.8 Bottom line summary

The identified evidence was conducted in eight different countries, with the USA contributing the highest number of research studies (eight). The evidence was mainly focused on hip and other upper and lower extremity fractures (14 studies). The majority (14 studies) reported on inpatient waiting times, such as time to surgery (measured in days and hours) (12 studies). While some evidence also focused on emergency (six studies) and routine care waiting times (four studies), the number of studies were much lower, indicating a lack of evidence on the effectiveness of integrated care interventions on routine care diagnostic and elective treatment waiting times. All interventions aimed to integrate different services (for example hospital and social care), although the mechanism and strength of integration varied. Main elements of the integrated care interventions were MDT working, development of pathways and protocols, and care coordination. Findings for each intervention and outcome were graded^2^ and are summarised below and in Table 3.

- Weak international evidence (grade CII) from two studies suggests that **<u>multidisciplinary orthogeriatric care</u>** may improve **time to admission** for older people with hip fracture, although results were not consistently statistically significant.
- Weak international evidence (grade CII) from 12 studies suggests that **<u>multidisciplinary orthogeriatric care</u>** may improve **time to surgery** for older people with hip and other upper and lower extremity fractures, although two studies report no change, showing signs of inconsistency across the findings.
- Weak international evidence (grade CII) found that **<u>multidisciplinary orthogeriatric</u> <u>care</u>** for older people with hip fracture increased the **percentage of patients undergoing surgery within 48 hours in** three studies and reduced in one, leading to inconclusive results.
- Weak evidence (grade CII) from one study from the Netherlands shows that **<u>multidisciplinary orthogeriatric care</u>** for older people with hip fracture may not increase **percentage of surgeries performed on the day of admission**.
- Weak evidence (grade CII) from one US study, although not statistically significant, suggests that a **<u>multidisciplinary geriatric institute</u>** may **reduce ED stay, time to geriatric medicine evaluation and time to physical therapy** for older patients experiencing non-surgical traumatic injuries.
- Weak evidence (grade CII) from one US study shows that **a <u>multidisciplinary</u> <u>geriatric pathway</u>** may reduce **time to goals-of-care assessment** for older people with non-surgical traumatic injury.
- Strong evidence (grade AII) from two studies from Ireland shows that a **<u>multidisciplinary assessment</u>** for older people with various concerns, is effective in reducing **time spent in the ED.**
- Weak evidence (grade CII) from one UK study shows that a **<u>dedicated</u> <u>multidisciplinary assessment zone within the ED</u>** may reduce the number of days that an older person has to **wait for a geriatric review**.
- Weak evidence (grade CII) from one US study indicates that community paramedic assessment combined with a primary care telemedicine visit may decrease **primary care wait time** for older people with urgent needs compared to usual primary care provider home visit.
- Weak evidence (grade CII) from one Canadian study indicates that an **<u>integrated</u> <u>care pathway starting from primary care</u>** may increase the likelihood of **earlier treatment initiation** for older people with mental health conditions.
- Weak evidence (grade CII) from one US study suggests that **<u>integrated</u> <u>telepsychiatry</u>** in primary care may reduce **time to appointment** for older people with mental health conditions, although no statistical analysis was conducted.
- Weak evidence (grade CII) from one study from the Netherlands found that quality of care, including **self-reported timely investigation of concerns and care provision**, may improve following the implementation of **<u>multidisciplinary palliative</u> <u>care</u>**.

**Table 3:**
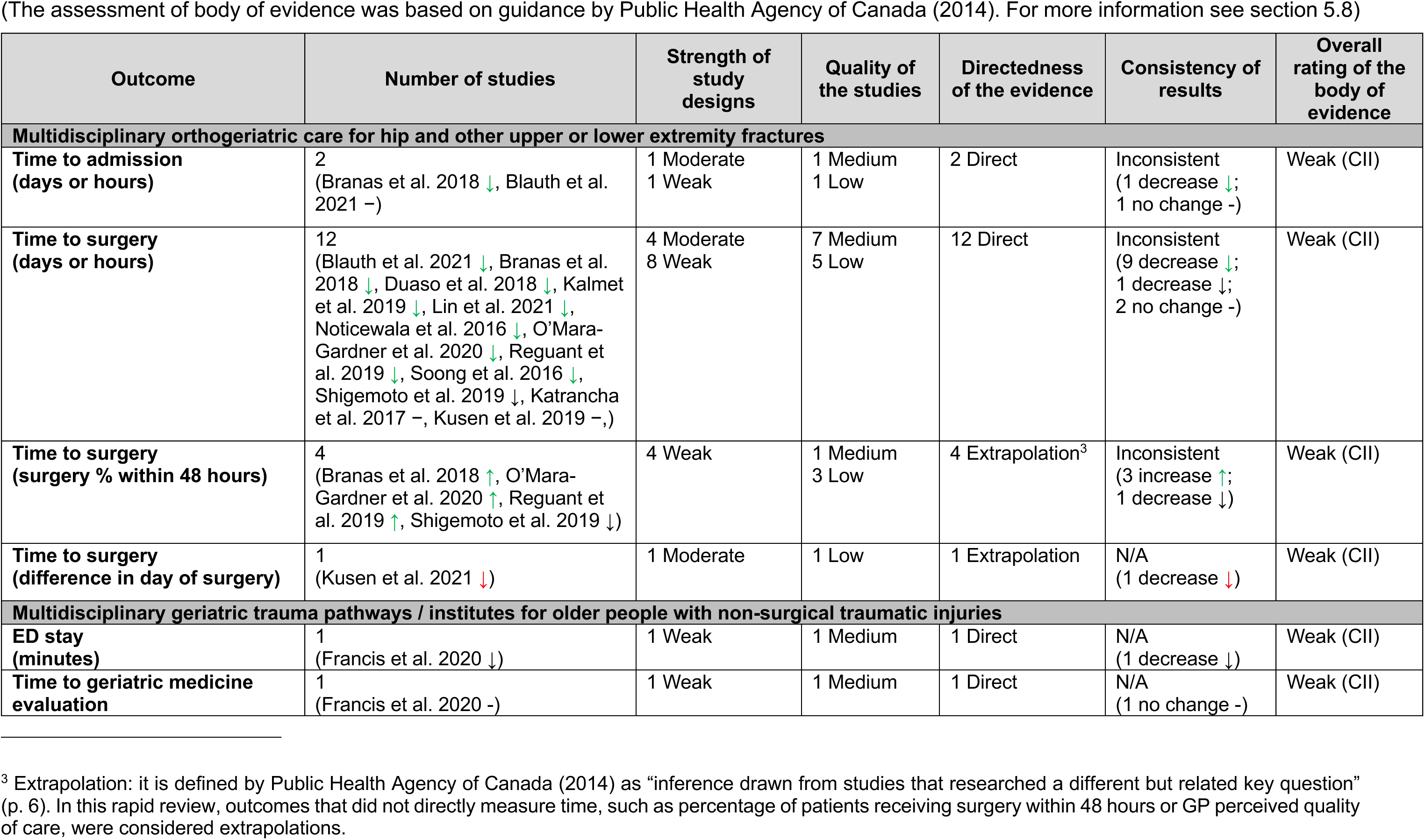
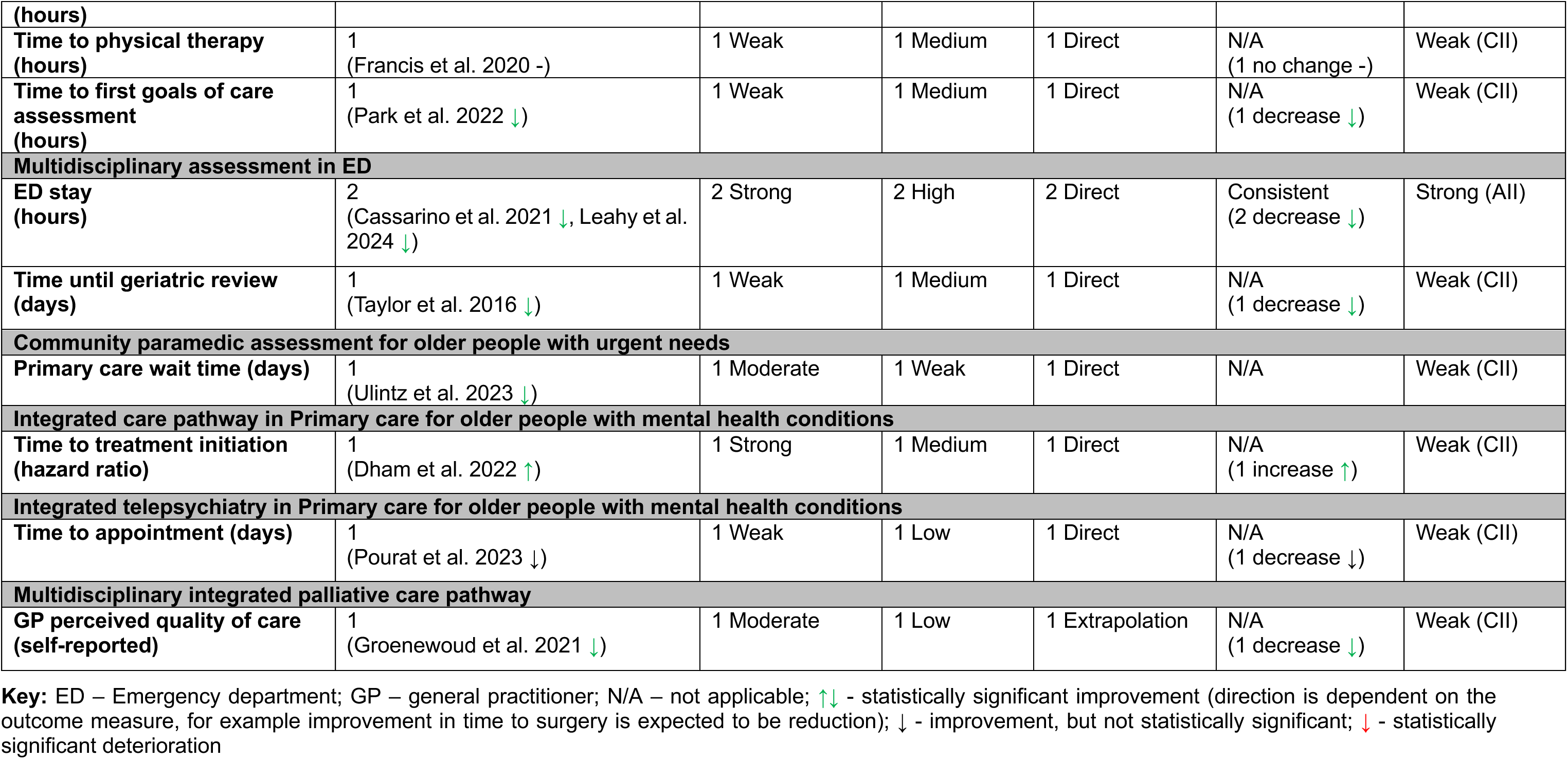
Summary of the rating of the body of the evidence.

### 2.3 Qualitative review of waiting time experiences in the context of integrated care

This section addresses the qualitative component of this review that aims to explore the views of healthcare professionals and older people regarding waiting times in the context of integrated care. The section starts with an overview of the identified qualitative studies, summarising study designs, country of origin, and the population focus. Then quality of the studies is presented, followed by the characteristics of the integrated care interventions in the focus of the qualitative studies. Waiting time categories covered by the studies is described and finally results of each study and experiences of waiting times in the context of integrated care interventions is reported.

#### 2.3.1 Overview of the evidence base

From the 30 studies with integrated care interventions across two or more services, four had qualitative descriptive designs (Aberg & Ehrenberg 2017, Fox et al. 2023, Greene et al. 2023, Luckett et al. 2017). Two studies had a mixed-methods design (including both qualitative and quantitative data) with one conducting structured interviews (Lee et al. 2015) and the other clinical observations as the qualitative arm of the research (Chow et al. 2015). One study was a descriptive survey with open-ended responses (Lee et al. 2017). Out of the four qualitative descriptive designs, one used focus groups (Aberg & Ehrenberg 2017), and three used semi-structured interviews (Fox et al. 2023, Greene et al. 2023, Luckett et al. 2017). The participants, whose perspectives were explored, were healthcare professionals (6 studies) (Aberg & Ehrenberg 2017, Chow et al. 2015, Fox et al. 2023, Lee et al. 2015, Lee et al. 2017, Luckett et al. 2017) and patients and relatives (1 study) (Greene et al. 2023).

Two qualitative descriptive studies were part of larger projects involving randomised controlled trials (Luckett et al. 2017) and service evaluations (Greene et al. 2023).

The seven studies were conducted in:

- Australia (three studies) (Chow et al. 2015, Greene et al. 2023, Luckett et al. 2017)
- Canada (two studies) (Lee et al. 2015, Lee et al. 2017)
- Sweden (one study) (Aberg & Ehrenberg 2017)
- United Kingdom (one study) (Fox et al. 2023)

The qualitative studies focused on a range of populations including older people with dementia (two studies) (Lee et al. 2017, Luckett et al. 2017), complex chronic geriatric diseases (one study) (Lee et al. 2015), urgent but non-emergency care needs (one study) (Greene et al. 2023), aging associated diseases (one study) (Aberg & Ehrenberg 2017), aged care needs (one study) (Chow et al. 2015) and hip fractures (one study) (Fox et al. 2023). The detailed characteristics of each included qualitative studies with social care involvement can be found in Section 6.2.

#### 2.3.2 Quality of the qualitative studies

Qualitative evidence was appraised using the JBI checklist for qualitative research (Lockwood et al. 2015). Most studies (five studies) showed a clear alignment between the research methodology, the research questions, and the data collection methods ensuring that these were appropriate for addressing the research aims. Six studies provided examples from the interview, focus group or survey data to support the researchers’ interpretation. However, there were notable areas where transparency in methodology was not reported. All seven included studies had unclear congruity between their stated philosophical perspective and the research methodology. Five studies did not clearly declare the beliefs and values of the researcher, while none of the seven studies reported the researchers’ influence on the data collection and analysis process, impacting the confirmability of the findings.

Overall, the quality of the qualitative evidence was variable. Two studies met seven out of 10 quality criteria on the JBI checklist (Aberg & Ehrenberg 2017, Greene et al. 2023). Three studies (Fox et al. 2023, Lee et al. 2015, Luckett et al. 2017) were rated six out of 10, and one study met four criteria out of 10 (Lee et al. 2017). The lowest number of criteria was three out of 10 (Chow et al. 2015). Detailed results from the critical appraisal can be found in section 6.3

#### 2.3.3 Characteristics of integrated care interventions

Included studies were initially grouped according to the target population and medical specialty they focused on, namely older people with dementia, complex chronic geriatric conditions, urgent but non-emergency care, aging associated diseases and aged care, and hip fracture. The integrated care interventions comprised of a combination of elements, such as MDTs, pathways or protocols, care coordination, new unit or co-location, joint patient review or discharge, integrated patient records, agreed referral criteria, joint assessment, CGA, and professional role change. These intervention elements are detailed in section

2.1.3. The elements of each integrated care intervention are summarised in Table 4 Table 3. A detailed description of the integrated care interventions is provided below.

**Table 4:** Elements of integrated care interventions from qualitative studies.

| Authors<br>Interventions | Service level integration |  |  |  | MDT | Pathway/ protocols | Care coordination | New unit / Co- location | Joint patient review/ discharge | Integrated patient records | Agreed referral criteria | Joint assessment | CGA | Professional role change | Mechanism of integration between services |
| --- | --- | --- | --- | --- | --- | --- | --- | --- | --- | --- | --- | --- | --- | --- | --- |
|  | PC | H | CC | SC |  |  |  |  |  |  |  |  |  |  |  |
| Multidisciplinary care for older people with dementia and complex chronic geriatric conditions |  |  |  |  |  |  |  |  |  |  |  |  |  |  |  |
| Lee et al. 2015<br>Expansion of a multidisciplinary collaborative memory clinic model to complex chronic geriatric conditions | ✓ | ✓ | ✓ | ✓ | ✓ | ✓ | ✓ |  | ✓ | ✓ | ✓ | ✓ |  | ✓ | **Primary care-Hospital:**<br>Geriatric specialist physicians are linked with a primary care clinic to facilitate access to consultation support;<br>**Primary care-Community care:**<br>Primary care coordinating community care services;<br>**Primary care-social care:** Social worker in MDT |
| Lee et al. 2017<br>Multidisciplinary collaborative memory clinic model | ✓ | ✓ | ✓ | ✓ | ✓ | ✓ | ✓ |  | ✓ | ✓ | ✓ | ✓ |  | ✓ | **Primary care-Hospital:**<br>Geriatric specialist physicians are linked with a primary care clinic to facilitate access to consultation support;<br>**Primary care-Community care:**<br>Primary care coordinating community care services;<br>**Primary care-social care:** Social worker in MDT |
| Luckett et al. 2017<br>Multidisciplinary input into case conferences | ✓ |  | ✓ | ✓ | ✓ |  | ✓ |  |  |  | ✓ |  |  |  | **Social care-Primary care & community care:**<br>Palliative care planning coordinator engaging GPs and other community services in case conferences set up in nursing homes |
| **Integration of primary and community care for older people with aged care needs** |  |  |  |  |  |  |  |  |  |  |  |  |  |  |  |
| Chow et al. 2015<br>Triple I (Hub): integrated intake, information and intervention service | ✓ |  | ✓ |  | ✓ | ✓ | ✓ | ✓ |  |  |  | ✓ |  |  | **Primary care-Community care:** case managers working collaboratively with primary care practitioners |
| **Multidisciplinary hospital based geriatric care for older people with aging associated diseases** |  |  |  |  |  |  |  |  |  |  |  |  |  |  |  |
| Aberg and Ehrenberg. 2017<br>Interdisciplinary hospital based geriatric care |  | ✓ |  | ✓ | ✓ | ✓ | ✓ |  | ✓ |  | ✓ | ✓ | ✓ |  | **Hospital-Social care:**<br>Social worker in MDT |
| **Multidisciplinary hip fracture care pathway for older people** |  |  |  |  |  |  |  |  |  |  |  |  |  |  |  |
| Fox et al. 2023<br>Multidisciplinary care across multiple hospital departments |  | ✓ | ✓ |  | ✓ | ✓ | ✓ |  | ✓ | ✓ |  | ✓ |  |  | **Hospital-community care:** shared patient information system between hospital and community care for successful management of discharge |
| **Emergency department avoidance service for older people with urgent but non-emergency care needs** |  |  |  |  |  |  |  |  |  |  |  |  |  |  |  |
| Greene et al. 2023<br>Emergency department avoidance service (CARE) |  | ✓ |  | ✓ | ✓ |  |  | ✓ |  |  | ✓ |  |  |  | **Hospital-Social care:**<br>Social worker in MDT |

**Key:** CC - community care, CARE - Complex And Restorative, CGA – comprehensive geriatric assessment, H - hospital care (secondary / tertiary care), MDT - multidisciplinary team, PC - primary care, SC - social care, TC - tertiary care

##### Multidisciplinary care for older people with dementia and complex chronic geriatric conditions

Two studies focusing on **dementia** (Lee et al. 2017, Luckett et al. 2017) and one on **complex chronic geriatric conditions** (Lee et al. 2015) reported healthcare professionals’ experiences in relation to two unique integrated care interventions. Out of these two unique interventions, one was a **multidisciplinary collaborative memory clinic model** and its expansion to complex chronic geriatric diseases. These multidisciplinary collaborative memory clinics integrated care across primary, hospital (secondary / tertiary care), community, and social care by creating predetermined pathways, enabling MDT working, and coordinating care across different services (Lee et al. 2015, Lee et al. 2017). Accredited comprehensive training was also provided for healthcare professionals enabling primary care professionals to enhance their roles (Lee et al. 2015, Lee et al. 2017).

The other integrated care intervention that focused on older people with dementia was facilitated **family case conferencing** for nursing home residents with advanced dementia, organised by Palliative Care Planning Coordinators (PCPCs) (Luckett et al. 2017). Care planning coordinators were also responsible for developing care plans, and trained staff in person-centred palliative care (Luckett et al. 2017). The model emphasised a MDT approach where PCPCs worked alongside other professions from primary and community care services to improve communication and care planning (Luckett et al. 2017).

##### Integration of primary and community care for older people with aged care needs

One integrated care intervention focused on co-location of six previously disparate primary and community care services (Chow et al. 2015). These services were all relocated to the same premises to create the Triple I (Hub) where multiple referrals to different service providers could be made through a single pathway (Chow et al. 2015). Intervention providers included primary care practitioners, case managers and community nurses.

Processes were formalised so that case managers worked collaboratively with primary care practitioners to integrate care, facilitate assessment and care planning, provide individualised information and to ensure general practitioner engagement during care transitions (Chow et al. 2015).

##### Multidisciplinary hospital based geriatric care for older people with aging associated diseases

One study exploring care for **aging associated diseases** reported one unique intervention (Aberg & Ehrenberg 2017). The intervention was provided across hospital and social care by enabling MDT working in a geriatric clinic located across four wards. The geriatric clinic also coordinated care based on joint assessment by the MDT which included the provision of CGA.

##### Multidisciplinary hip fracture care pathway for older people

One integrated care intervention was a hip fracture care pathway (Fox et al. 2023). The pathway involved multiple hospital departments (three urban and one rural), a wide range of professionals and spanned hospital and community care by ensuring prompt communication via integrated patient records. The pathway started with patient admission to an acute care hip fracture ward, rapid optimisation of fitness for surgery and time-specific targets for surgery. Guidelines advocate coordinated orthogeriatric and multi-disciplinary review enabling successful discharge to community care services (Fox et al. 2023).

##### Emergency department avoidance service for older people with urgent but non-emergency care needs

One study focusing older people requiring **urgent but non-emergency care** reported patients’ and relatives’ experiences in relation to one unique intervention (Greene et al. 2023). The Complex And RestorativE (CARE) centre, an ED avoidance service, provided care across hospital and social care by bringing health and social care professionals together in a MDT at a new unit. The CARE centre emphasised rapid assessment and treatment without overnight stays (Greene et al. 2023).

#### 2.3.4 Waiting time categories

The included studies reported a variety of findings related to the impact of the integrated care interventions upon access to healthcare and waiting times. These were categorised into routine care outpatient diagnostic waiting times (Lee et al. 2017), elective and routine care treatment waiting times (Chow et al. 2015, Lee et al. 2015), emergency waiting times in hospital (Greene et al. 2023), inpatient waiting times for surgery or treatment (Aberg & Ehrenberg 2017, Fox et al. 2023) and other types of waiting times (Luckett et al. 2017).

### 2.4 Experiences of waiting time in the context of integrated care

The qualitative component of this review aims to explore older people’s experiences of waiting time in the context of integrated care. In this section, the experiences of healthcare professionals, older people and their relatives are reported, with findings grouped according to population, type of intervention, and waiting time category.

#### 2.4.1 Healthcare professionals’ experiences of multidisciplinary care for older people with dementia and complex chronic geriatric conditions

In this section the experiences of healthcare professionals of two integrated care interventions for older people with dementia and complex chronic geriatric conditions are reported. The integrated care interventions were a multidisciplinary collaborative memory clinic model and its expansion to complex chronic geriatric diseases (Lee et al. 2015, Lee et al. 2017) and family case conferencing in nursing homes organised by a PCPC (Luckett et al. 2017). The waiting time experiences reported by healthcare professionals were categorised into three groups: routine care outpatient diagnostic waiting times, elective and routine care treatment waiting times, other types of waiting times. The waiting time experiences are reported by category.

##### Routine care outpatient diagnostic waiting times

The study by Lee et al. (2017) reflects upon multidisciplinary collaborative memory clinics in Canada and their significance in the care of older adults with dementia, with one particular focus on improving access to care. Two subthemes were identified from open ended responses to a survey: “timely access to assessment” and “early access to detection and intervention”.

The findings related to timely access showed several service improvements, specifically in relation to the collaboration of healthcare professionals within the integrated team, such as being able to be assessed. Healthcare professionals felt that the integrated team approach reduced waiting times, allowing patients to be assessed without the need to wait for a geriatrician and offering much quicker access compared to traditional specialist referrals, with assessments typically completed within 1–2 months.

> *“Access without having to wait and see the geriatrician”* (Lee et al. 2017, p.60)

> *“Much improved access to the team over traditional long wait referrals to specialists”* (Lee et al. 2017, p.60)

> *“Able to have assessment in timely fashion -zseen within 1-2 months (as compared to specialist).”* (Lee et al. 2017, p.60)

Early access was also identified in this study, specifically related to the detection of dementia and early access to health interventions. Healthcare professionals noted that cognitive deficits were identified earlier and patients were started on cognitive enhancers as soon as dementia was diagnosed.

> *“Earlier identification of cognitive deficits through use of our memory clinic”* (Lee et al. 2017, p.60).

> *“Early pick up of dementia – patients are started on cognitive enhancers as soon as dementia identified”* (Lee et al. 2017, p.60).

The impact of this early detection and the receiving appropriate and prompt treatment was captured by one healthcare professional. They observed that some patients were assessed early for reversible causes of cognitive decline and, after implementing recommended changes, subsequently tested within the normal range.

> *“Assessed some patients early with reversible causes; once they implemented recommended changes, tested within the normal range.”* (Lee et al. 2017, p.60).

The value of the accredited comprehensive training offered by the model was further recognised with one participant noting that the clinic promptly addressed memory concerns by ensuring assessments were carried out by individuals with appropriate training.

> *“Addressing memory concerns in a timelier manner with an individual who has training to properly assess.”* (Lee et al. 2017, p.60).

##### Elective and routine care treatment waiting times

The study by Lee et al. (2015) highlighted the potential benefits of the multidisciplinary collaborative memory clinic model in relation to elective and routine care treatment waiting times. One healthcare professional suggested that expanding the memory clinic model to complex chronic geriatric diseases could help reduce waiting times, decrease the demand for acute care, and improve cost-effectiveness, provided there was adequate remuneration for physicians and sufficient funding for training and staffing.

> *“I think that expanding the memory clinic would reduce wait times, reduce acute care, and be cost effective if there was adequate remuneration for physicians and funding for appropriate training and staffing.”* (Lee et al. 2015, p.153).

##### Other types of waiting times

The study by Luckett et al. (2017) highlighted the benefits of facilitated case conferencing in improving communication and care planning for nursing home residents with advanced dementia living in Australia. One of the most commonly perceived benefits was the enhancement of family–nursing home communication. Case conferences allowed the team to address care planning earlier in the disease trajectory. This gave family members time to absorb information, reflect on their loved one’s wishes, and engage in discussions between meetings, thereby increasing their involvement in decision-making.

> *“Families have been involved in decisions earlier than previously. This has enabled a softer approach to advance care planning.”* - Assistant in nursing (Luckett et al. 2017, p.1716).

The role of PCPCs was seen as crucial in this process. The dedicated time allocated for coordinating case conferences enabled a concerted effort to engage physicians and other medical specialists. Healthcare professionals felt that this involvement allowed symptoms and medical concerns to be addressed more promptly.

> *“We’ve got the GPs involved and that’s helped because … the pain gets addressed straight away.”* - PCPC (Luckett et al. 2017, p.1717).

#### 2.4.2 Healthcare professionals’ experiences of integration of primary and community care for older people with aged care needs

In this section the experiences of healthcare professionals with one integrated care intervention for older people with age care needs are reported. The integrated care intervention was the Triple I (Hub) aiming to co-locate and coordinate primary and community services (Chow et al. 2015). The waiting time experiences reported by healthcare professionals were categorised into one group: elective and routine care treatment waiting times.

##### Elective and routine care treatment waiting times

One of the key aspects of the Triple I (Hub), located in Australia, was the impact on waiting times for aged care referrals. The findings revealed substantial reduction in processing time of referrals.

> *“significant reduction in processing time of aged care referrals from three weeks to less than 24 hours.”* (Chow et al. 2015, p.235).

The study did acknowledge that the initial implementation did face some challenges related to role clarity and task allocation as specific tasks were not assigned to all staff members and as a result improvements were suggested, such as the development of clear procedure manuals. Establishing clearer guidelines was identified as a crucial step towards achieving greater efficiency. By providing staff with well-defined roles and responsibilities the hub aimed to enhance coordination and reduce any potential confusion or overlap in duties.

> *“Responsibilities were accepted by staff members, rather than assigned by management at the start of the process. Clear position descriptions, policy documents and detailed procedures manuals including role allocations for all staff to refer to were identified as a necessary step towards greater efficiency in the newly amalgamated service group.”* (Chow et al. 2015, p.235).

Additionally, the absence of purpose-designed intake forms at the implementation stage was recognised as a barrier to optimal efficiency. This practical step was seen as important for improving the intake process, ensuring that any relevant information was captured accurately and efficiently.

> *“Purpose designed intake forms were not designed for the Triple I (Hub) at the implementation stage. Efficiency could be increased by taking the practical steps such as designing a form for referrers to complete, and a screening questionnaire for patients, which would contain information that could be entered into the system by administrative, rather than clinical staff who would be available for triaging cases where clinical need was indicated by the referrer on completion of the questionnaire.”* (Chow et al. 2025, p.235).

#### 2.4.3 Healthcare professionals’ experiences of multidisciplinary hospital based geriatric care for aging associated diseases

This section covers the experiences of healthcare professionals with one integrated care intervention for older people with aging associated diseases (Aberg & Ehrenberg 2017). The integrated care intervention was multidisciplinary hospital based geriatric care that aimed to provide coordination of hospital and social services based on CGA. The waiting time experiences reported by healthcare professionals were categorised into one group: inpatient waiting times for surgery or treatment (Aberg & Ehrenberg 2017).

##### Inpatient waiting times for surgery or treatment

The findings within the study by Aberg & Ehrenberg (2017) reported findings related to inpatient waiting times. Healthcare professionals working in the multidisciplinary geriatric clinic in Sweden recognised the value of early assessment following hospital admission with aging associated diseases.

> *“The goal is for the team to meet the patient on the day they are admitted so they can do at least an initial assessment, and also so they can initiate contact with the municipality, as well as the other relevant care operators – that this is part of it from the start.”* (Aberg & Ehrenberg 2017, p.116).

Early assessment and contact with patients and relatives prior to hospitalisation was also reported as crucial:

> *“The more and the earlier we can inform the patient and inform the relatives, the calmer things get, the easier it is to work undisturbed and the more effective we can be…”* (Aberg & Ehrenberg 2017, p.116).

This early involvement helped to map out expectations and potential problems:

> *“You notice a tremendous difference in how things progress when you take it at the start, straight away, and you can see what the expectations are and what problems might exist…”* (Aberg & Ehrenberg 2017, p.116).

#### 2.4.4 Healthcare professionals’ experiences of multidisciplinary hip fracture care pathway for older people

In this section the experiences of healthcare professionals with one integrated care intervention for older people with hip fracture are presented (Fox et al. 2023). The integrated care intervention was multidisciplinary hip fracture care pathway that aimed to provide rapid optimisation of fitness for surgery and time-specific care targets. The waiting time experiences reported by healthcare professionals were categorised as inpatient waiting times for surgery or treatment (Fox et al. 2023).

##### Inpatient waiting times for surgery or treatment

The study by Fox et al. 2023 explored the organisational factors influencing multidisciplinary hip fracture care pathways across four UK hospitals. Differences between hospitals was identified, particularly the lack of clear guidelines in relation to admissions and related procedures, which could cause delays. One healthcare professional reported that the lack of guidance and designated equipment led to time being wasted in the Accident and Emergency (A&E) department, often resulting in patients not receiving timely anaesthesia.

This issue was resolved by developing a designated trolly with all necessary equipment for delivering the fascia iliaca nerve blocks, training a range of staff to deliver them, and ensuring reliable access to equipment.

> *“Our fascia iliaca block trolley is good. We were having problems where our equipment was all over the place and we were taking an awful lot of time to find the stuff to do the blocks to the extent that people just couldn’t be bothered. We’re incredibly busy in A&E and the amount of time people were finding trying to get the stuff was an issue. We found that we’d got a trolley that locks and has everything you would need in it. You bring the trolley to the patient to give the block … a number of nurse practitioners have competency packs now”* (Lucy, an ED Consultant at Springhill) (Fox et al. 2023, p.4).

Joint assessments by physiotherapists and occupational therapists, along with maintaining shared plans for patients’ rehabilitation and discharge were found to be most efficient for hospital care.

> *“Now, we tend to do a lot more joint assessments and joint working and actually, I think it’s better for the patient because it’s all much more coordinated. You’re getting two separate viewpoints but at the same time and then it’s easier to come up with those discharge options and which route is going to be more suitable, or what to try next because it’s much more coordinated. I think communication is improved a lot, again, over the past few years to make it work better”* (Chloe, Specialist Occupational Therapist at Maplegrove) (Fox et al. 2023, p.4)

A barrier to timely care was identified as potential lack of communication and awareness around practice targets and guidelines. A junior doctor highlighted that they only learnt about the 48-hour surgical targets for hip fracture by accident.

> *“PPT:” So, I don’t actually know what the targets are, well I was never told them or I was never explained them or anything. I only knew this because I saw one of my colleagues doing an audit, the national hip fracture audit or putting them on the national database, and I was like, ‘oh what’s that you’re doing’? And then she said, ‘oh yes, we have to operate on them within 48 h if we can, and that’s the national target’. So, I don’t think people are aware unless there’s some kind of hearsay or they figure it out. But I think that is a good thing to know if possible …*

> *IV: why would it be particularly helpful for you to know about the expected targets? PPT: “So, obviously we’re only there for four month and you could spend a month and a half doing it wrong before you do it right and then you’ve only got two and a half months left trying to do it right. Or for example even as a junior it’s important for us to understand why it’s, from an education point of view, why it’s important to operate on a patient within 48 h, are there risks to not operating on them, what are the complications. Just as an education stand, is it a national target because of money or is it a national target because of patient care or is it a national target because of bed flow in the hospitals like, is there a reason behind this? So in that sense its quite important as well”* (Alice, F2 Doctor at Maplegrove) (Fox et al. 2023, p.5)

The embedded use of the hip fracture pathway documentation and regular performance monitoring motivated the multi-disciplinary team to constantly reflect on each patients’ progress.

> *“a lot of the drive comes from the hip fracture pathway. That you’re on a bit more of a schedule. Whereas if you haven’t got a pathway, everything’s a bit wishy-washy. But I think the hip fracture pathway does motivate people. Because there is a constant pressure. Because it’s, ‘okay, it’s seven days after their operation, what’s happening? Why aren’t we progressing’? Because we’re an MDT team, you’re not left alone, there’s a constant drive from every member or profession of the MDT to progress that patient … So I think everyone has a bit of a collective drive, but I think that being on that ward and on the pathway is a mega drive. Because it’s constantly evaluated”* (Jane, Occupational Therapist, Springhill) (Fox et al. 2023 p.5)

It was recognised that regular performance monitoring could help to identify delays and participants suggested that an investigation should be automatically triggered when targets were not met.

> *“we are always looking at our figures, we are always downloading the NHFD data to see, you know, are there any trends … and then between us we will look at it and go, ‘ooh you know we are getting a few delays here due to DOACs [direct oral anticoagulants]’, that kind of thing, so then we will just remind the team of the DOAC guidelines and so we are very proactive instead of reactive […]”* (David, Orthogeriatric Advanced Nurse Specialist, Springhill) (Fox et al, 2023 p.5)

#### 2.4.5 Patients’ and relatives’ experiences of an emergency department avoidance service

In this section the experiences of patients and relatives with one integrated care intervention for older people with urgent but non-emergency care needs (Greene et al. 2023). The integrated care interventions was CARE centre, an ED avoidance service that emphasised MDT working, rapid assessment and treatment without overnight stays (Greene et al. 2023). The waiting time experiences reported were categorised into one group: emergency waiting times.

##### Emergency waiting times

The study by Greene et al. (2023) highlighted the impact of the CARE Centre in Australia on emergency waiting times in hospital for older people with urgent non-emergency care needs. Patients reported feeling positive about being taken to the CARE Centre and did not express any reluctance when offered the opportunity compared with traditional EDs. One relative also expressed feelings of relief at the prospect of their family member being seen quicker.

> *“So, when he said you’ll get looked at very quickly at the other place, absolutely we’ll go there”.* (Relative, 2001) (Greene et al. 2023, p. 643)

All participants favoured the CARE Centre over traditional EDs, with many mentioning a fear of ramping, which was described as prolonged waits in an ambulance due to overcrowding.

> *“We would have been there two, three, four, five – the paramedic was saying the day before they were ramping in the ambulance for eight or 9 hours…Then you get stuck in there and it’s stuck in a corridor and everyone’s walking past, and no one’s really taking care of you. Then you start to get stressed that you’re going to get locked in there overnight. It’s not a good place, so the CARE centre was just paradise”* (Relative, 2001) (Greene et al. 2023, p.643)

Participants valued that the CARE Centre was a day service only and that issues were resolved promptly.

> *“In comparison to when I’ve had falls and gone to (the usual ED), I know they’re very busy there and of course I’ve had to wait and stay the night. Here it was all resolved in that day”* (Patient, 1081) (Greene et al. 2023, p. 643)

#### 2.4.6 Bottom line summary

The identified evidence was conducted in four different countries, three studies were from Australia, two from Canada, one from Sweden and one from the United Kingdom. The evidence covered a range of populations, including older people living with dementia (two studies), complex chronic geriatric diseases (one study), aged care needs (one study), aging associated disease (one study), hip fracture (one study), and urgent but non-emergency care needs (one study). Integration between primary care services and other providers was the most commonly reported (four studies).

The studies explored healthcare professionals’, patients’ and relatives’ experiences of a wide range of interventions including multidisciplinary collaborative memory clinic models, case conferencing, multidisciplinary hospital based geriatric care, multidisciplinary hip fracture pathways, and ED avoidance service. Waiting time experiences could be categorised as inpatient, routine care, emergency and other waiting times and across all the studies the participants mainly reported positive experiences in relation to the timeliness of care provision. Findings related to waiting times are summarised below for each study.

- Healthcare professionals in Canada suggested that the multidisciplinary collaborative memory care clinic may support early assessment and diagnosis of dementia and complex chronic geriatric conditions.
- Healthcare professionals in Australia suggested that case conferencing could support earlier dementia care planning, strengthen family involvement, and enable more timely symptom management through greater physician engagement in nursing home care.
- Healthcare professionals in Australia suggested that the processing time of aged care referrals substantially reduced following integration of primary and community care for older people with aged care needs.
- Healthcare professionals in a multidisciplinary geriatric clinic in Sweden suggested that conducting early assessments may help streamline inpatient care, improve communication with families, and address potential issues from the outset.
- Healthcare professionals in the UK suggested that clear guidelines, joint assessment and regular performance monitoring as part of a multidisciplinary hip fracture pathway can lead to more efficient patient care and reduction of delays, although communication of certain care targets were identified as barriers.
- Patients and relatives in Australia suggested that the CARE Centre, an ED avoidance service for older adults with urgent but non-emergency needs, may help reduce emergency waiting times.

However, the findings highlight a distinct lack of qualitative evidence on experiences of waiting times in relation to integrated care. This was particularly evident from the perspectives of patients and older people living with health conditions.

### 2.5 Summary of integrated care interventions within hospital settings

This section summarises the studies that reported interventions providing integration within hospital settings. These interventions did not report collaboration or coordination with other service providers, such as primary, community or social care. The section starts with a summary of study designs, country of origin, integrated care interventions and population focus. Finally, a summary of waiting time outcomes is reported.

#### 2.5.1 Overview of studies reporting integrated care interventions within hospital settings

All of the 31 studies utilised a quantitative study design. The studies were conducted across 10 countries:

- USA (nine studies) (Ackermann et al. 2023, Burton et al. 2020, Cieremans et al. 2023, Godin et al. 2015, Hansen et al. 2020, Jackson et al. 2019, Morris et al. 2020, Steffensmeier et al. 2022, VanTienderen et al. 2021)
- Netherlands (five studies) (de Gans et al. 2023, Folbert et al. 2017, Nijmeijer et al. 2018, Schuijt et al. 2020, van Voorden et al. 2020)
- Italy (five studies) (Aletto et al. 2020, Bano et al. 2020, Baroni et al. 2019, Quaranta et al. 2021, Rostagno et al. 2016)
- Australia (four studies) (Goh et al. 2016, Ling et al. 2015, Lynch et al. 2015, Talevski et al. 2020)
- UK (two studies) (Middleton et al. 2017, Mubark et al. 2020)
- Germany (two studies) (Hafner et al. 2021, Werner et al. 2020)
- Denmark (one study) (Kristensen et al. 2016)
- Ireland (one study) (Murphy et al. 2019)
- Norway (one study) (Solberg et al. 2023)
- Spain (one study) (Pablos-Hernandez et al. 2020)

Twenty-eight studies focused on integrated care interventions for **hip fracture and other orthopaedic trauma** patients, all of which examined different models of multidisciplinary care. One study focused on interprofessional and intraprofessional clinical collaboration in multimorbid older patients (de Gans et al. 2023). One study reported on the integration of a medical resident into an aged psychiatry inpatient unit to address physical health issues of older patients in a more timely manner (Goh et al. 2016). Finally, one study investigated the effectiveness of multidisciplinary care in the treatment of head and neck squamous cell carcinoma in older patients (Hansen et al. 2020).

#### 2.5.2 Waiting time outcomes

The included studies reported a range of different waiting time outcomes which can be categorised into inpatient and emergency settings. **Inpatient waiting times** were the most frequently reported (27 studies), and included the following: time to rehabilitation (days) (Murphy et al. 2019), time to initial assessment (hours) (Goh et al. 2016, Hafner et al. 2021, Middleton et al. 2017); time from admission to clinical evaluation (days) and percentage of patients undergoing early clinical evaluation (<24 hours) (Rostagno et al. 2016); time to medical readiness (hours) (Steffensmeier et al. 2022); time to radiological procedure (De Gans et al. 2023); time from diagnosis to treatment (surgical resection, chemotherapy or radiotherapy) and time from surgical resection to chemotherapy/radiotherapy (Hansen et al. 2020); and time to surgery. Time to surgery was reported across 27 studies and was usually calculated from the time of admission to the hospital (after the decision for surgery is made) until the start of the surgical procedure (25 studies). In four studies time to surgery was also defined as the period from the point of arrival at the emergency department (ED) to the beginning of the procedure (Ackermann et al. 2023, Cieremans et al. 2023, Jackson et al. 2019, van Voorden et al. 2020). Time to surgery was usually measured in hours (17 studies) or days (4 studies), but in 13 studies percentage of patients undergoing surgery within specific timeframes (within 12, 24, 36 or 48 hours) was also calculated.

**Emergency waiting times** at a hospital were reported in five studies. Emergency department stay captured the time spent in the ED (Lynch et al. 2015, Middleton et al. 2017, Nijmeijer et al. 2018, Schuijt et al. 2020), while other measures included time from ED arrival to cardiology consultation (Ackerman et al, 2023).

#### 2.5.3 Summary of integrated care interventions within hospital settings

The identified evidence was conducted in 10 different countries, with the USA contributing the highest number of research studies (nine studies). The studies reported care integration within hospital settings (secondary/tertiary care) by forming MDTs (31 studies). Most (27 studies) reported on inpatient waiting times, such as time to surgery (27 studies). The evidence consists of quantitative studies with majority focusing on hip fracture and other orthopaedic trauma (28 studies). While some evidence also focused on emergency waiting times (five studies), the number of studies were much lower.

## 3. DISCUSSION

### 3.1 Summary of the findings

This rapid review incorporated both quantitative and qualitative evidence to evaluate the impact of integrated care interventions on waiting times and waiting lists for older adults and people living with frailty. A total of 61 studies exploring different models of integrated care were included. Of these, 30 studies focused on interventions operating across two or more services, while the remaining 31 examined integrated care implemented solely within the hospital (secondary or tertiary care) setting. Due to the breadth of included studies and the constraints of the rapid review process, the final synthesis focused on the 30 studies operating across two or more services.

Identified studies were mainly of quantitative research design (n=23), addressing the first aim of this rapid review focusing on the effectiveness of integrated care in reducing waiting times. These studies covered diverse integrated care interventions for older people with various health conditions and reported a range of waiting time outcomes. While most integrated care interventions included in this review indicates some improvement in waiting time outcomes, such as time to surgery, ED stay and others, the overall body of evidence is mostly considered weak, due to low quality research and the lack of robust study designs. Additionally, with regards to other types of waiting times or lists, such as routine care and elective waiting times, only a few studies measured these (4 quantitative studies), limiting any conclusion to be drawn.

The other aim of this rapid review was to explore the views of healthcare professionals and older people or individuals living with frailty regarding waiting times in the context of integrated care. Seven qualitative studies were identified, mainly exploring the experiences of healthcare professionals (6 studies) across a range of integrated care interventions and medical specialties. While all qualitative research reported that integrated care interventions helped reduce waiting times and improved timely care provision, the lack of patient perspective makes the findings less confirmable and transferable to the wider population.

### 3.2 Comparison with the wider literature

The findings of this rapid review align with the wider literature that also found that inconsistent evidence existed regarding the effectiveness of integrated care interventions in reducing waiting times (Van Heghe et al. 2022, Baxter et al. 2018b). Baxter et al. (2018b) identified a limited number of UK and international literature (nine research studies and two reviews), and found that while some integrated care interventions reduced waiting times, others did not improve or even increased the time patients spent waiting. However, Baxter et al. (2018b) focused on a wide range of different populations including children and older adults, a variety of conditions from gynaecological issues to diabetes, and different interventions which could have led to the inconsistency in the findings. Another review by Van Hedge et al. (2022) focused on multidisciplinary orthogeriatric care for hip fracture patients and found that integrated care models did not statistically significantly change time to surgery, although the findings indicated a small reduction. Additionally, the studies reported in the review of Van Hedge et al. (2022) had moderate to high risk of bias (medium to low quality) and were all observational studies, potentially contributing to the inconclusive results. This is similar to the findings of this rapid review, as the majority of multidisciplinary orthogeriatric care interventions for older people with hip and other upper and lower extremity fractures were of moderate or low quality with the results indicating inconsistency.

While most integrated care interventions identified in this rapid review had similar elements (MDT, pathways / protocols, care coordination, and others), differences in how these elements were utilised, what medical specialties were involved, and the number of services integrated (two or more) makes it difficult to draw an overall conclusion regarding their effectiveness. This issue was also highlighted in Baxter et al. (2018b), who concluded that future research needs to focus on the link between particular integrated care intervention elements (MDT, pathways / protocols, care coordination, and others) and change in outcomes to enable the identification of what works.

This rapid review identified multiple different waiting times, with the majority of research focusing on inpatient waiting times (16 out of 23 quantitative studies), and smaller subset of studies reporting emergency (6 studies) and routine care waiting times (4 studies). This indicates that limited research is focusing on routine and elective waiting times. This is supported by the wider literature. Baxter et al. (2018b) also identified a limited number of quantitative studies (7) that focused on emergency (2) or routine care waiting times (2).

Reed et al. (2021) undertook a review of UK integrated care initiatives and found that waiting times for clinical assessment data was not available as an outcome. These are surprising findings, considering that increasing waiting times, particularly for emergency and elective care is a growing issue in the UK and worldwide (Welsh Government 2024b, Welsh Government 2024c, Welsh Parliament 2022, OECD 2020). However, evidence suggests that measuring the effectiveness of integrated care interventions is notoriously difficult, which could also explain the lack of evidence focusing on routine and elective care waiting times.

Keeble (2019) identified areas that make outcome measurement difficult in integrated care, including lack of data availability across organisations, changes in service data collection over time, and finding a true control group. Kelly et al. (2020) found multiple challenges in measuring integrated care, which included a lack of robust measurement tools, and the infrequent use of common outcome measures. This aligns with the findings of this rapid review, as while waiting time outcomes could be categorised as inpatient, emergency, and routine care, almost all of them covered different time periods related to specific assessment and procedures, such as time to geriatric assessment, or time to physical therapy.

Additionally, some waiting times were captured as the proportion of patients seen during a certain period (48 hours) or relied on healthcare professionals self-report (Groenewoud et al. 2021), instead of time being measured. This highlights the need for common waiting time measurement across studies.

### 3.3 Strengths and limitations of the available evidence

All of the evidence came from academic papers with no grey literature reports included or appraised. The majority of grey literature reports retrieved lacked detailed methodologies, raising concerns about the certainty and reliability of any conclusions drawn. Among those that did include methods, most focused on integrated care in general rather than evaluating specific interventions or did not report waiting times as an outcome. This highlights a significant gap in the grey literature, both in terms of relevance to the question and the quality required to draw meaningful conclusions. Some examples of good practice case studies were identified, although these lacked sufficient methodological detail for critical evaluation within the rapid review (Bevan Commission 2025).

The available quantitative studies cover a range of health conditions, although most focused on hip fractures. The studies reported various outcomes related to waiting times, but many considered waiting times as a secondary outcome rather than the primary focus. Most of the waiting time data originated from inpatient settings, with a few studies reporting from emergency settings or routine care. Notably, a limited number of studies were identified that demonstrated the impact of integrated services on routine and elective waiting times, which aligns with wider evidence suggesting that waiting time is not a commonly measured outcome (Reed et al. 2021). However, it must be noted that evidence investigating the effectiveness of integrated care interventions on other outcomes, such as patient satisfaction, quality of care, hospital admission, length of stay, does exist (Baxter et al. 2018b, Damery et al. 2016), but was not covered by this review.

The majority of quantitative studies were of a weak design, limiting the robustness of the included evidence. Two randomised controlled trials were evaluated as strong, providing more reliable insights. The disparity in study quality highlights a limitation in the available evidence, affecting the overall strength and reliability of the review’s conclusions. More rigorous research is needed to provide clearer and more definitive evidence of the effect of integrated care on waiting times across all contexts covered by this review.

There was also a lack of qualitative research focusing on people’s experiences of waiting times in relation to integrated care, although qualitative evidence exploring integrated care more broadly does exist (Lawless et al. 2020, Smith et al. 2021). Most qualitative studies conducted interviews with healthcare professionals with only one study focusing on patient experiences.

### 3.4 Strengths and limitations of this Rapid Review

The strength of this rapid review is that comprehensive systematic search methods were employed, which included searching five bibliographic databases, a complementary search of two clinical trial registers and grey literature sources. Identified systematic reviews and scoping reviews were also checked for additional studies that met the inclusion criteria. This enabled the identification of all relevant studies. All studies were screened for relevance, and full-text screening was performed by two reviewers to ensure accuracy. Selected studies were critically appraised and included regardless of their methodological quality to provide a full account of the state of the literature on the topic. However, methodological limitations were considered when reporting the results. A unified critical appraisal tool was used for all quantitative studies, which allowed the comparison of methodological quality across the different studies while accounting for study design during the appraisal process.

This work also has a number of limitations arising from the time constraints associated with a rapid review. Firstly, it was not possible to provide an in-depth analysis of all included studies within the available timeframe. Therefore, only a subset of studies (n=30) that focused on integrated care across two or more services underwent critical appraisal and data extraction. The remaining studies (n=31) were described but not evaluated in detail, meaning that critical analysis of integrated care interventions within hospital settings could change the overall assessment of the evidence, particularly regarding time to surgery in hip fracture.

Secondly, the bibliographic database searches were restricted by adding search terms specific to waiting time/list outcomes. Due to the large volume of existing evidence on integrated care that is not specific to waiting times (Baxter et al. 2018b, Damery et al. 2016), it was not possible to search for integrated care interventions more broadly (without these restrictions). A broad search would have retrieved too many hits and it would not have been possible to scan all of them within the time available. Adding waiting time/list terms to the searches helped make the rapid review manageable. However, this could also mean that studies that reported waiting time outcomes, but not explicitly mentioned this in the title or the abstract, may have been missed. However, a wide range of terms related to waiting time/lists were included in the searches to ensure that relevant studies were identified.

Additionally, complimentary searches of clinical trial registers, Google, and checking the list of studies included in existing systematic and scoping reviews enabled the identification of reports not found via bibliographic databases. Another potential limitation is that due to the large volume of studies identified and the time constraints associated with a rapid review, citation searching was not performed.

### 3.5 Implications for policy and practice

- There is some evidence that multidisciplinary working, development of care pathways and protocols, and care coordination may improve inpatient waiting times to surgery, and emergency waiting times in an ED. Thus, initiatives supporting the development and implementation of these integrated care interventions is crucial.
- While the majority of the evidence on the effectiveness of integrated care in reducing waiting times was rated weak, this should not be interpreted as a lack of effect. Instead, this highlights an important gap in the literature and the need for more high quality research specifically on integrated care and its impact on waiting times before any firm conclusions can be made. However, it is also noted that the evidence suggests improvements in waiting times alongside better patient experience, and there was no evidence indicating that integrated care could make waiting times worse.

### 3.6 Implications for future research

- There is a need for high quality studies investigating the effect of integrated care on waiting times, particularly on routine care and elective waiting times.
- Majority of the available research studies seem to focus on multidisciplinary team working and the development of pathways and protocols as a form of integrated care and its impact on waiting times. However, there seems to be less focus on organisational integration, such as coordination of governance systems across providers, development of contractual arrangements across different services or joint commissioning (Reed et al. 2021). More research with rigorous study designs is necessary in this topic to evaluate the effectiveness of organisational integration on waiting times.
- There is a need for high quality qualitative research that explores people’s experiences with waiting times in relation to integrated care, particularly from older and frail people’s perspectives.

### 3.7 Economic considerations*

- The relationship between inpatient waiting times and total hospital costs has been estimated to exhibit an initial linear component that is negative, while the (overall) quadratic is positive (U shaped) (Siciliani et al. 2009). Suggesting that increasing waiting times up to a certain level decreases total hospital costs, but past this level, the effect is reversed and total hospital costs increase with length of inpatient waiting time. This point of inflection also suggests there is an optimal period of wait that minimises total costs (Siciliani et al. 2009). Inpatient waiting times of less than 10 days (categorised as the days between the decision of being admitted to the waiting list and the actual admission for treatment, across all hospital treatment functions) minimise total hospital costs (Siciliani et al. 2009). Initiatives reducing time spent waiting to near or below this minimal expenditure length may bring positive economic benefit to the NHS through reduced hospital costs.
- Reductions in total elective waiting lists/times in the UK may generate significant economic benefits. An estimated £73 billion in total benefits may be generated between 2023 and 2027 if the NHS meets its elective waiting list reduction targets (Williamson & Patel 2023). These benefits are mostly made up of contributions to the informal economy (including familial childcare and caring for sick or elderly relatives). Reducing the waiting lists to target may save £14 billion in expenditure by government and households through lower spending on health and social care, and informal care services (Williamson & Patel 2023).

## Data Availability

All data produced in the present study are available upon reasonable request to the authors

## Abbreviations

Acronym: Full Description
A&E: Accident and Emergency
CARE: Complex And RestorativE
CGA: Comprehensive geriatric assessment
CI: Confidence interval
COPE: Comprehensive older person’s evaluation
GP: General Practitioner
ED: Emergency department
MDT: Multidisciplinary team
PCPCs: Palliative care planning co-ordinators
RCT: Randomised controlled trial

## Glossary

**Agreed referral criteria:** Agreed referral criteria outline the predetermined conditions under which referral or transfer from one service to another is initiated (Baxter et al. 2018b).

**Care coordination:** Care coordination describes the process of care organisation that is usually performed by a named point of contact who aims to bring together different healthcare providers and specialists to support the patient (Skills for Care 2018, Baxter et al. 2018b). Care coordination may also include the assessment and regular monitoring of the patient and care delivery (Skills for Care 2018, Baxter et al. 2018b).

**Cohort study:** “An observational study in which a defined group of persons is followed or traced over a period of time. The outcomes are compared between exposed and non- exposed subjects (or between subjects exposed at different levels) to a particular intervention or other factor of interest. A prospective cohort study assembles participants and follows them into the future. A retrospective cohort study identifies subjects from past records and follows them from a pre-specified starting point to the present or to the end of a pre-specified data collection period.” (Public Health Agency of Canada 2014, p. 70)

**Comprehensive geriatric assessment (CGA):** Comprehensive geriatric assessment can be defined as a multidimensional holistic assessment that considers older people’s concerns with the aim to develop a plan that can help meet their needs (British Geriatric Society 2019).

**Controlled before and after study:** “There is no random or quasi-random assignment to group. In general, participants are assigned as part of a natural grouping, e.g., they work together in the same geographic area.” “There is also a period of baseline assessment, rather than baseline assessment occurring at a single point in time.” (Public Health Agency of Canada 2014, p. 15)

**Extrapolation:** A concept used for rating the overall body of evidence for quantitative studies. It is defined by Public Health Agency of Canada (2014) as “inference drawn from studies that researched a different but related key question” (p. 6). In this rapid review, outcomes that did not directly measure time, such as percentage of patients receiving surgery within 48 hours or GP perceived quality of care, were considered extrapolations.

**Frailty:** “Frailty is a long-term condition. It describes a state of health whereby body systems gradually lose their biological, physical, and mental resilience. […] In simple terms, frailty affects the person’s ability to cope with even minor illness, infection, or stressful life events such as a change in living circumstances, or bereavement (particularly of a spouse or partner).” (Welsh Government 2024a)

**Hazard ratio:** A measure of how often an outcome or event occurs in the intervention group compared to how often it happens in the control group, over time (National Cancer Institute 2025).

**Integrated care:** Integrated care can be defined as the joining up of different health and/or social services to deliver care that meets individuals’ needs in an efficient way (Scobie 2021). Various terms can be used to refer to integrated care and these could include: coordinated care, collaborative care, multidisciplinary care, etc. Moreover, multiple interventions could be considered as integrated care, which could include integrated pathways/protocols, staff co-location, multidisciplinary teams, and new units among others (Baxter et al. 2018a, Baxter et al. 2018b).

**Integrated patient records:** Integrated patient records refer to the process of documentation that allows different health and social care providers to share patient information efficiently usually by providing access to the same system (Baxter et al. 2018b).

**Joint assessment:** Joint assessment can be defined as patient examination that is performed by two or more healthcare professionals from different specialisms or disciplines that enables efficient identification of patients’ needs (Baxter et al. 2018b).

**Joint patient review or discharge:** Joint patient review or discharge describes the process of patient evaluation and discharge planning that is performed by two or more healthcare professionals and can reduce duplication, resulting in more efficient care provision (Baxter et al. 2018b).

**Multidisciplinary team (MDT):** Multidisciplinary team refers to a group of healthcare professionals from different disciplines (for example medicine, nursing, physiotherapy, and others) working together to provide specific services for patients (Baxter et al. 2018b).

**New unit / co-location:** New unit or co-location refers to the development of new health and social care departments that brings healthcare professionals from different specialism or discipline under one roof (Baxter et aal. 2018b).

**Non-randomised controlled trial:** “Participants are assigned to being in the intervention or control group in a systematic way that is not truly randomised, e.g., alternating between groups, or using birth years. Baseline assessment occurs at a single point in time.” (Public Health Agency of Canada 2014, p. 15)

**Pathways / protocols:** Pathways or protocols are outlines of care provision that have set timeframes for anticipated procedures and role allocation for different healthcare professionals to ensure patients move progressively through the health and/or social care system (Baxter et al. 2018b).

**Randomised controlled trial (RCT)**: “Participants are randomly assigned to groups by the researcher, e.g., by random number generation or a coin toss. Randomisation allows for better control of unknown confounders.” (Public Health Agency of Canada 2014, p. 15)

**Uncontrolled before and after study:** “There is no concurrent control group. One group of participants received an intervention and results are compared before and after the intervention. The individuals in the post-intervention group may not be the same individuals as in the pre-intervention group.” “The design is considered weak due to inadequacy of the control group.” (Public Health Agency of Canada 2014, p. 14)

**Waiting times/lists:** Waiting time outcomes were conceptualised as any period where patients were waiting for an appointment, diagnosis or treatment, whether this was in the emergency department (ED), inpatient or outpatient (routine) setting. Waiting lists could include number of people on a waiting list or the number of people waiting more than a defined period. Based on a preliminary literature search, a waiting time framework was developed that provides further description of waiting times. Waiting time categories identified were: Routine care/outpatient initial consultation waiting times; Inpatient waiting times for surgery or other treatment; Routine care/outpatient diagnostic waiting times; Elective and routine care treatment waiting times; Emergency waiting times at a hospital; Time to follow-up; Social care waiting times; Other Waiting Times.

**Elective and routine care** waiting times could further be defined as time spent waiting for non-urgent, planned care activities.

## RAPID REVIEW METHODS

### 5.1 Eligibility criteria

The PICO (Population, Intervention, Comparison, Outcome) and PICo (Population, Phenomena of Interest, Context) frameworks were applied to inform the eligibility criteria used to select studies for inclusion in the review.

**Table 5:** Eligibility criteria for review question 1. What is the effectiveness of integrated care in reducing waiting times/lists for older people or individuals living with frailty?

|  | Inclusion criteria | Exclusion criteria |
| --- | --- | --- |
| **Population** | Frail<sup>1</sup>,<br>Older people (Adults over 65 years) | Children and young people<br>Adults under age 65 who are not frail |
| **Intervention** | Integrated care<sup>2</sup> |  |
| **Comparison** | Any |  |
| **Outcome** | Waiting time/lists<sup>3</sup> |  |
| **Study design** | Randomised controlled trials<br>Quasi-experimental studies<br>Observational studies (both controlled and single group)<br>Mixed methods studies | Evidence synthesis (systematic, umbrella, scoping reviews)<br>Narrative reviews<br>Expert opinion |
| **Geographical location** | OECD<sup>4</sup> (peer-reviewed publications)<br>UK (grey literature) | Low and middle income countries |
| **Language of publication** | English | Non-English |
| **Publication date** | 2015 - Current | Primary research studies published prior to 2015 |
| **Publication type** | Peer-reviewed publications, grey literature |  |

**Table 6:** Eligibility criteria for review question 2. What are the experiences of older people or individuals living with frailty regarding waiting times in the context of integrated care?

|  | Inclusion criteria | Exclusion criteria |
| --- | --- | --- |
| **Population** | Frail<sup>1</sup>,<br>Older people (Adults over 65 years),<br>Healthcare professionals | Children and young people<br>Adults under age 65 who are not frail |
| **Phenomena of Interest** | Waiting times/lists<sup>3</sup> |  |
| **Context** | Integrated care<sup>2</sup> |  |
| **Study design** | Qualitative studies<br>Mixed methods studies | Evidence synthesis (systematic, umbrella, scoping reviews)<br>Narrative reviews<br>Expert opinion |
| **Geographical location** | OECD<sup>4</sup> (peer-reviewed publications)<br>UK (grey literature) | Low and middle income countries |
| **Language of publication** | English | Non-English |
| **Publication date** | 2015 - Current | Primary research studies published prior to 2015 |
| **Publication type** | Peer-reviewed publications, grey literature |  |

#### Definitions

1. Frailty: “Frailty is a long-term condition. It describes a state of health whereby body systems gradually lose their biological, physical, and mental resilience. […] In simple terms, frailty affects the person’s ability to cope with even minor illness, infection, or stressful life events such as a change in living circumstances, or bereavement (particularly of a spouse or partner).” (Welsh Government 2024a)
2. Integrated care: Integrated care can be defined as the joining up of different health and/or social services to deliver care that meets individuals’ needs in an efficient way (Scobie 2021). Various terms can be used to refer to integrated care and these could include: coordinated care, collaborative care, multidisciplinary care, etc. Moreover, multiple interventions could be considered as integrated care, which could include integrated pathways/protocols, staff colocation, multidisciplinary teams, and new units among others (Baxter et al. 2018a, Baxter et al. 2018b).
3. Waiting time outcomes were conceptualised as any period where patients were waiting for an appointment, diagnosis or treatment, whether this was in the emergency department (ED), inpatient or outpatient (routine) setting. Waiting lists could include number of people on a waiting list or the number of people waiting more than a defined period. Based on a preliminary literature search, a waiting time framework was developed that provides further description of waiting times. Waiting time categories identified were: Routine care/outpatient initial consultation waiting times; Inpatient waiting times for surgery or other treatment; Routine care/outpatient diagnostic waiting times; Elective and routine care treatment waiting times; Emergency waiting times at a hospital; Time to follow-up; Social care waiting times; Other Waiting Times
4. OECD: Organization for Economic Co-operation and Development https://www.oecd.org/en/about/members-partners.html

### 5.2 Literature search

A comprehensive search of bibliographic databases was conducted for English language publications from 2015 to January 2025. The Well-being of Future Generations Act (2015), which provides a policy directive for public bodies to work on integrated approaches in Wales, was published in 2015. Therefore, this time limit was used for this rapid review.

Searches were limited to the OECD countries, as healthcare systems in these geographical locations may be more comparable to the UK context.

The following bibliographic databases were searched:

- On the OVID platform: Medline, Embase
- On the EBSCO platform: CINAHL
- Cochrane CENTRAL
- Scopus

An initial search of Medline, Embase, CINAHL, and Cochrane Library was undertaken (using the following concepts: Integrated care/pathway/system or Coordinated care or Collaborative care or co-management or partnership/joint working AND waiting times or waiting lists) followed by analysis of the text words contained in the title and abstract, and of the index terms used to describe the articles. This informed the development of the main search strategy which was tailored for each information source (see Appendix 1). Forward and backward citation tracking was completed using Citationchaser and relevant studies were be added to the review.

The websites of key UK third sector and government organisations, Google Advanced, and the Overton database were also searched for grey literature reports (see Appendix 2).

Additionally, clinical trial registers (Clinicaltrials.gov, and WHO International Clinical Trials Registry Platform (ICTRP)) were searched for completed trials, the findings of which may not have been published in peer-reviewed journals. Additionally, systematic and scoping review evidence identified via the bibliographic and grey literature searches was checked for relevant primary studies that were not found via other methods.

Forward and backward citation searching was completed using Citationchaser. Records identified via citation searching were screened to make sure that no relevant studies were missed. As the identified evidence was very similar (hip fracture and its management via MDT) to what was included via other methods, the decision was made not to include these in the final report to ensure that the rapid review was manageable within the timeframes.

### 5.3 Study selection process

All citations retrieved from the database searches were imported or entered manually into EndNote^TM^ (Thomson Reuters, CA, USA) and duplicates removed. Following deduplication, the citations that remained were exported as a Txt file and then imported to Rayyan^TM^. Two reviewers dual screened at least 20% of citations using the information provided in the title and abstract using Rayyan^TM^. Any conflicts in the title and abstract screening were resolved by a third reviewer. The rest of the citations were screened by a single reviewer. For citations that appeared to meet the inclusion criteria, or in cases in which a definite decision could be made based on the title and/or abstract alone, the full texts of all citations were retrieved. The full texts were screened for inclusion by two reviewers and any disagreements were resolved by a third reviewer. The flow of citations through each stage of the review process is displayed in a PRISMA flowchart (Page et al. 2021).

### 5.4 Data extraction

For quantitative primary research studies, all relevant data were extracted directly into tables by one reviewer and checked by another. Microsoft Excel^TM^ was used for initial data extraction and mapping, while detailed data extraction was performed in Microsoft Word^TM^. The data extracted included specific details about the included primary research studies (research design, methods), the populations, the interventions (type, length, setting and country), and waiting time outcomes as described within the primary research.

For qualitative primary research studies, relevant data on research design, methods, populations, and interventions (type, setting and country) and the type of waiting time (e.g.: diagnostic waiting times, time to treatment, etc.) were extracted. All qualitative findings (data extracts (quotes), interpretations) were extracted independently by one reviewer and checked by another and the software package NVIVO^TM^ was be used to facilitate this process.

### 5.5 Study design classification

The study designs were classified based on the definitions and algorithm developed by the Public Health Agency of Canada (2014). Studies were classified based how participants were chosen (natural or deliberate intervention/exposure), how many groups were assessed, at what time points participants were assessed, and whether allocation to groups were random. Based on these criteria, studies could be classified as case-control, cohort study, RCT, non-randomised controlled trial, controlled before and after study, interrupted time series, or uncontrolled before and after study. Classification is necessary as some study designs have inherent flaws due to methods used or participant selection, among others.

The guidance by Public Health Agency of Canada (2014) rates the strength of these different study designs, which can help grading the overall body of evidence.

Different study designs can be rated as follows:

- **Strong:** RCTs, non-randomised controlled trials, controlled before and after (more than two groups)
- **Moderate:** controlled before and after studies (only two groups), cohort studies, case control studies, interrupted time series (three or more pre and post assessment)
- **Weak:** Uncontrolled before and after studies, interrupted time series (less than three pre or post assessment)

### 5.6 Quality appraisal

The Analytic Study Critical Appraisal Tool (CAT) is a unified tool that can assess the methodological quality of RCT, non-randomised controlled trials, controlled before and after studies, lab-based studies, cohort studies, case-control studies, interrupted time series studies and uncontrolled before and after studies (Moralejo et al. 2017, Public Health Agency of Canada 2014). It was anticipated that a wide variety of quantitative study designs would be included in this rapid review, thus the decision was made to use the CAT tool. The CAT has 14 items each of which can be rated as strong, moderate or weak.

An overall quality decision can be made based on responses to items 2 to 12, which could be high, medium or low. The overall quality of quantitative studies in this rapid review was determined based on criteria by Public Health Agency of Canada (2014). Some modifications were made to ensure relevancy to the studies included. Criteria with modifications can be seen below.

- **Rate the quality as HIGH if**: “most [at least 7] or all appraisal items were rated as strong, and none were rated as weak. In addition, there are no major threats to internal validity of the study or the ability to draw the conclusion that there is a clear association between the exposure and the outcome of interest.” (Public Health Agency of Canada 2014, p. 43).
- **Rate the quality as MEDIUM if**: “appraisal items 4 and/or 11 are rated as at least moderate, and the other appraisal items rated as weak or moderate are not sufficient to compromise the internal validity of the study. Also, these other items do not interfere with the ability to draw the conclusion that there is a probable association between the exposure and the outcome of interest.” (Public Health Agency of Canada 2014, p. 43).
- **Rate the quality as LOW if**: “appraisal items 4 and/or 11 are rated as weak, or if other items rated as weak are sufficient to interfere with the ability to rule out other explanations for the findings and draw a conclusion about the association of the exposure and the outcome of interest.” (Public Health Agency of Canada 2014, p. 43). Additionally, studies were rated low if they contained more weak items than moderate. If items 8 (comparability of control group and intervention group) and item 9 (adequacy of control of major confounders) were rated weak, the overall study quality should be low.

To assess the methodological quality of qualitative primary research studies, the 10-item JBI checklist for qualitative research was used (Lockwood et al. 2015). When a study met a criterion (question answered as “Yes”) a score of one was given. When the answer to an item was regarded as “unclear” or “no”, it was given a score of zero. If a question was regarded as “not applicable” this point was taken off the total score. Overall scores were presented by adding up points for each applicable question. In addition to overall judgements and scores, a textual description of methodological quality was also be provided for both quantitative and qualitative primary research studies.

### 5.7 Synthesis

Informed by the JBI mixed methods guidance, this rapid review adopted a segregated mixed methods approach, given that both quantitative and qualitative data were considered for inclusion. This approach entailed performing separate syntheses of quantitative and qualitative findings to ensure that each type of data is appropriately analysed while contributing to a comprehensive understanding of the research question (Lizarondo et al. 2020).

Quantitative and qualitative data was reported narratively and organised into separate thematic summaries (Thomas et al. 2017). Thematic summaries were based on the coding of population, interventions, and waiting time outcomes. The framework developed by Baxter et al. (2018b) was the basis of coding interventions. Waiting time descriptions were categorised based on the framework developed for the preliminary literature review^4^, which contained the following categories:

1. Routine care/outpatient initial consultation waiting times
2. Inpatient waiting times for surgery or other treatment
3. Routine care/outpatient diagnostic waiting times
4. Elective and routine care treatment waiting times
5. Emergency waiting times at a hospital
6. Time to follow-up
7. Social care waiting times
8. Other Waiting Times

Additionally, intervals described in the published works of Weller et al. (2012) and Neal et al. (2015) were also considered for categorisation.

### 5.8 Assessment of body of evidence

To assess the overall body of evidence a set of five items suggested by Moralejo et al. (2017) were considered. The items included: the strength of study design; the CAT quality decisions; the number of studies evaluating the same population; directedness of the evidence; and consistency of results. Based on these items, the overall body of evidence can be described as strong, moderate, or weak (Moralejo et al. 2017). Grading criteria is depicted below.

**Table 7:**
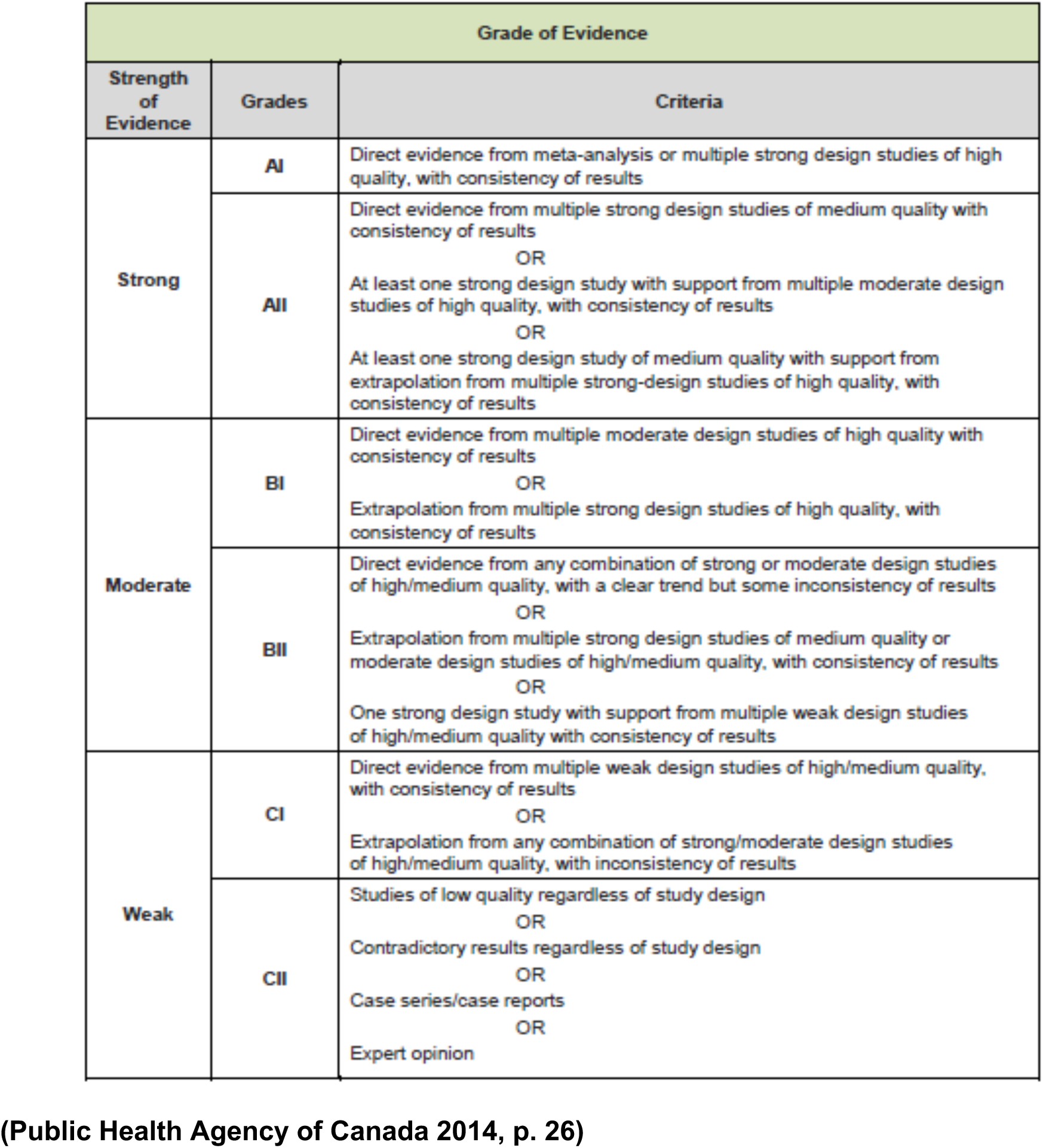
Grading criteria for the overall assessment of the body of evidence.

## EVIDENCE

### 6.1 Search results and study selection

**Figure 1:**
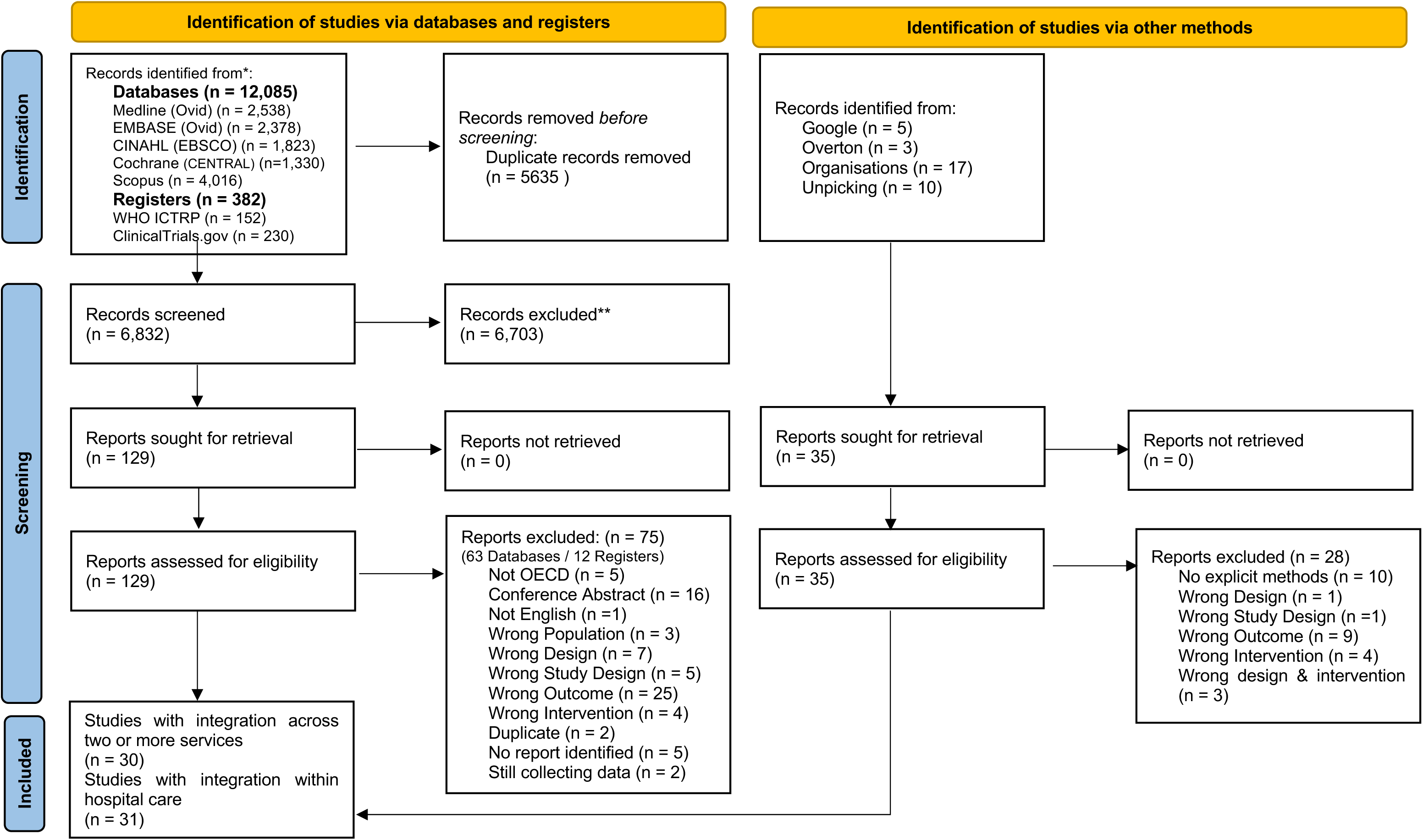
PRISMA 2020 flow diagram

### 6.2 Data extraction

**Table 8:**
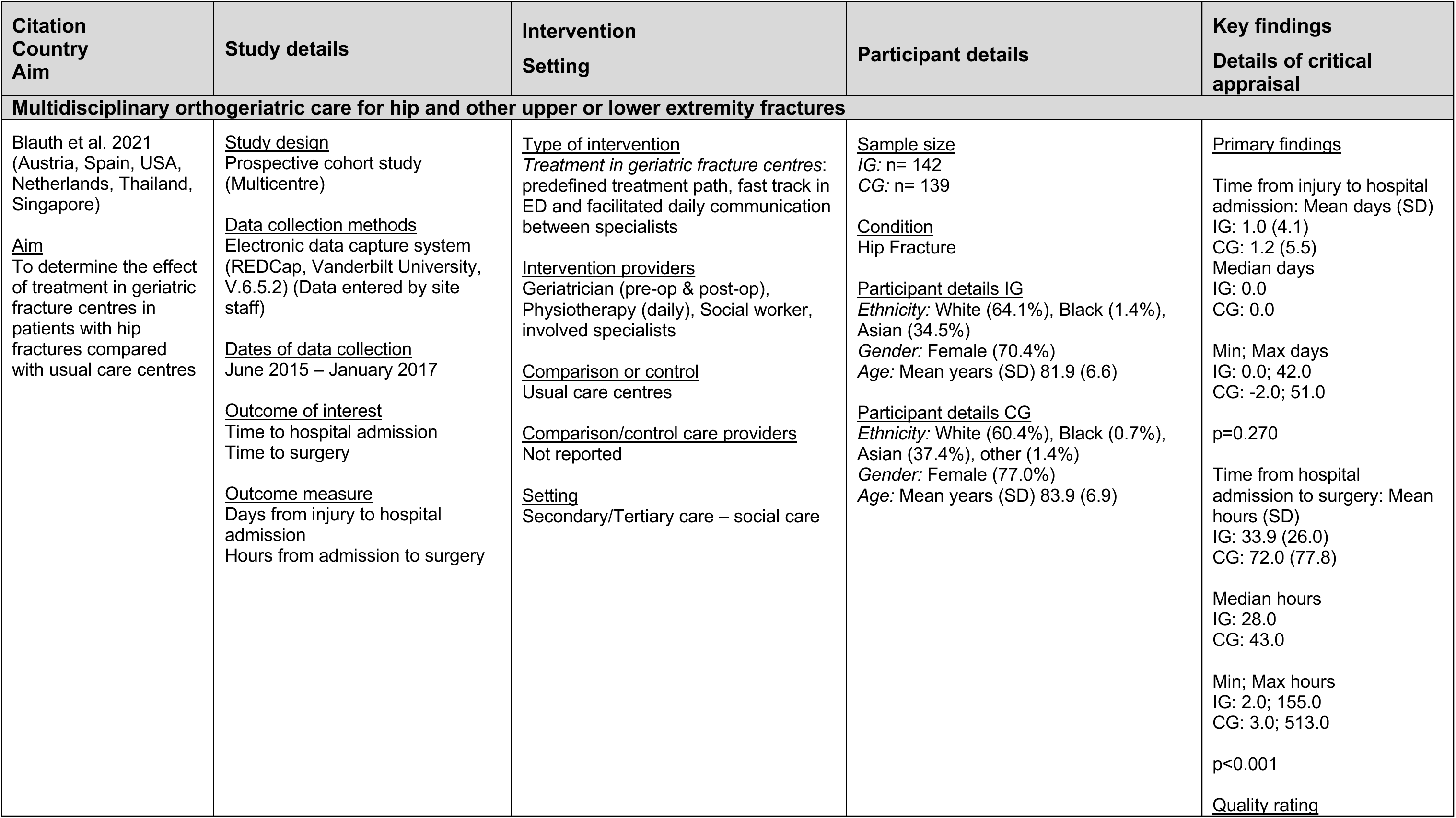
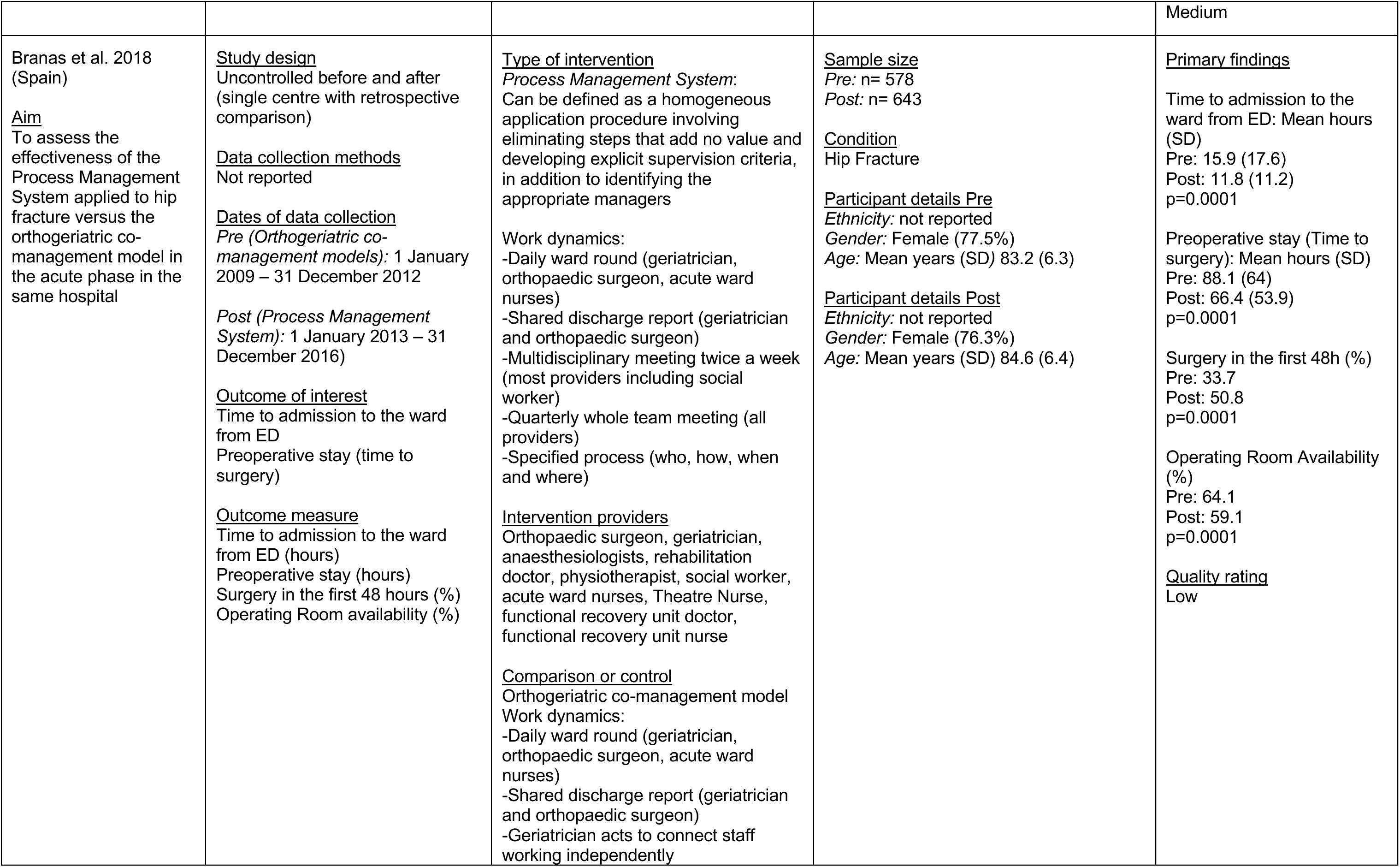
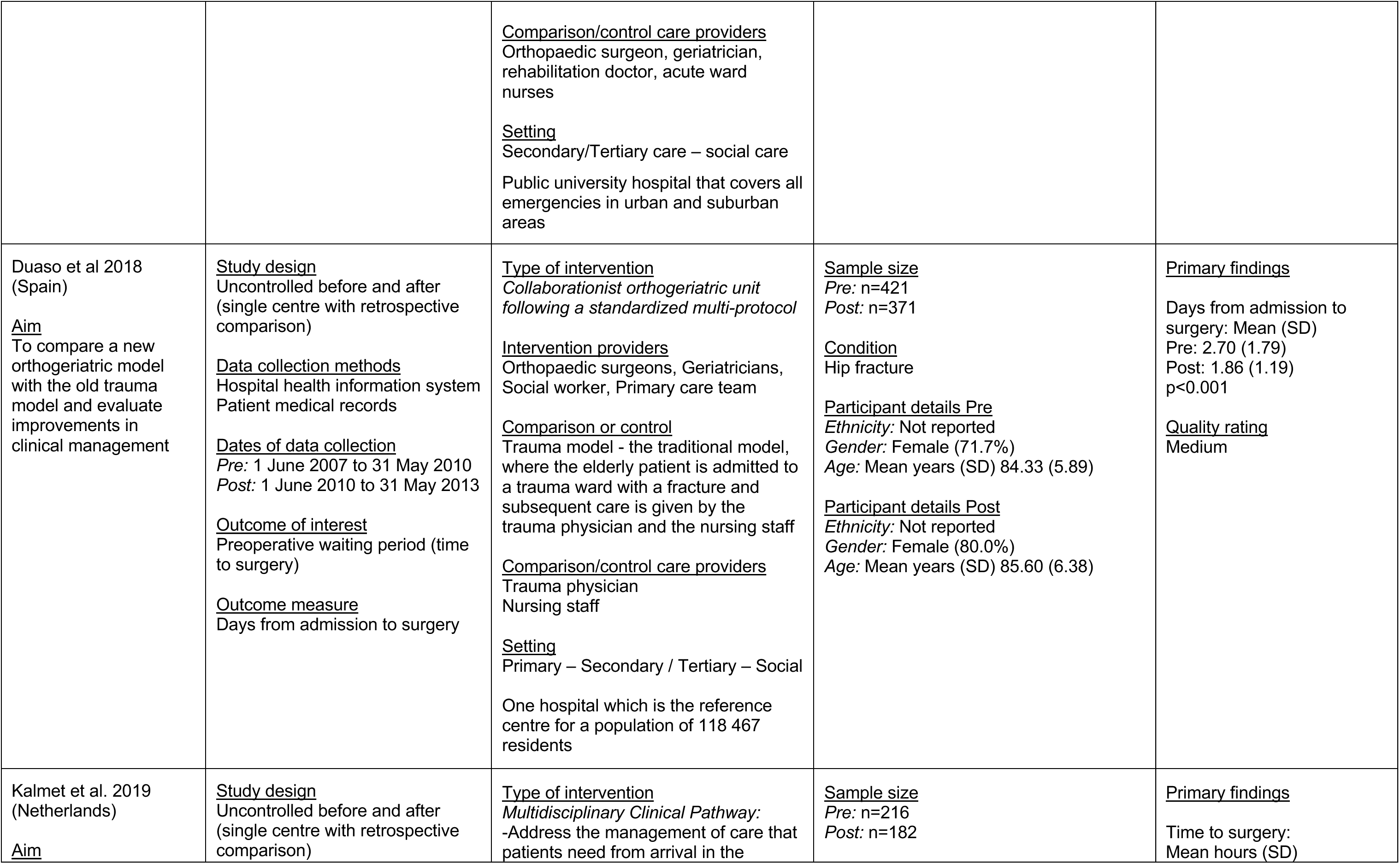
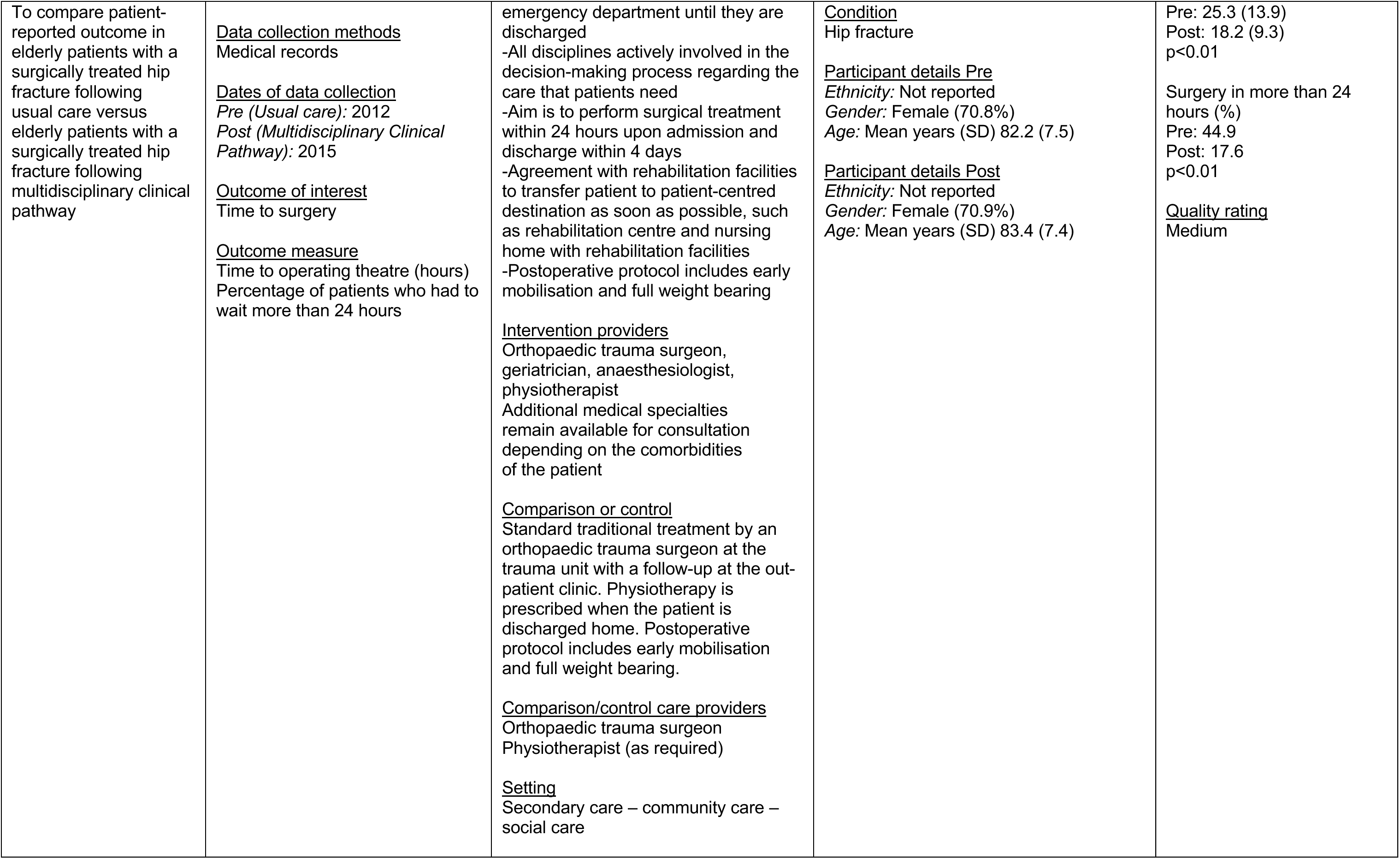
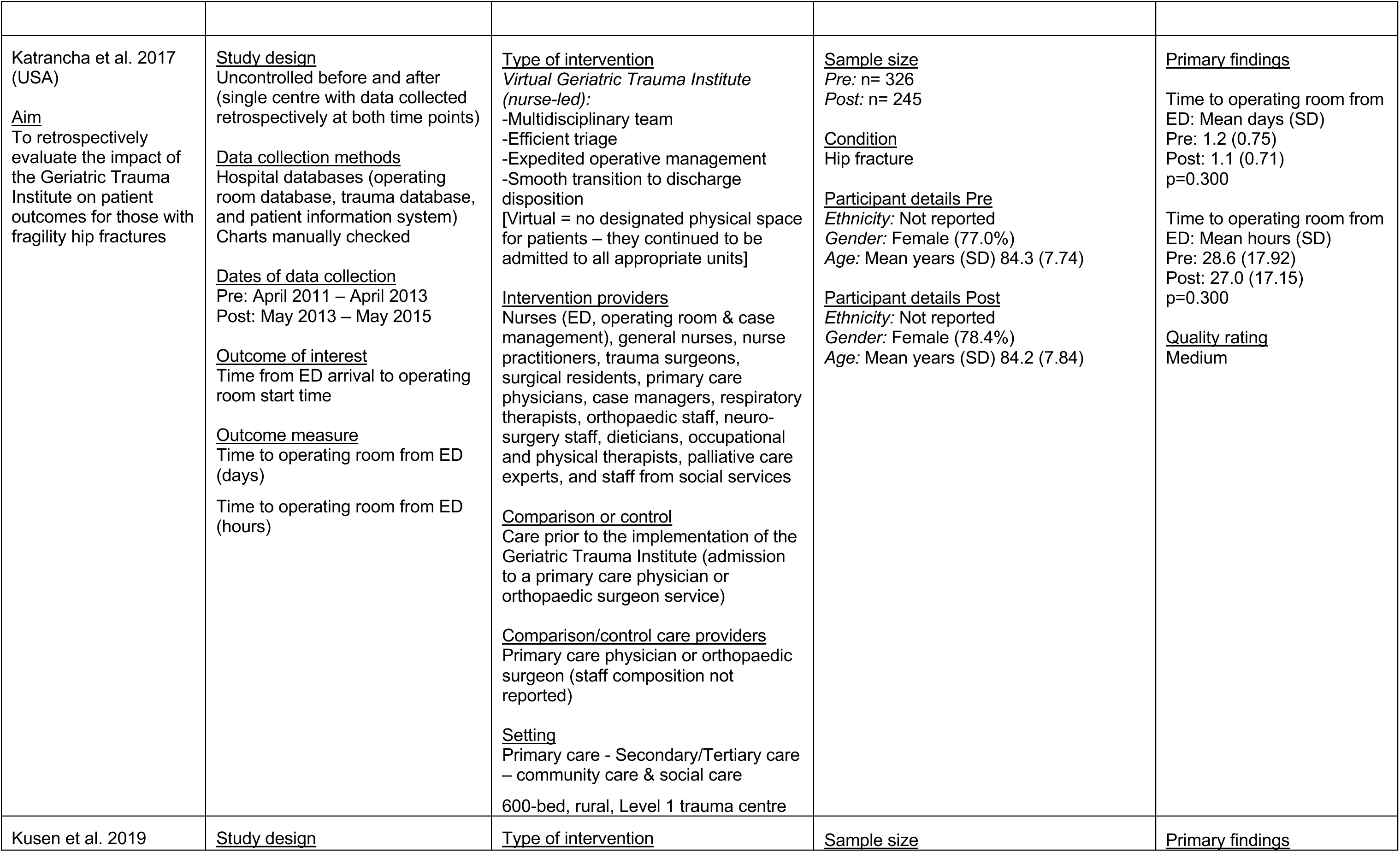
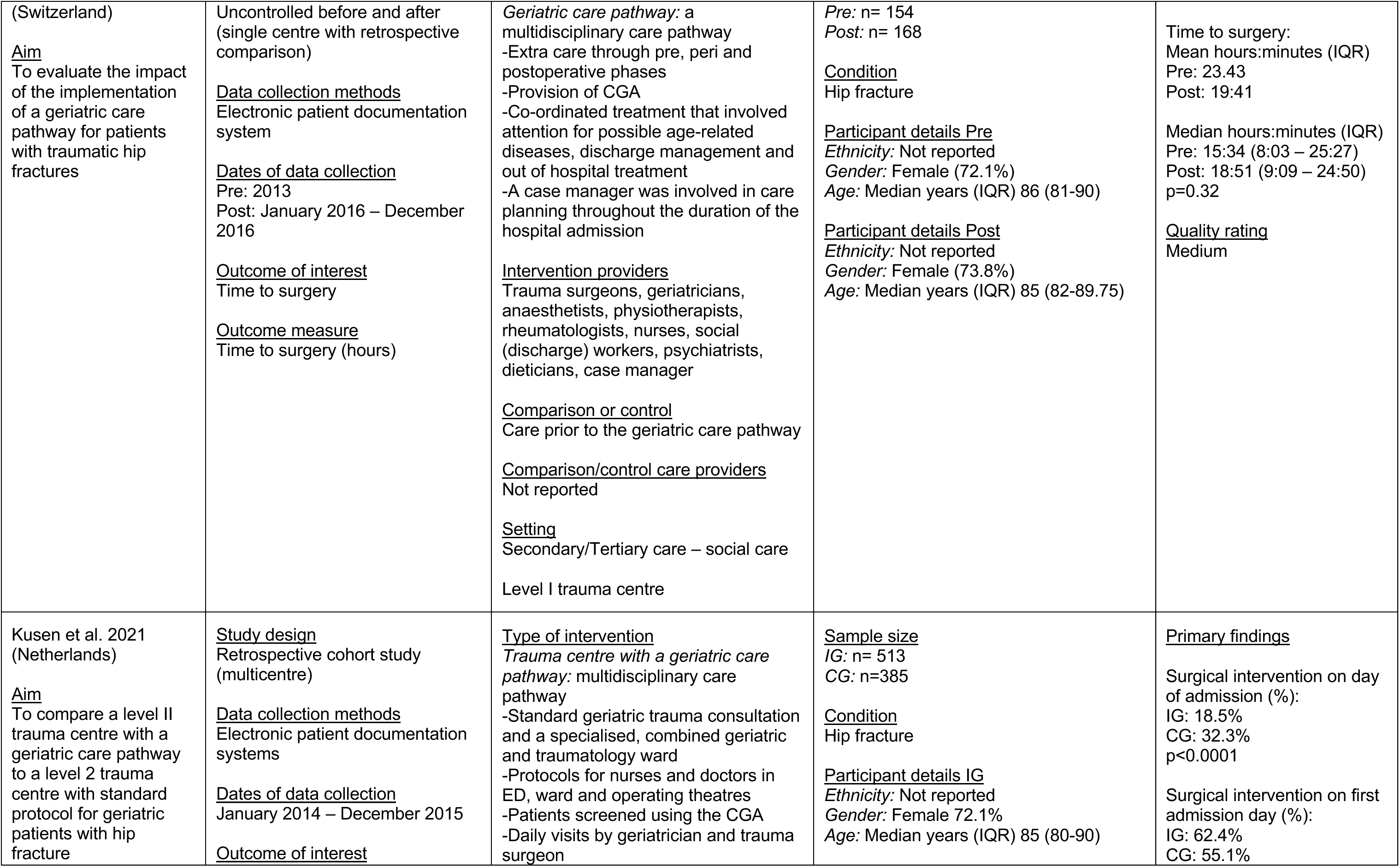
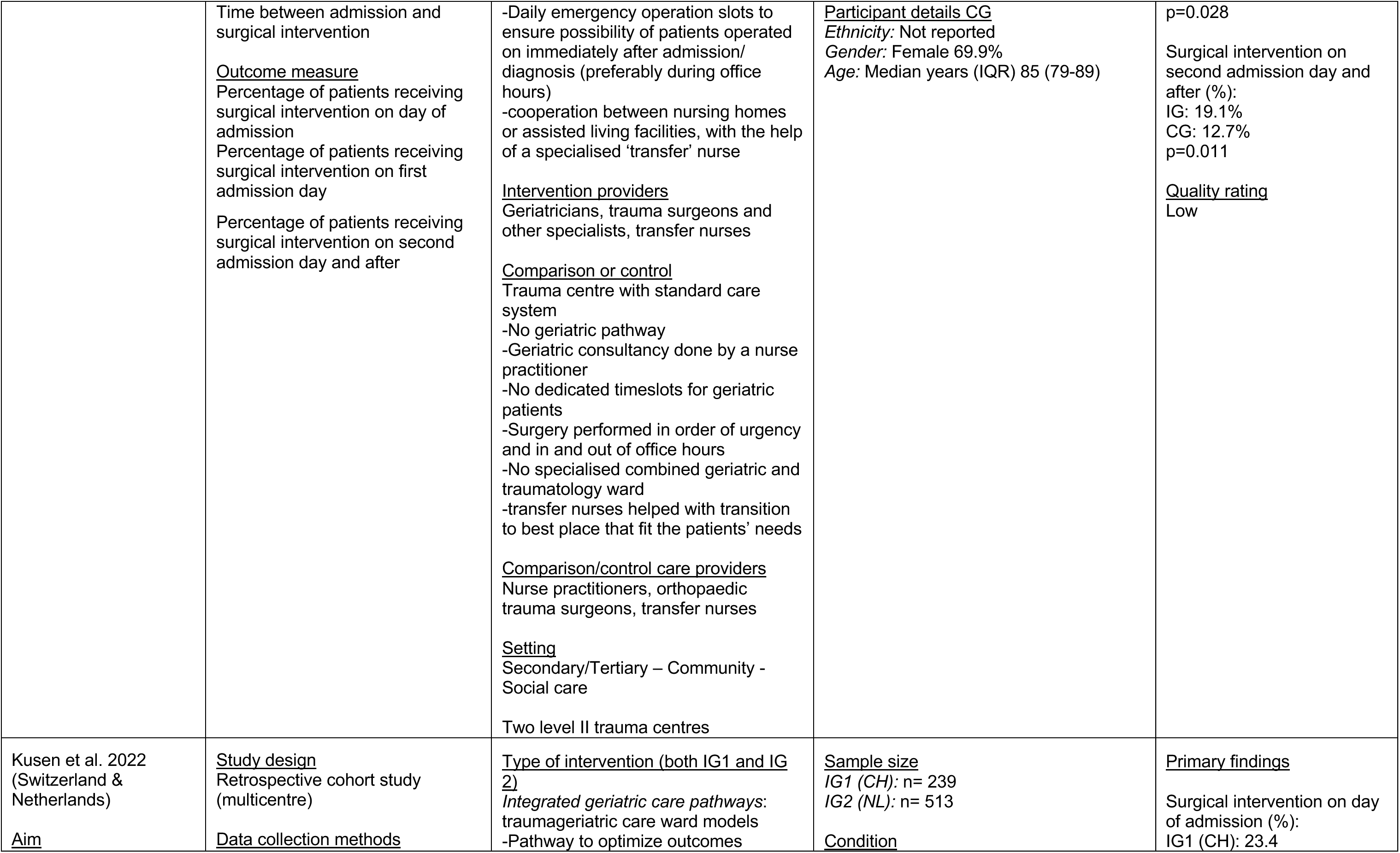
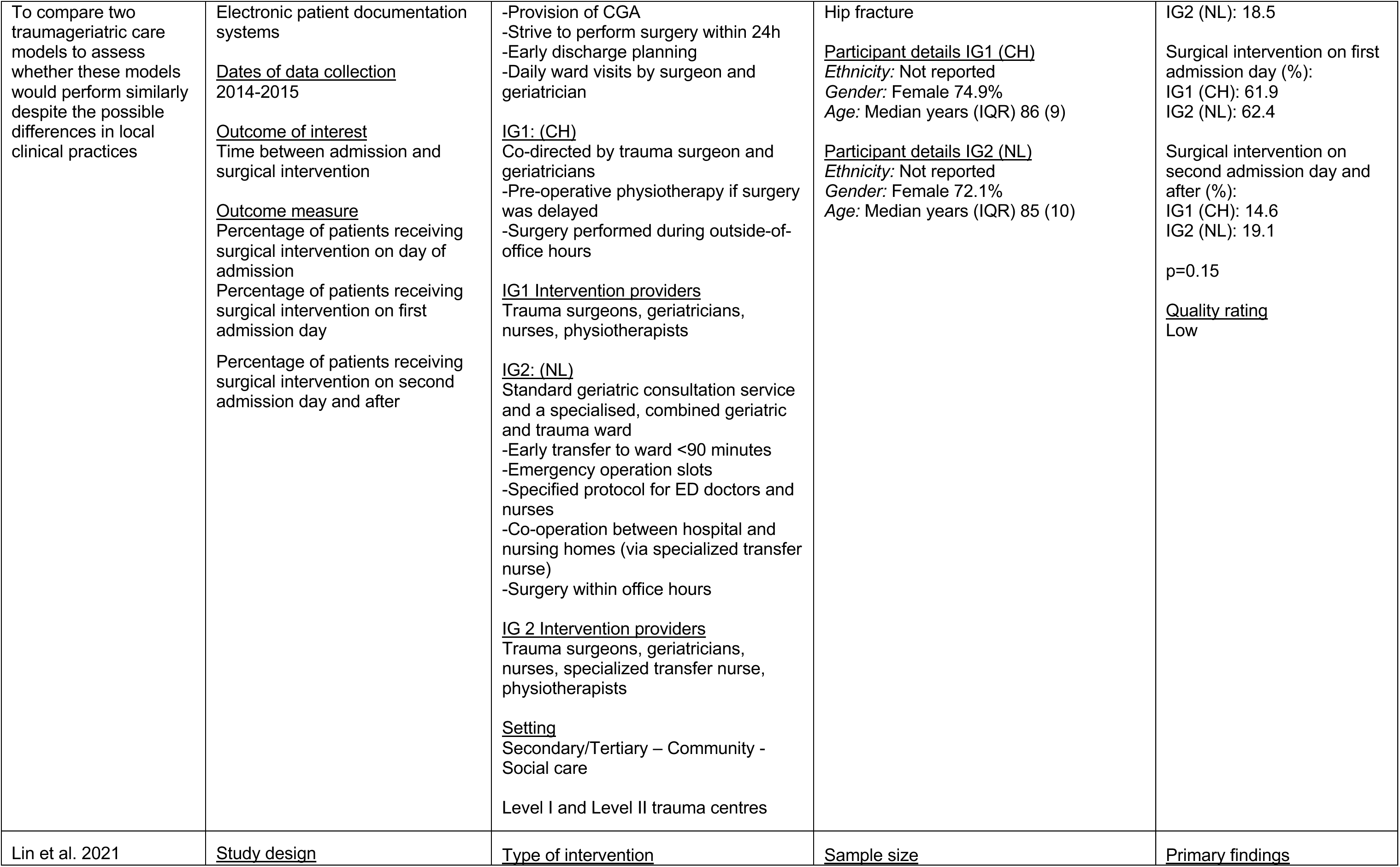
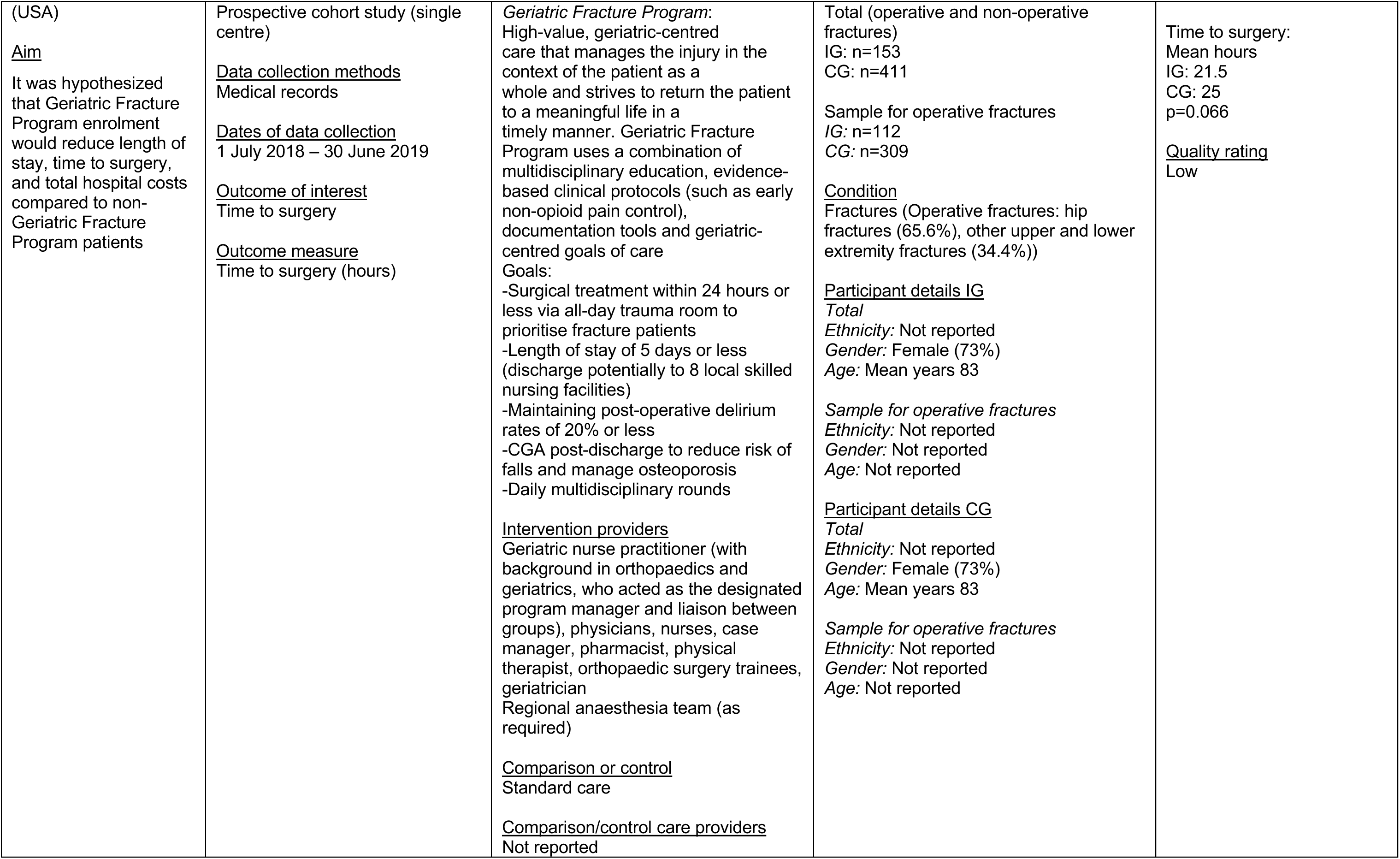
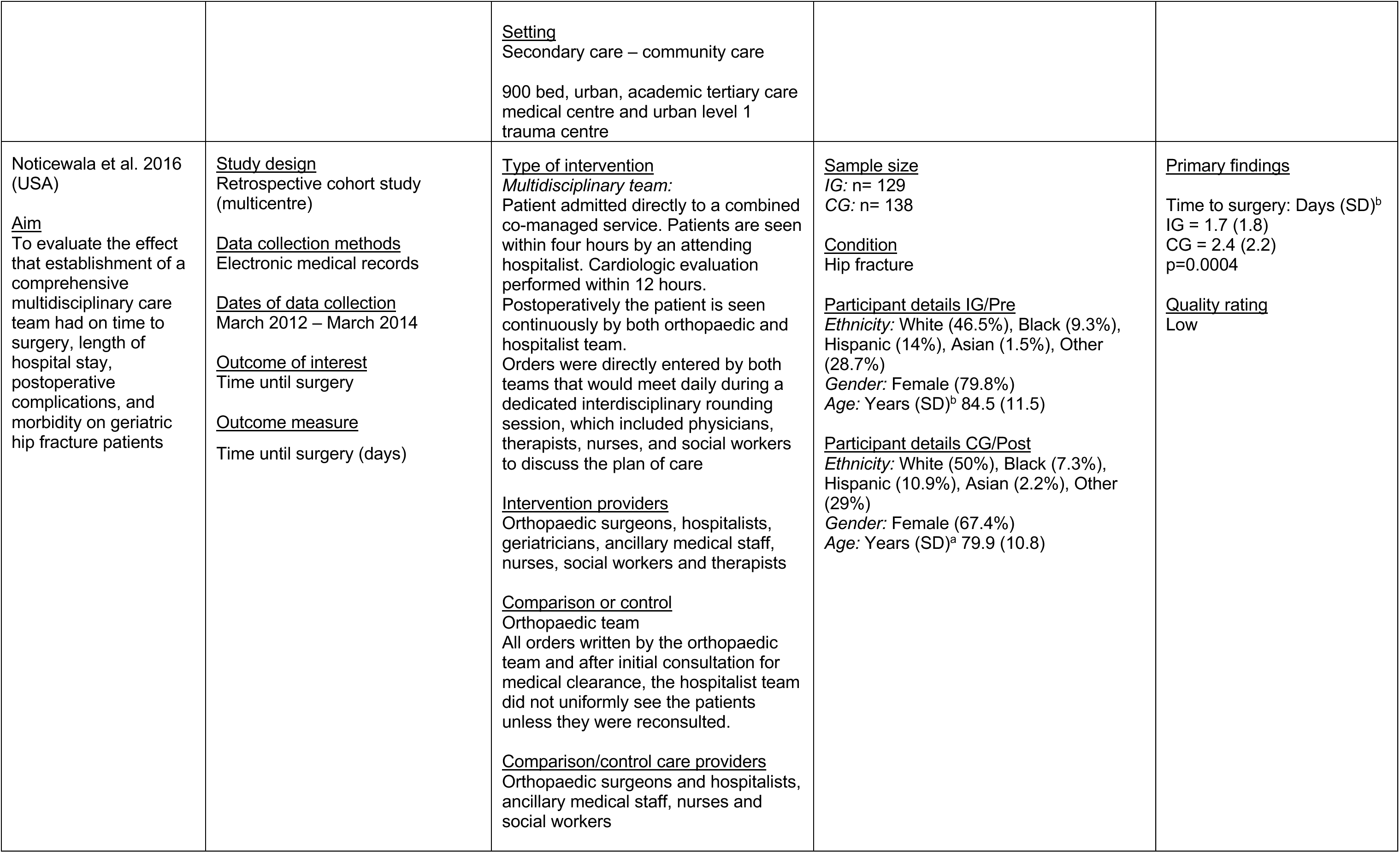
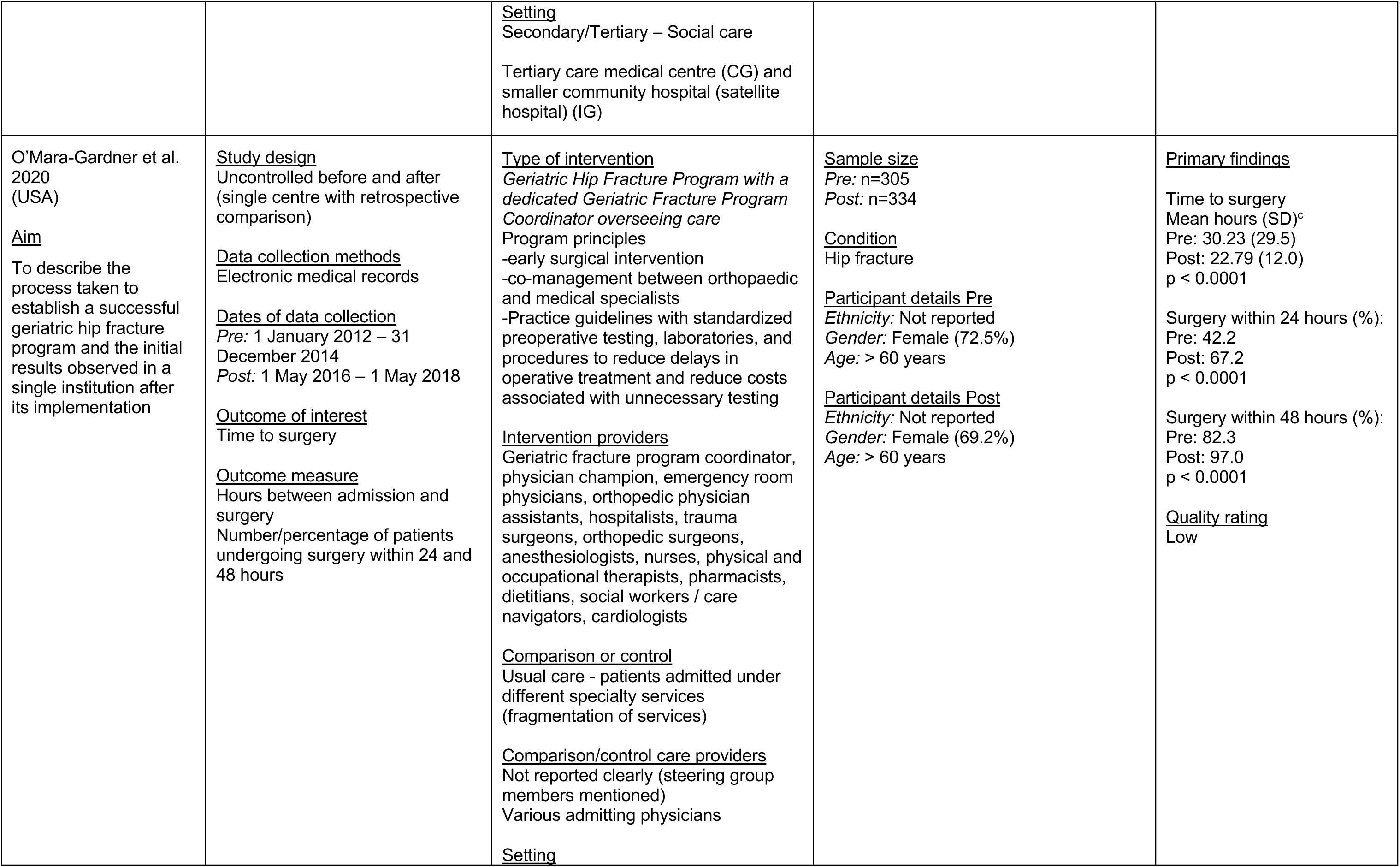
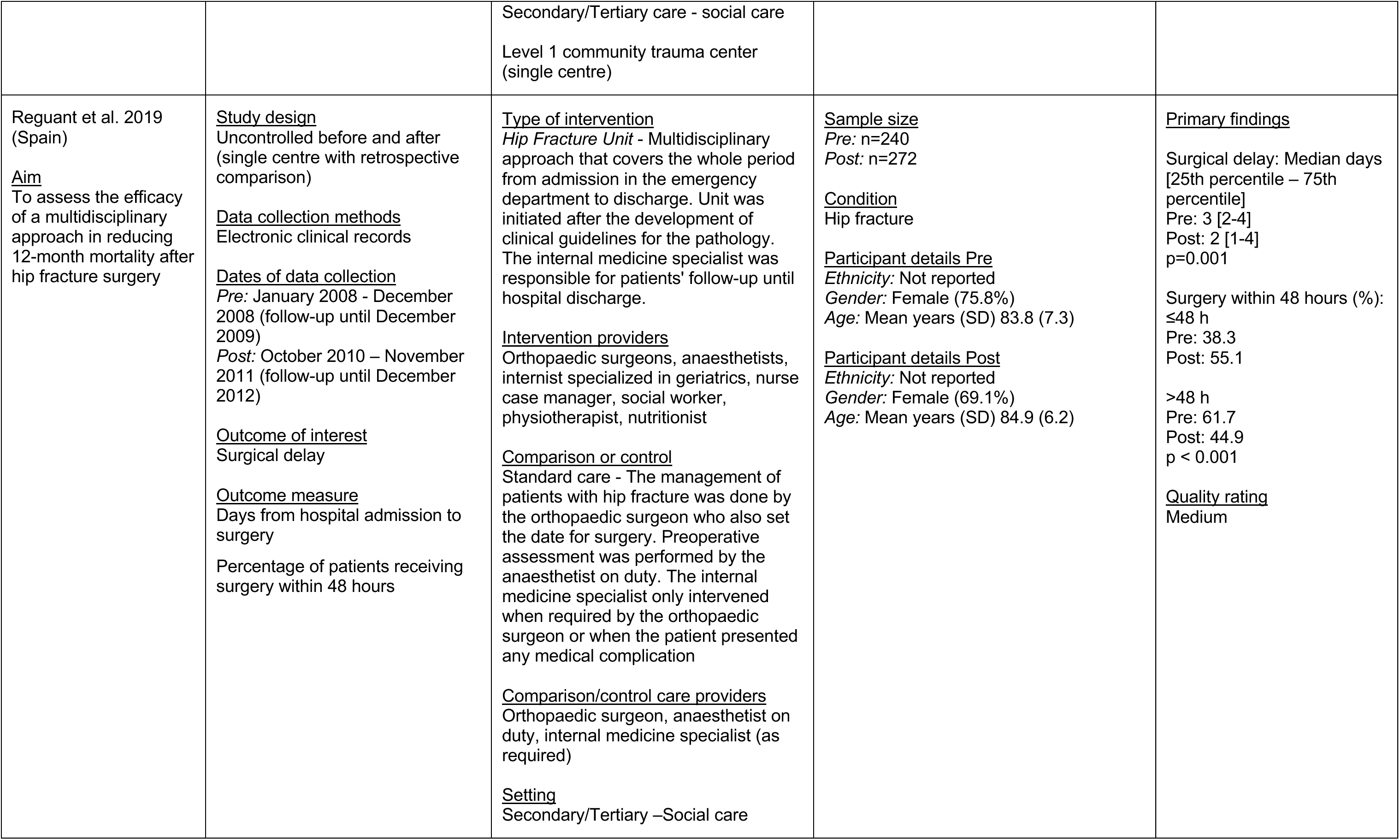
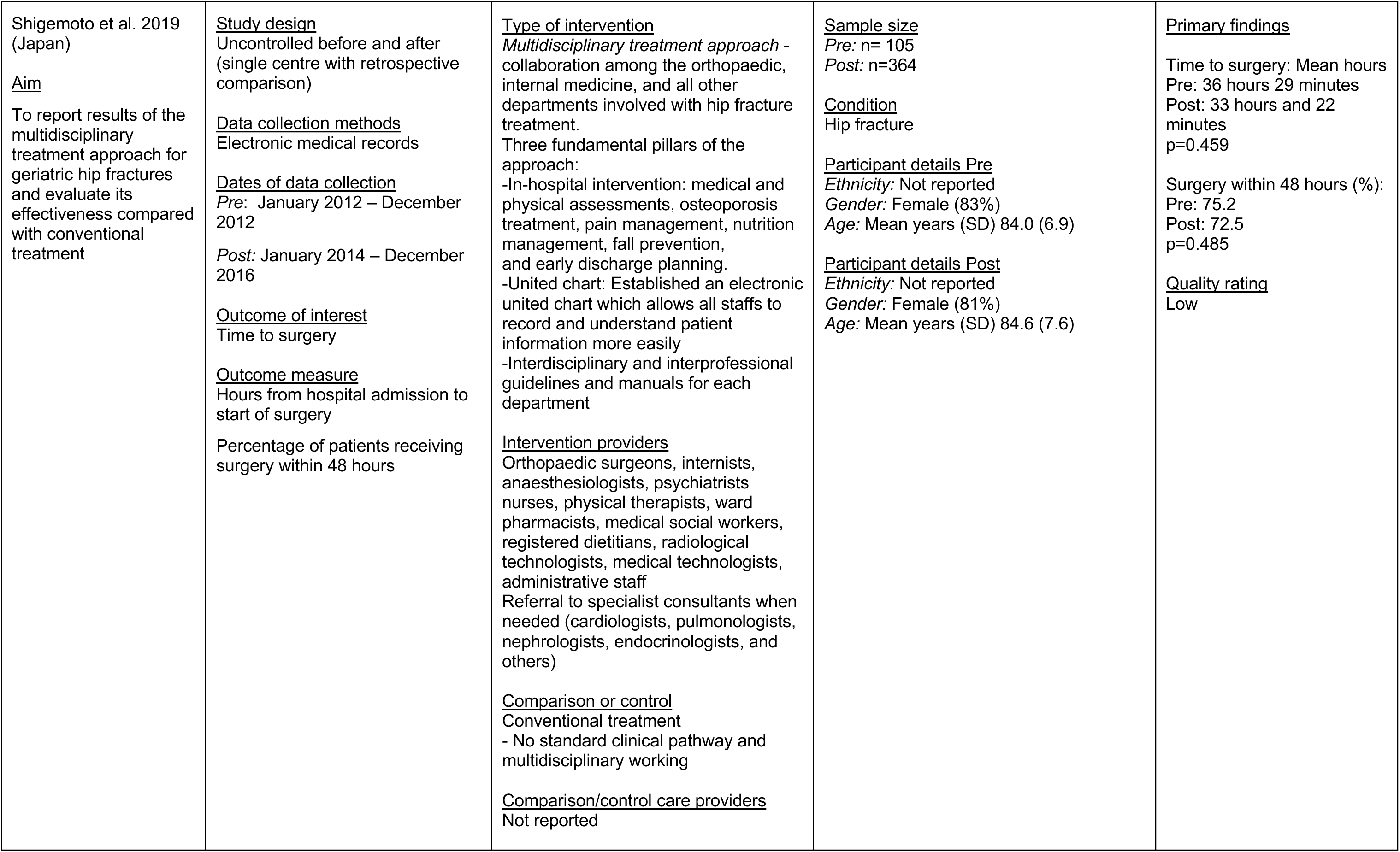
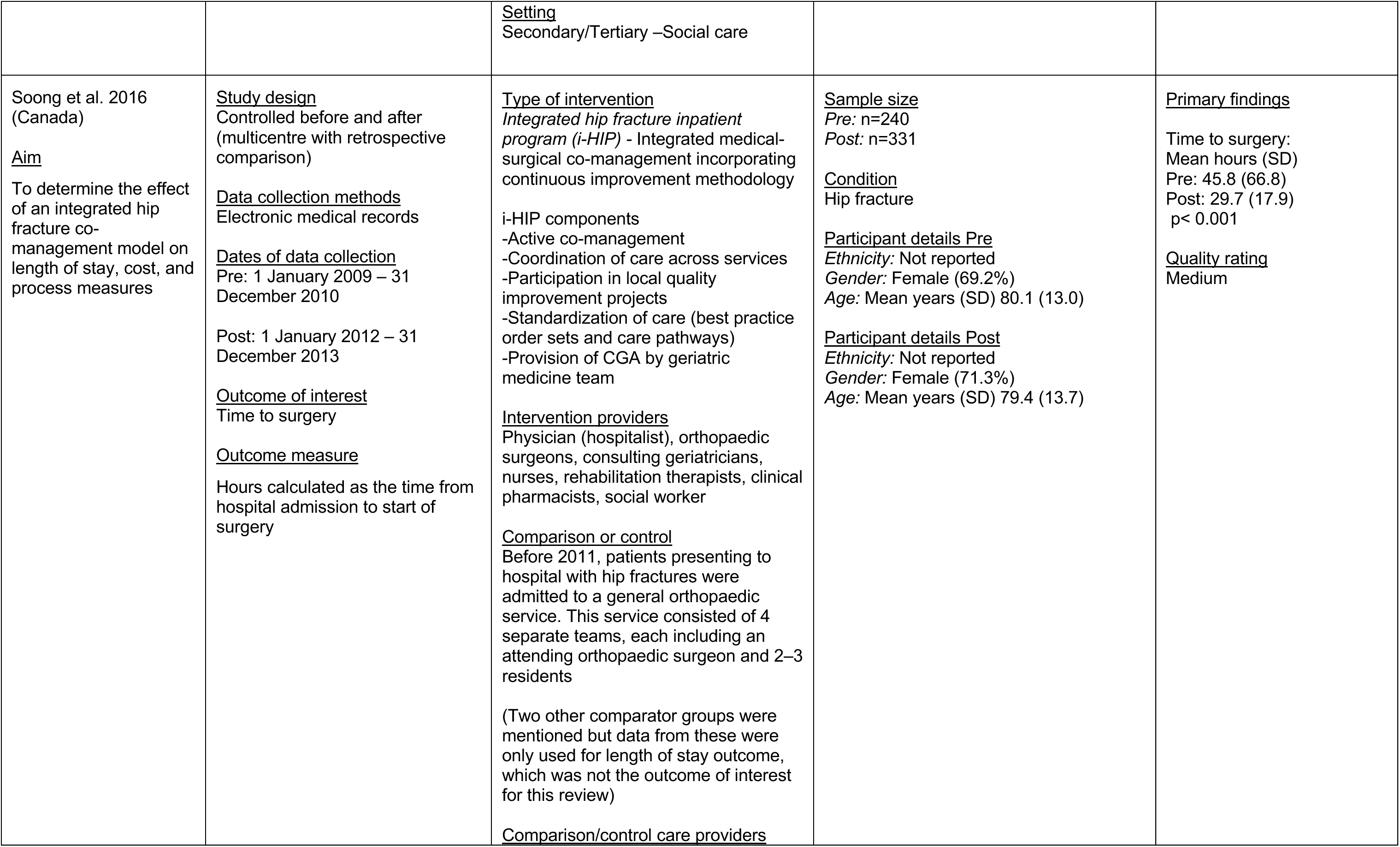
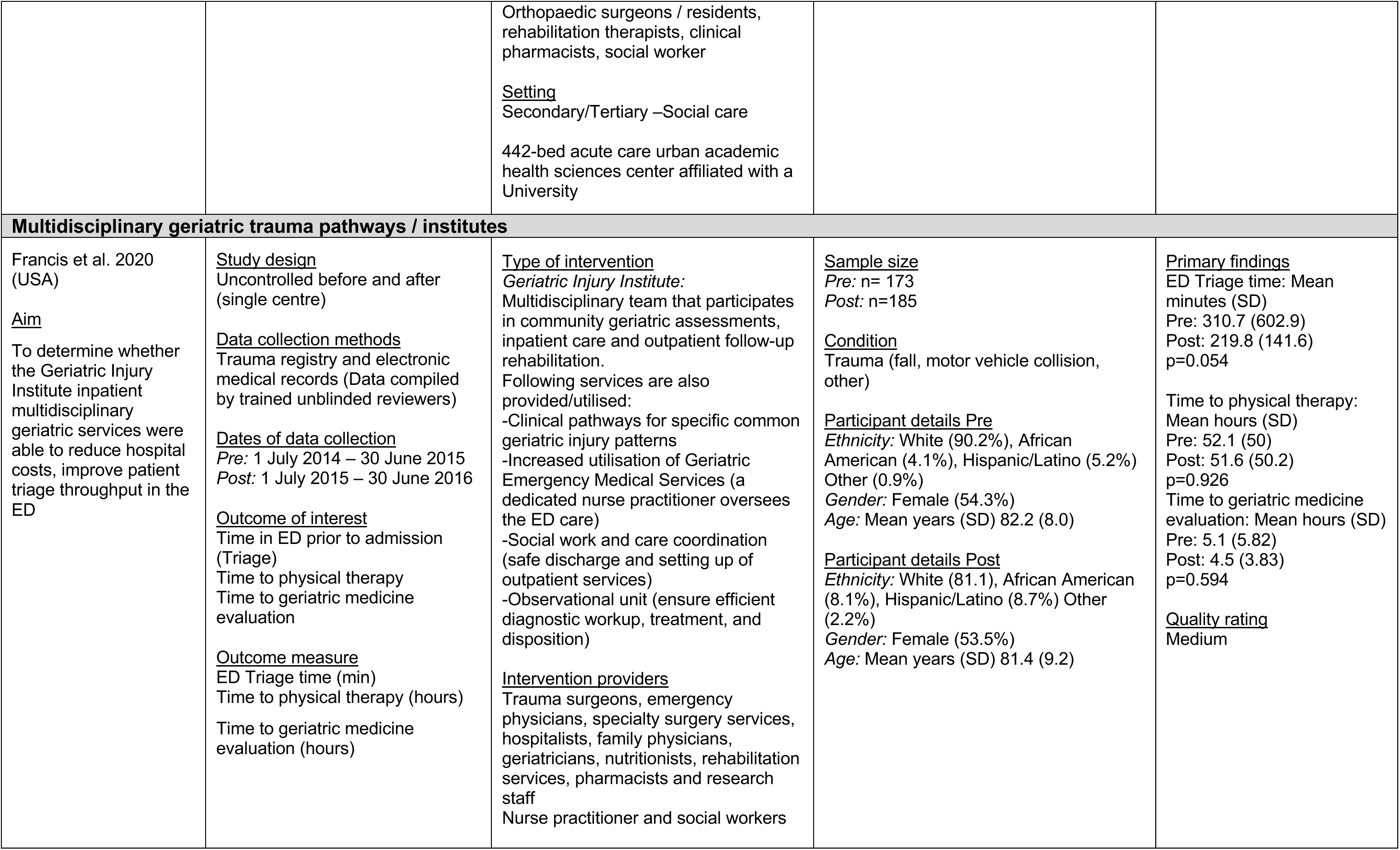
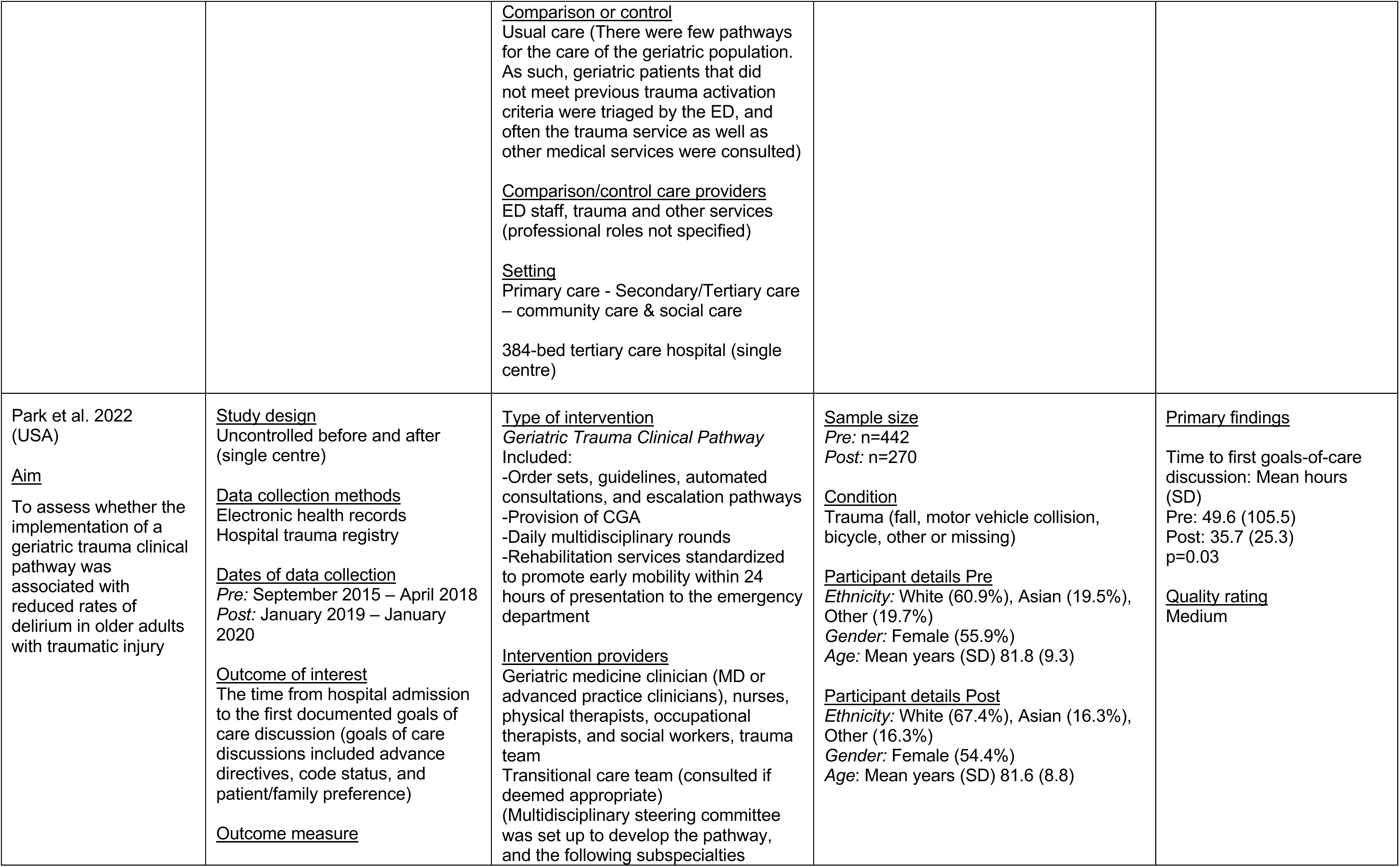
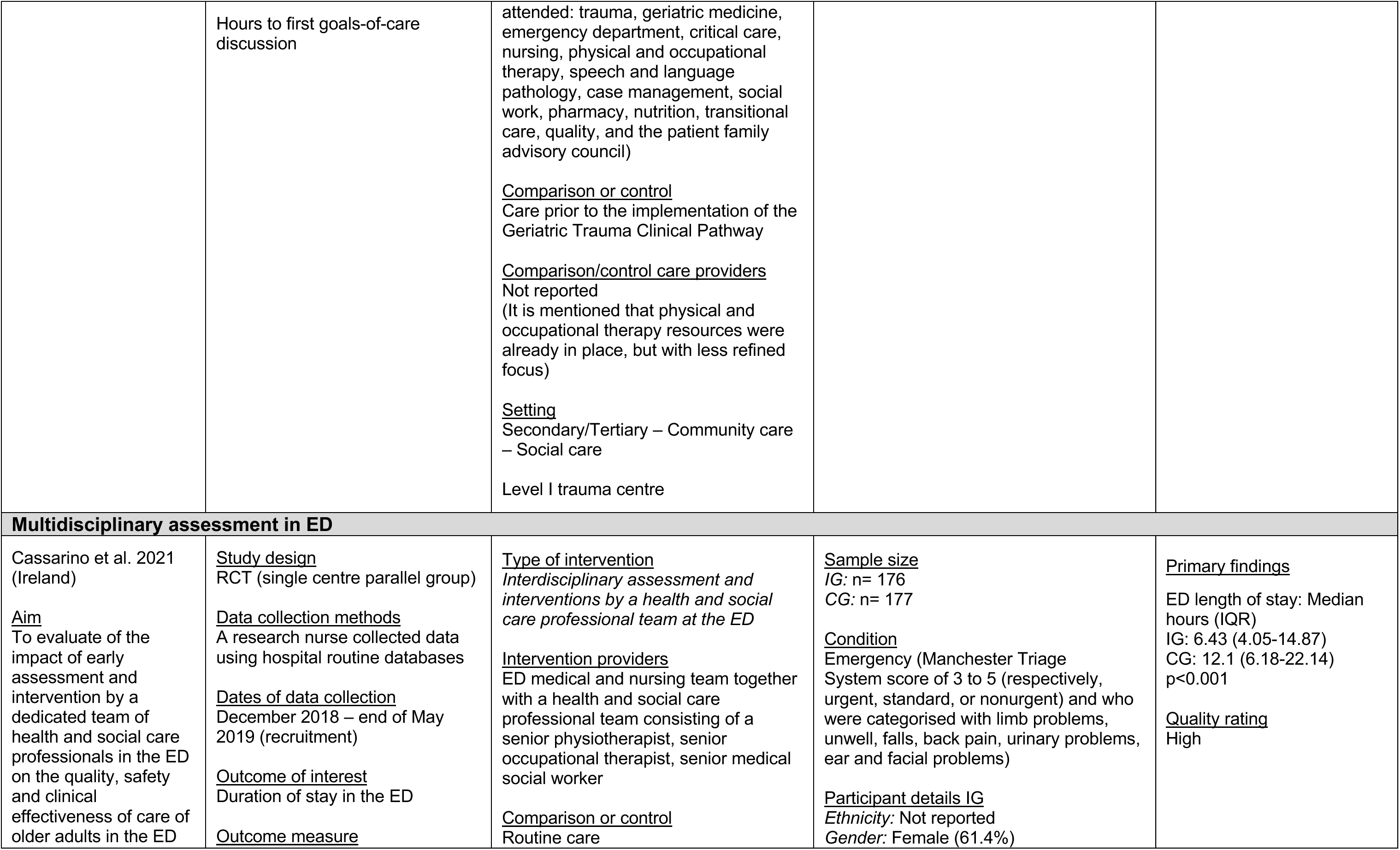
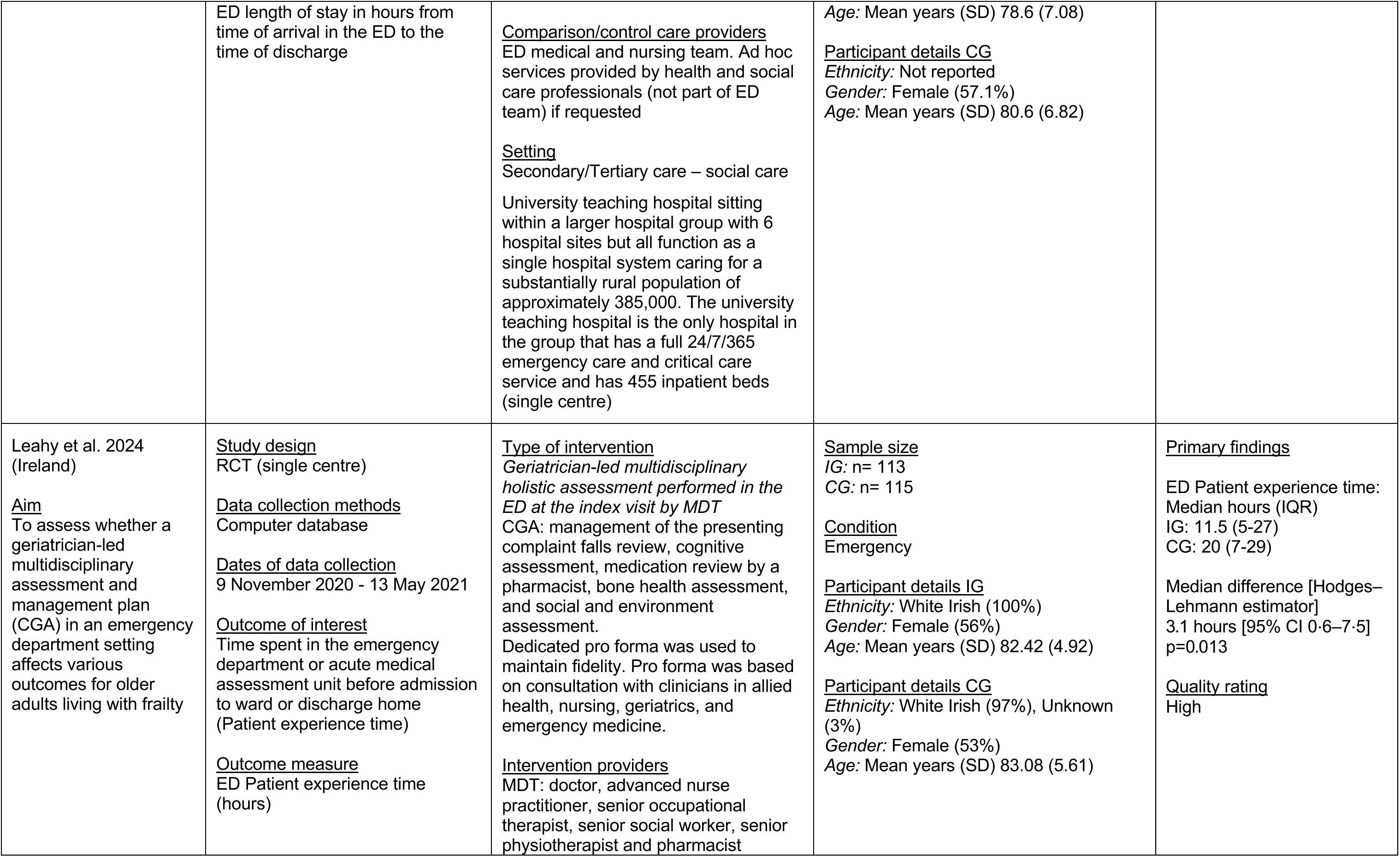
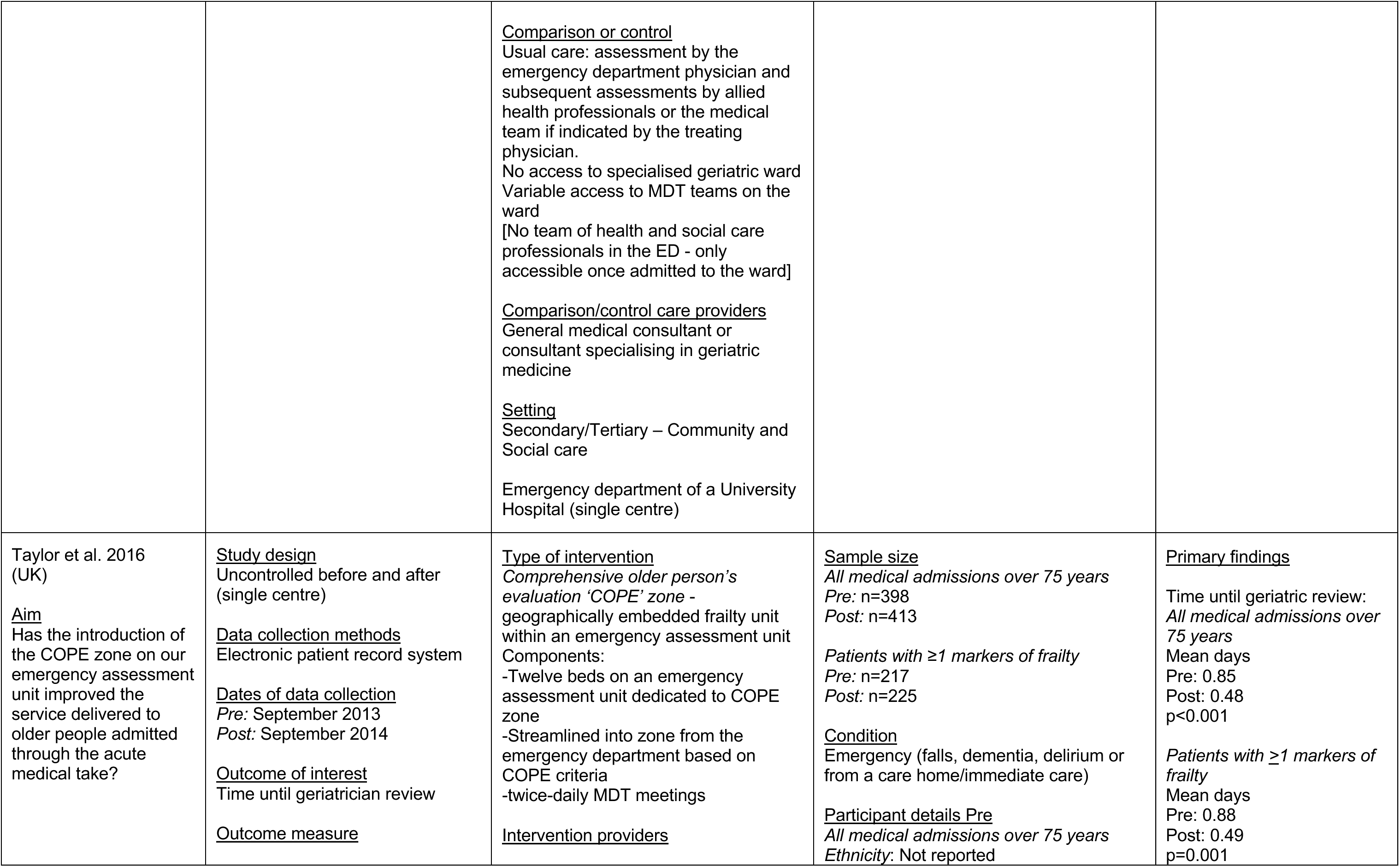
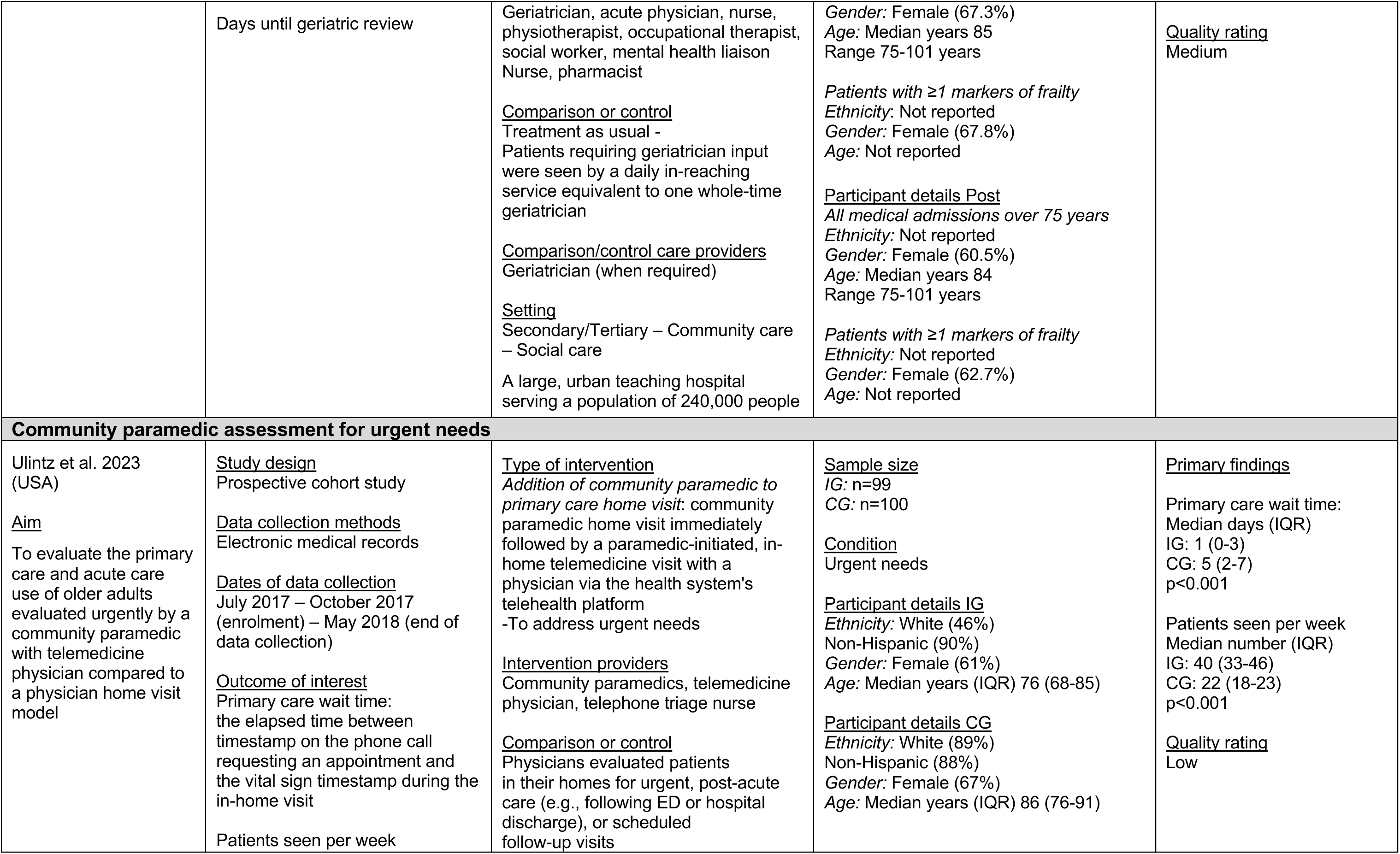
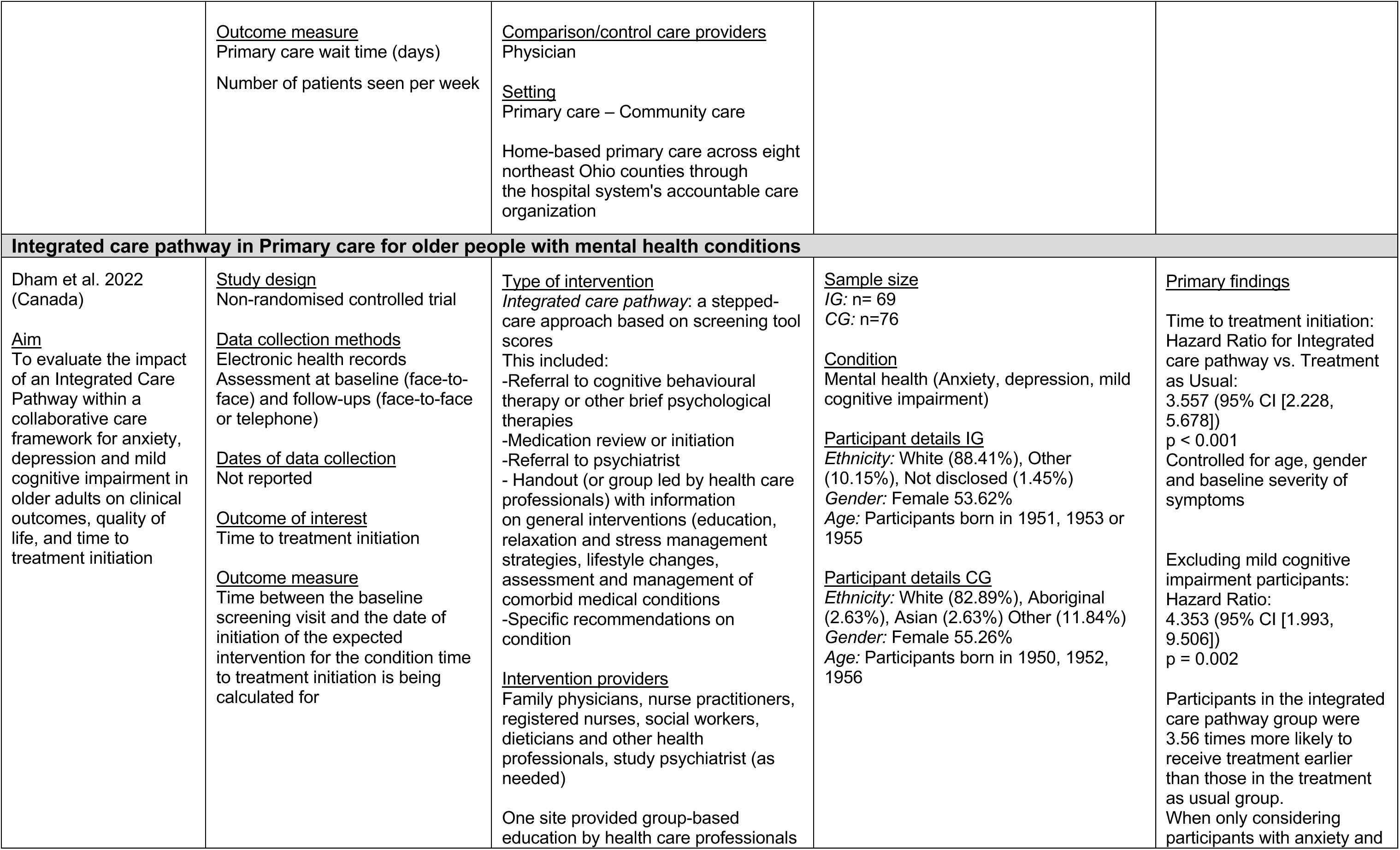
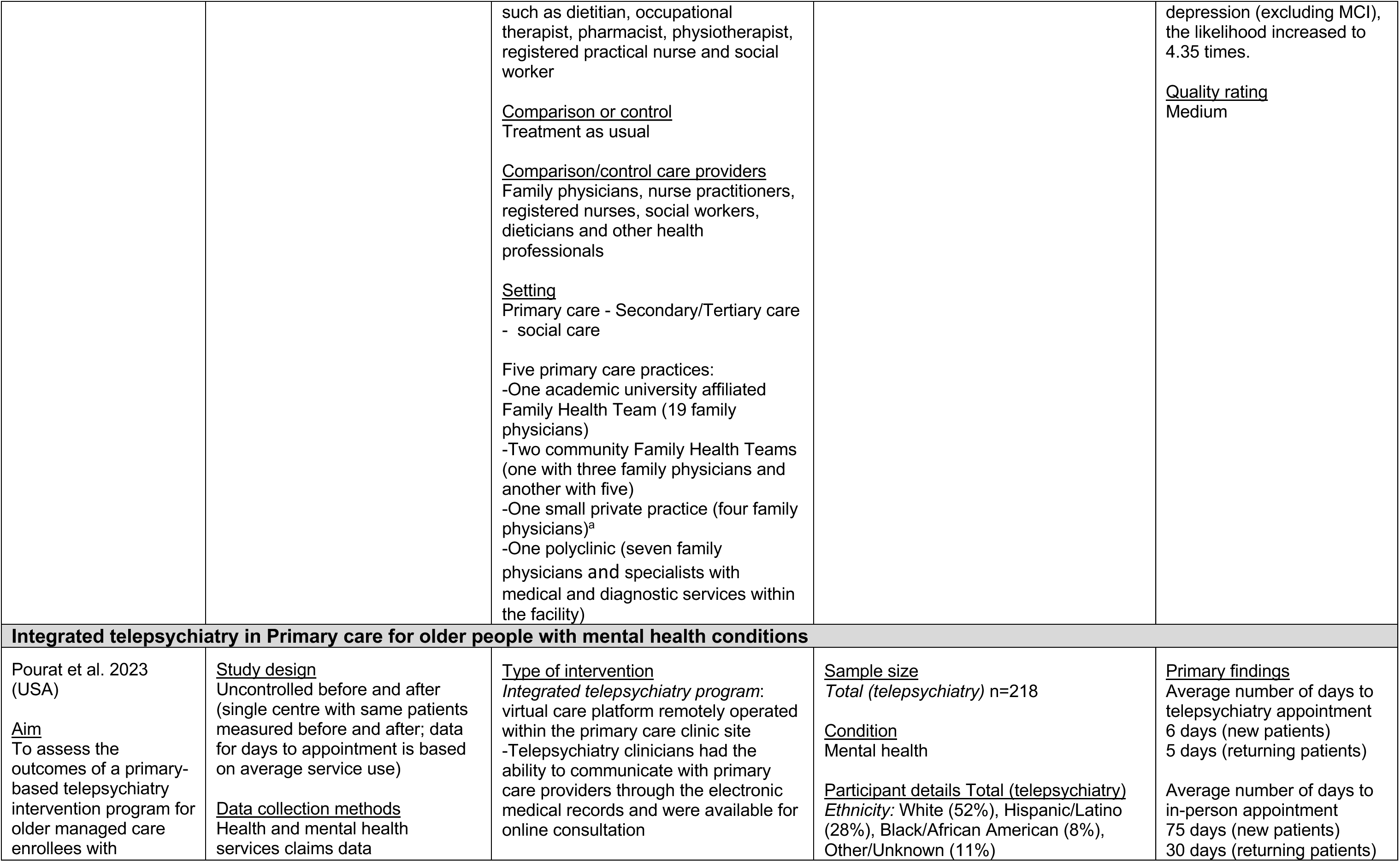
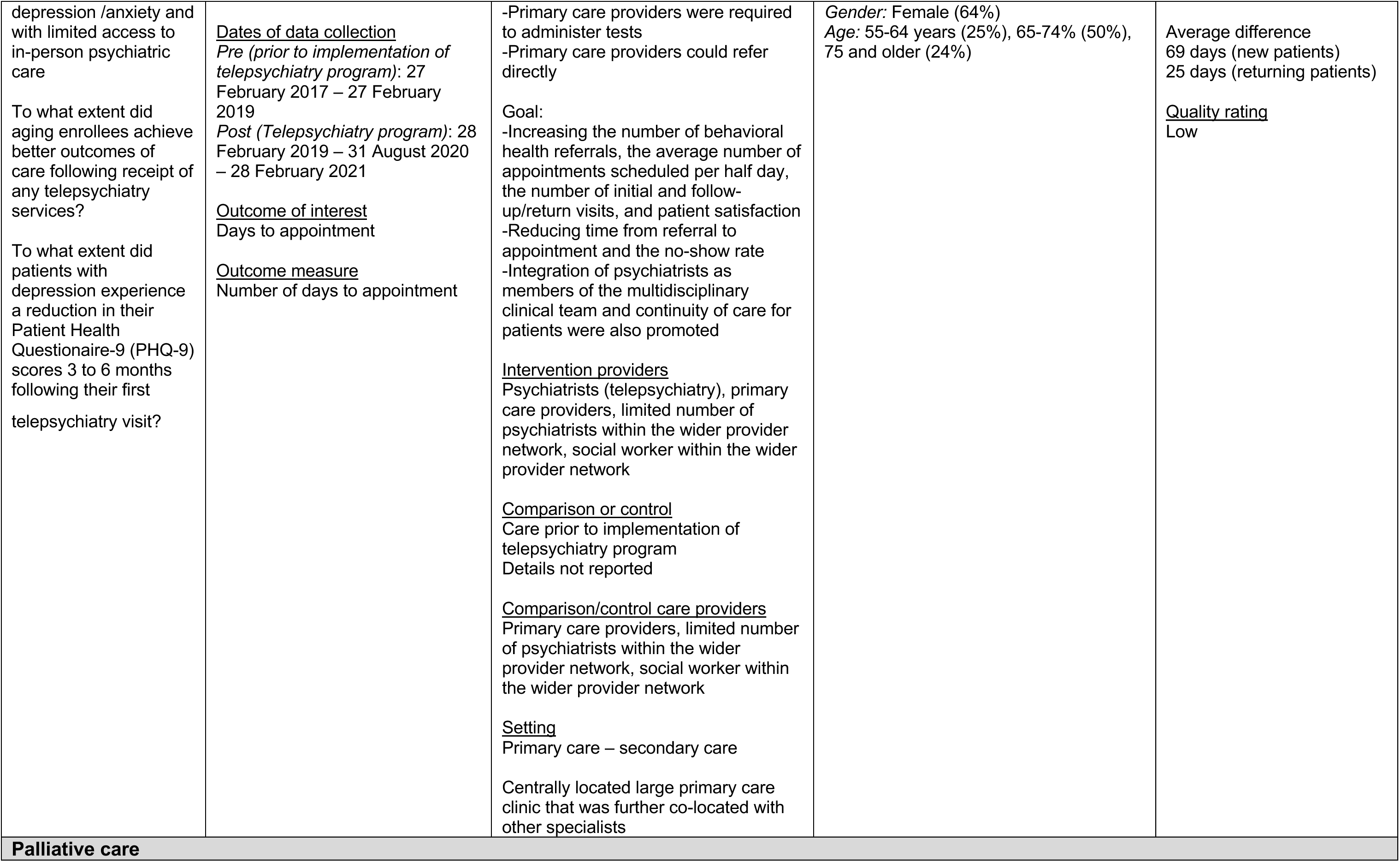
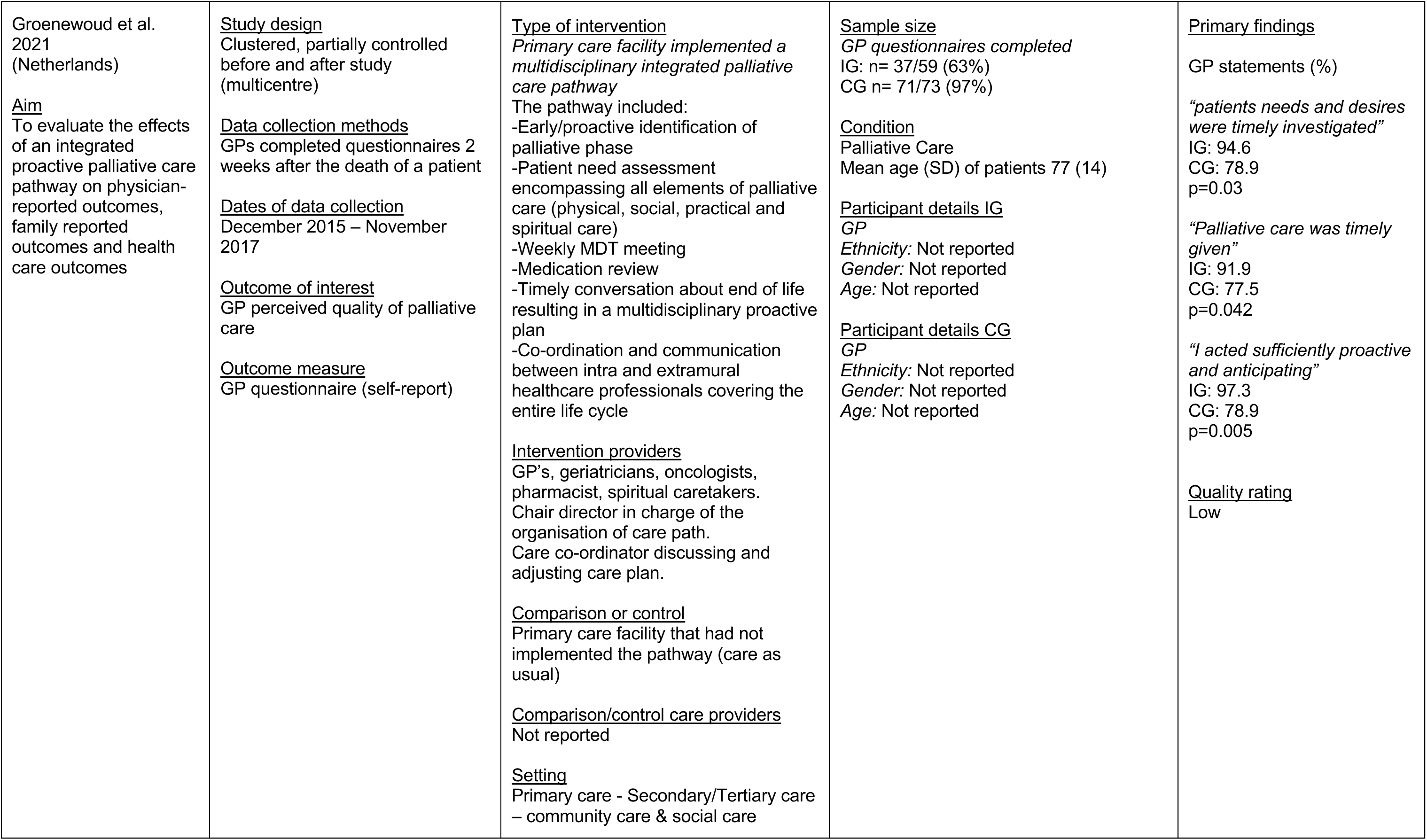
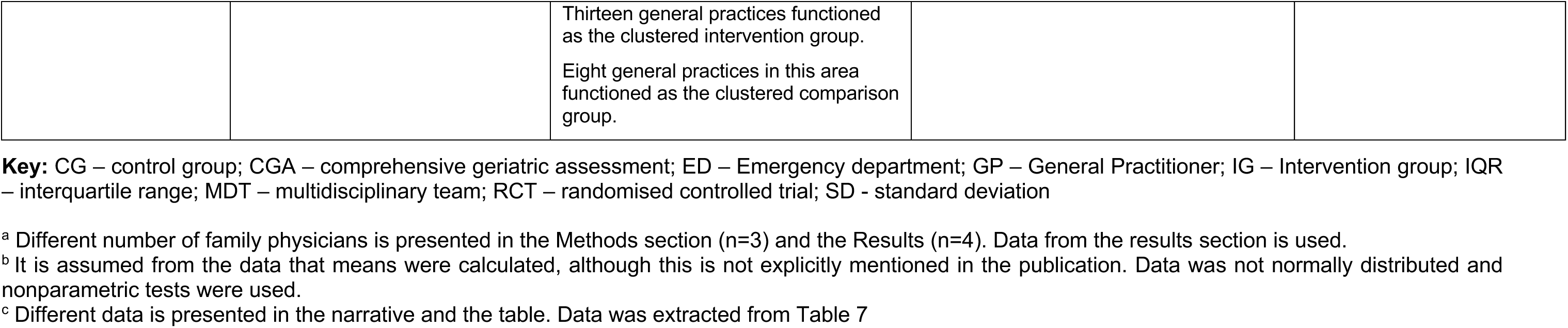
Summary of quantitative studies investigating integrated care operating across two or more services.

**Table 9:** Summary of qualitative studies exploring integrated care operating across two or more services.

| Citation<br>Country<br>Aim | Study details | Intervention<br>Setting | Participant details | Details of critical<br>appraisal |
| --- | --- | --- | --- | --- |
| Aberg & Ehrenberg<br>2017<br>(Sweden)<br><u>Aim</u><br>To describe factors of importance for the quality of hospital-based geriatric care from an inter-disciplinary team perspective | <u>Study design</u><br>Qualitative descriptive<br><u>Data collection methods</u><br>Focus groups (n=5)<br><u>Dates of data collection</u><br>Not reported | <u>Type of intervention</u><br>Interdisciplinary hospital based geriatric team care<br><u>Intervention providers</u><br>Occupational therapist, social workers, physician, physiotherapist, registered nurse, assistant nurse, access to dietitian and speech therapist<br><u>Setting</u><br>Secondary/Tertiary care – social care<br>Geriatric Clinic for specialized care located in a University hospital; Geriatric Clinic had four wards, one of which was located in the community | <u>Sample size</u><br>n=32<br>Occupational therapist (n=3)<br>Dietitian (n=1)<br>Social worker (n=3)<br>Physician (n=8)<br>Physiotherapist (n=4)<br>Registered nurse (n=7)<br>Assistant nurse (n=6)<br><u>Condition</u><br>Aging associated diseases (medical disorders, orthopaedic post-surgery care, stroke, palliative care)<br><u>Participant details</u><br>HCPs<br>*Ethnicity:* Not reported<br>*Gender:* Not reported<br>*Age:* Not reported | <u>Quality rating</u><br>7 out of 10 criteria met on JBI critical appraisal checklist for qualitative research |
| Chow et al. 2015<br>(Australia)<br><u>Aim</u><br>To evaluate the effectiveness of the Triple I (Hub), an integrated intake, information, and intervention service, in co-locating six previously disparate services to determine its impact on service | <u>Study design</u><br>Mixed methods (only the qualitative arm is extracted)<br><u>Data collection methods</u><br>Site visit: Clinical observations by external expert<br>-Examination of records<br>-Informal discussions with staff members<br><u>Dates of data collection</u><br>May 2013 (six months after the opening of the Triple I (Hub)) | <u>Type of intervention</u><br>Six pre-existing services were relocated to be co-located within the same premises of the South West Sydney Medicare Local:<br>1. Community Health Nursing (formerly CHAIN)<br>2. Aged Care & Rehabilitation Services Referral and Information Centre's (RIC South)<br>3. Aged Care & Rehabilitation Services (RIC Central) | <u>Sample size</u><br>n=Not reported<br><u>Condition</u><br>Aged care<br><u>Participant details</u><br>HCPs<br>*Ethnicity:* Not reported<br>*Gender:* Not reported<br>*Age:* Not reported | <u>Quality rating</u><br>3 out of 10 criteria met on JBI critical appraisal checklist for qualitative research |
| delivery, efficiency and patient satisfaction five months after implementation. |  | 4. Aged Care & Chronic Care Triage (formerly ACCT)<br>5. Community Palliative Care<br>6. ComPacks (post-hospitalisation support)<br>Service delivery model:<br>Designed to empower clients towards self-management, informed decision-making and exercise their right to choose to exit services<br>From one intake points, multiple referrals to different service providers are made through a single contact point<br>Processes were formalized for case managers to work collaboratively with primary care practitioners to:<br>(1) Integrate care through linking up the clients with all required services<br>(2) Provide individualized information<br>(3) Facilitate assessment and care planning within a multidisciplinary team<br>(4) Ensure general practitioner engagement during the transition of care of clients<br><u>Intervention providers</u><br>Primary care practitioners, case managers, community nursing<br><u>Setting</u><br>Primary care – Community care |  |  |
| Fox et al. 2023 (UK)<br><u>Aim</u> | <u>Study design</u><br>Qualitative descriptive<br><u>Data collection methods</u> | <u>Type of intervention</u><br>*Hip fracture care pathway* involves multiple hospital departments and teams, spanning admission to | <u>Sample size</u><br>n=40 | <u>Quality rating</u><br>6 out of 10 criteria met on JBI critical appraisal checklist for qualitative research |
| To investigate the organisational processes that help and hinder the implementation of hip fracture services<br>To identify potentially modifiable barriers and facilitators to care delivery, to inform recommendations to improve care delivery, assist service improvements, and ultimately reduce health inequities and improve patient outcomes. | Semi-structured interviews (telephone and Microsoft Teams; Documentary analysis of anonymised British Orthopaedic Association hospital reports addressing under-performing hip fracture services (n=23)<br><u>Dates of data collection</u><br>*Interviews:* during the COVID-19 pandemic<br>*Documentary analysis:* 2012 - 2019 | discharge. National guidelines have tried to standardise key components of this pathway. Standards include admitting patients to an acute hip fracture ward; rapid optimisation of fitness for surgery; time-specific targets for surgery and first day post-operative mobilisation. Guidelines advocate continued, coordinated, orthogeriatric and multidisciplinary review, with the aim for patients to recover independence and return to pre-fracture residence. Adherence to these standards requires a multidisciplinary team who consistently communicate, collaborate, understand the care pathway and are responsive to change.<br><u>Intervention providers</u><br>A wide range of professionals were interviewed per hospital, the level of involvement with the care pathway is unclear<br><u>Setting</u><br>Secondary – Community care<br>*Interviews:* Four hospitals (three urban, one rural)<br>*Documentary analysis:* 23 hospitals | Consultant geriatrician / Orthogeriatrician n=7<br>ED Consultant n=1<br>Band 8 Advanced Nurse Practitioner ED n=1<br>Anaesthetist n=6<br>Orthopaedic surgeon n=7<br>Orthogeriatric Advanced Nurse Specialist n=1<br>Physiotherapist n=2<br>Occupational Therapist n=4<br>Musculoskeletal Matron n=1<br>Service Manager n=2<br>Ward Manager n=1<br>Senior Theatre Practitioner n=1<br>Advanced Nurse Practitioner n=2<br>Orthopaedic Registrar n=1<br>Foundation Year 2 Doctor n=1<br>Discharge coordinator n=1<br>Trauma coordinator n=1<br><u>Condition</u><br>Hip fracture<br><u>Participant details</u><br>*HCPs*<br>*Ethnicity:* Not reported<br>*Gender:* Not reported<br>*Age:* Not reported |  |
| Greene et al. 2023 (Australia)<br><u>Aim</u><br>To evaluate a new ED avoidance service (the Complex And Restorative (CARE) for | <u>Study design</u><br>Qualitative descriptive (Part of a wider service evaluation)<br><u>Data collection methods</u><br>Semi-structured interviews | <u>Type of intervention</u><br>Complex And Restorative (CARE) Centre, an emergency department avoidance service for older people requiring urgent but non-emergency care<br><u>Intervention providers</u> | <u>Sample size</u><br>Patients n=17<br>Relatives n=15<br>Participants reported on 32 attendances to the urgent CARE centre<br><u>Condition</u><br>Urgent but non-emergency | <u>Quality rating</u><br>7 out of 10 criteria met on JBI critical appraisal checklist for qualitative research |
| older people requiring urgent care | <u>Dates of data collection</u><br>September 2021 - September 2022 | Geriatrician, registered nurses, enrolled nurses, physiotherapists, occupational therapists, social workers<br>Paramedic performs triage based on CARE inclusion criteria<br><u>Setting</u><br>Secondary/Tertiary care - social care<br>600-bed tertiary hospital | Most presentations to the CARE centre were for acute symptoms, primarily falls, headaches, general pain, high blood pressure, chest pain, shortness of breath, and panic attacks<br>Some presentations were associated with chronic conditions such as chronic obstructive pulmonary disease, diverticulitis, and Hodgkin's lymphoma<br><u>Participant details</u><br>*Patients*<br>*Ethnicity:* None identified as Aboriginal or Torres Strait Islander<br>*Gender:* Females (65%)<br>*Age:* Mean years 83. 2<br>Range: 71-93 years<br>*Relatives*<br>*Ethnicity:* None identified as Aboriginal or Torres Strait Islander<br>*Gender:* Female (73%) (nine daughters, four sons and two wives)<br>*Age:* Not reported |  |
| Lee et al. 2015 (Canada)<br><u>Aim</u><br>To explore the barriers to and facilitators of adapting and expanding a primary care memory clinic model to integrate care of additional complex chronic geriatric conditions (heart failure, falls, chronic obstructive pulmonary disease, and frailty) into care processes with the | <u>Study design</u><br>Mixed methods (Only the qualitative arm is extracted)<br><u>Data collection methods</u><br>Structured interviews<br><u>Dates of data collection</u><br>February 2013 - March 2013 | <u>Type of intervention</u><br>Primary care-based interprofessional memory clinic model designed to enhance chronic disease management for elderly patients with complex medical conditions<br><u>Intervention providers</u><br>Family physicians, nurses, social workers<br>Geriatric specialists linked to clinics<br><u>Additional specialist in some clinics</u><br>Pharmacists, occupational therapists, representatives from local Alzheimer's Society | <u>Sample size</u><br>n=16<br>Primary (memory) clinic lead physicians n=8<br>Memory clinic co-lead physicians n=8<br><u>Condition</u><br>Complex chronic geriatric conditions (dementia, heart failure, falls, chronic obstructive pulmonary disease, frailty and multimorbidity)<br><u>Participant details</u><br>*Primary clinic lead physicians*<br>*Ethnicity:* Not reported<br>*Gender:* Not reported | <u>Quality rating</u><br>6 out of 10 criteria met on JBI critical appraisal checklist for qualitative research |
| goal of improving outcomes for seniors |  | <u>Setting</u><br>Primary care - Secondary/Tertiary care – community care - social care<br>Centre for Family Medicine, Family Health Team Memory Clinic | Age: Not reported<br>*Memory clinic co-lead physicians*<br>*Ethnicity: Not reported*<br>*Gender: Not reported*<br>*Age: Not reported* |  |
| Lee et al. 2017 (Canada)<br><u>Aim</u><br>To describe the significance of a primary care collaborative memory clinic model of care within the system of care for older adults with dementia in Ontario | <u>Study design</u><br>Descriptive survey with open-ended responses<br><u>Data collection methods</u><br>Post program descriptive survey with open-ended responses in which participants were asked to identify at least one way in which patients or family members have benefited<br><u>Dates of data collection</u><br>Individuals attending 15 Primary care collaborative memory clinic model of care training program sessions between October 2008 to June 2014 were invited to participate | <u>Type of intervention</u><br>Primary care collaborative memory clinic model of care – multidisciplinary clinics<br><u>Intervention providers</u><br>Family physician, nurse (registered nurses, registered practical nurses, nurse practitioner)<br>Geriatricians and/or geriatric psychiatrists linked to clinics<br>Social worker, pharmacists, occupational therapists, other allied health professionals (as available)<br>*Additional specialist in some clinics*<br>Representatives from local Alzheimer's Society,<br><u>Setting</u><br>Primary care - Secondary/tertiary care – community care - social care<br>46 primary care settings (Family health centres (n=41), community health centres (n=5)) | <u>Sample size</u><br>n=198/364 (54.4%) (post program survey)<br>Intervention providers who had completed training<br><u>Condition</u><br>Dementia<br><u>Participant details</u><br>*Ethnicity: Not reported*<br>*Gender: Not reported*<br>*Age: Not reported* | <u>Quality rating</u><br>4 out of 10 criteria met on JBI critical appraisal checklist for qualitative research |
| Luckett et al. 2017 (Australia)<br><u>Aim</u><br>To explore palliative care planning | <u>Study design</u><br>Qualitative descriptive (Part of a wider cluster RCT)<br><u>Data collection methods</u><br>Semi-structured interviews | <u>Type of intervention</u><br>Facilitated case conferencing<br>-Multidisciplinary input into case conferences<br>-Care planning | <u>Sample size</u><br>n=40<br>Palliative care planning coordinators n=11<br>Nursing home staff n=18 | <u>Quality rating</u><br>6 out of 10 criteria met on JBI critical appraisal checklist for qualitative research |
| coordinators and health professional perceptions of the benefits of facilitated case conferencing and identify factors influencing implementation | <u>Dates of data collection</u><br>Not reported | <u>Intervention providers</u><br>Palliative care planning coordinators, nursing home staff, Physicians (GP, Medical Officer, Geriatrician), Physiotherapist or aides, Diversional therapists, Dietician<br><u>Setting</u><br>Primary care – community care – social care<br>Nursing homes (n=10) | Physiotherapists or aides n=3<br>Diversional therapist n=4<br>Dietitian n=1<br>GP n=1<br>Medical Officer n=1<br>Geriatrician n=1<br><u>Condition</u><br>Dementia (advanced)<br><u>Participant details</u><br>*Ethnicity:* Not reported<br>*Gender:* Not reported<br>*Age:* Not reported |  |

**Key:** AHPs – allied health professionals, ED – emergency department, GP – General practitioner, RCT – randomised controlled trial

### 6.3 Quality appraisal

**Table 10:**
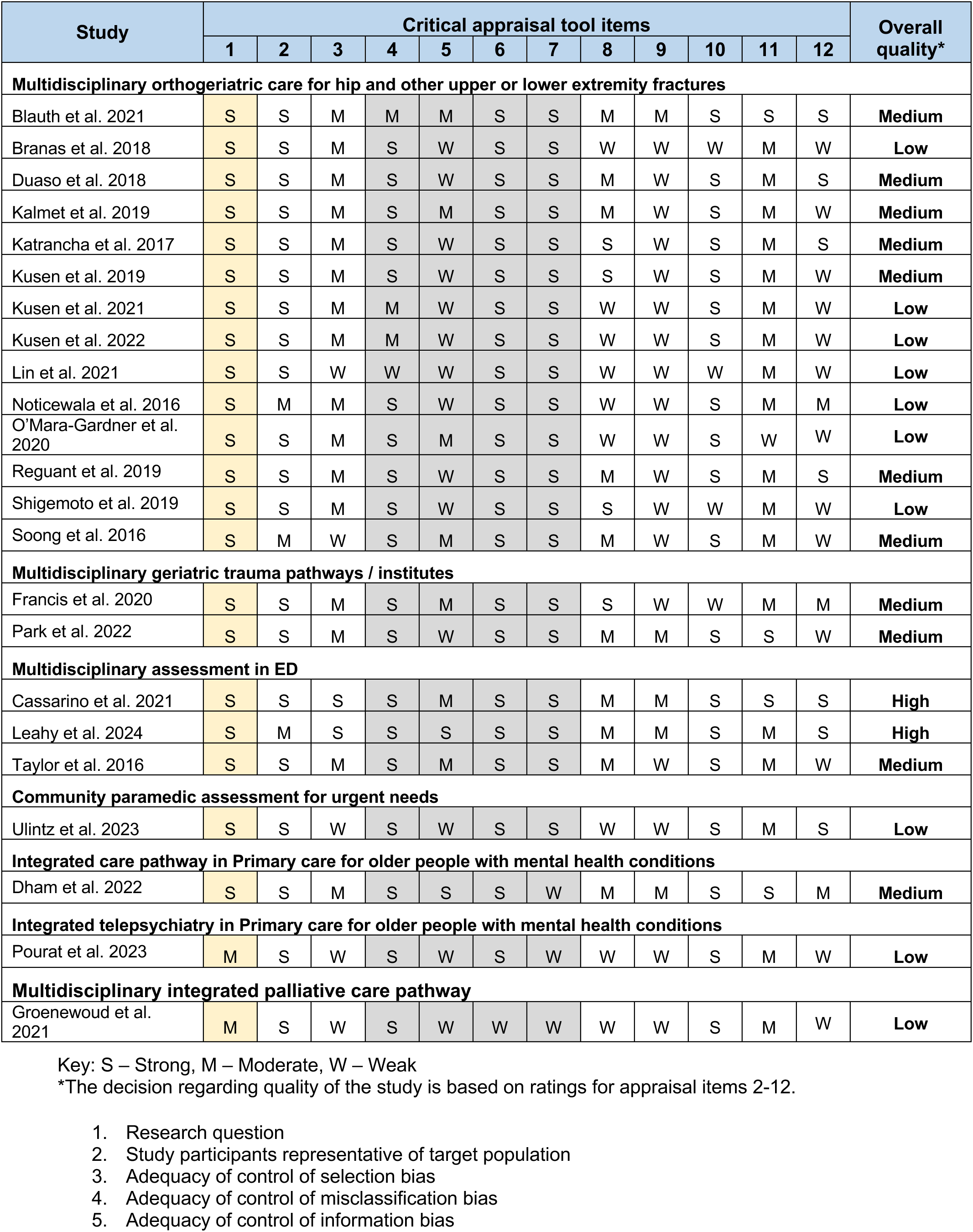
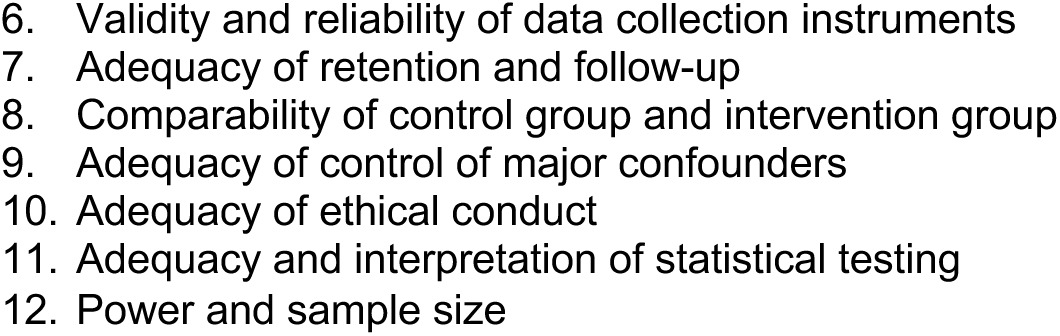
Critical appraisal tool – Analytic study.

| Study | Critical appraisal tool items |  |  |  |  |  |  |  |  |  |  |  | Overall quality* |
| --- | --- | --- | --- | --- | --- | --- | --- | --- | --- | --- | --- | --- | --- |
|  | 1 | 2 | 3 | 4 | 5 | 6 | 7 | 8 | 9 | 10 | 11 | 12 |  |
| Multidisciplinary orthogeriatric care for hip and other upper or lower extremity fractures |  |  |  |  |  |  |  |  |  |  |  |  |  |
| Blauth et al. 2021 | S | S | M | M | M | S | S | M | M | S | S | S | Medium |
| Branas et al. 2018 | S | S | M | S | W | S | S | W | W | W | M | W | Low |
| Duaso et al. 2018 | S | S | M | S | W | S | S | M | W | S | M | S | Medium |
| Kalmet et al. 2019 | S | S | M | S | M | S | S | M | W | S | M | W | Medium |
| Katrancha et al. 2017 | S | S | M | S | W | S | S | S | W | S | M | S | Medium |
| Kusen et al. 2019 | S | S | M | S | W | S | S | S | W | S | M | W | Medium |
| Kusen et al. 2021 | S | S | M | M | W | S | S | W | W | S | M | W | Low |
| Kusen et al. 2022 | S | S | M | M | W | S | S | W | W | S | M | W | Low |
| Lin et al. 2021 | S | S | W | W | W | S | S | W | W | W | M | W | Low |
| Noticewala et al. 2016 | S | M | M | S | W | S | S | W | W | S | M | M | Low |
| O'Mara-Gardner et al. 2020 | S | S | M | S | M | S | S | W | W | S | W | W | Low |
| Reguant et al. 2019 | S | S | M | S | W | S | S | M | W | S | M | S | Medium |
| Shigemoto et al. 2019 | S | S | M | S | W | S | S | S | W | W | M | W | Low |
| Soong et al. 2016 | S | M | W | S | M | S | S | M | W | S | M | W | Medium |
| Multidisciplinary geriatric trauma pathways / institutes |  |  |  |  |  |  |  |  |  |  |  |  |  |
| Francis et al. 2020 | S | S | M | S | M | S | S | S | W | W | M | M | Medium |
| Park et al. 2022 | S | S | M | S | W | S | S | M | M | S | S | W | Medium |
| Multidisciplinary assessment in ED |  |  |  |  |  |  |  |  |  |  |  |  |  |
| Cassarino et al. 2021 | S | S | S | S | M | S | S | M | M | S | S | S | High |
| Leahy et al. 2024 | S | M | S | S | S | S | S | M | M | S | M | S | High |
| Taylor et al. 2016 | S | S | M | S | M | S | S | M | W | S | M | W | Medium |
| Community paramedic assessment for urgent needs |  |  |  |  |  |  |  |  |  |  |  |  |  |
| Ulintz et al. 2023 | S | S | W | S | W | S | S | W | W | S | M | S | Low |
| Integrated care pathway in Primary care for older people with mental health conditions |  |  |  |  |  |  |  |  |  |  |  |  |  |
| Dham et al. 2022 | S | S | M | S | S | S | W | M | M | S | S | M | Medium |
| Integrated telepsychiatry in Primary care for older people with mental health conditions |  |  |  |  |  |  |  |  |  |  |  |  |  |
| Pourat et al. 2023 | M | S | W | S | W | S | W | W | W | S | M | W | Low |
| Multidisciplinary integrated palliative care pathway |  |  |  |  |  |  |  |  |  |  |  |  |  |
| Groenewoud et al. 2021 | M | S | W | S | W | W | W | W | W | S | M | W | Low |

Key: S – Strong, M – Moderate, W – Weak
\*The decision regarding quality of the study is based on ratings for appraisal items 2-12.
1. Research question 2. Study participants representative of target population 3. Adequacy of control of selection bias 4. Adequacy of control of misclassification bias 5. Adequacy of control of information bias
6. Validity and reliability of data collection instruments 7. Adequacy of retention and follow-up 8. Comparability of control group and intervention group 9. Adequacy of control of major confounders 10. Adequacy of ethical conduct 11. Adequacy and interpretation of statistical testing 12. Power and sample size

**Table 11:**
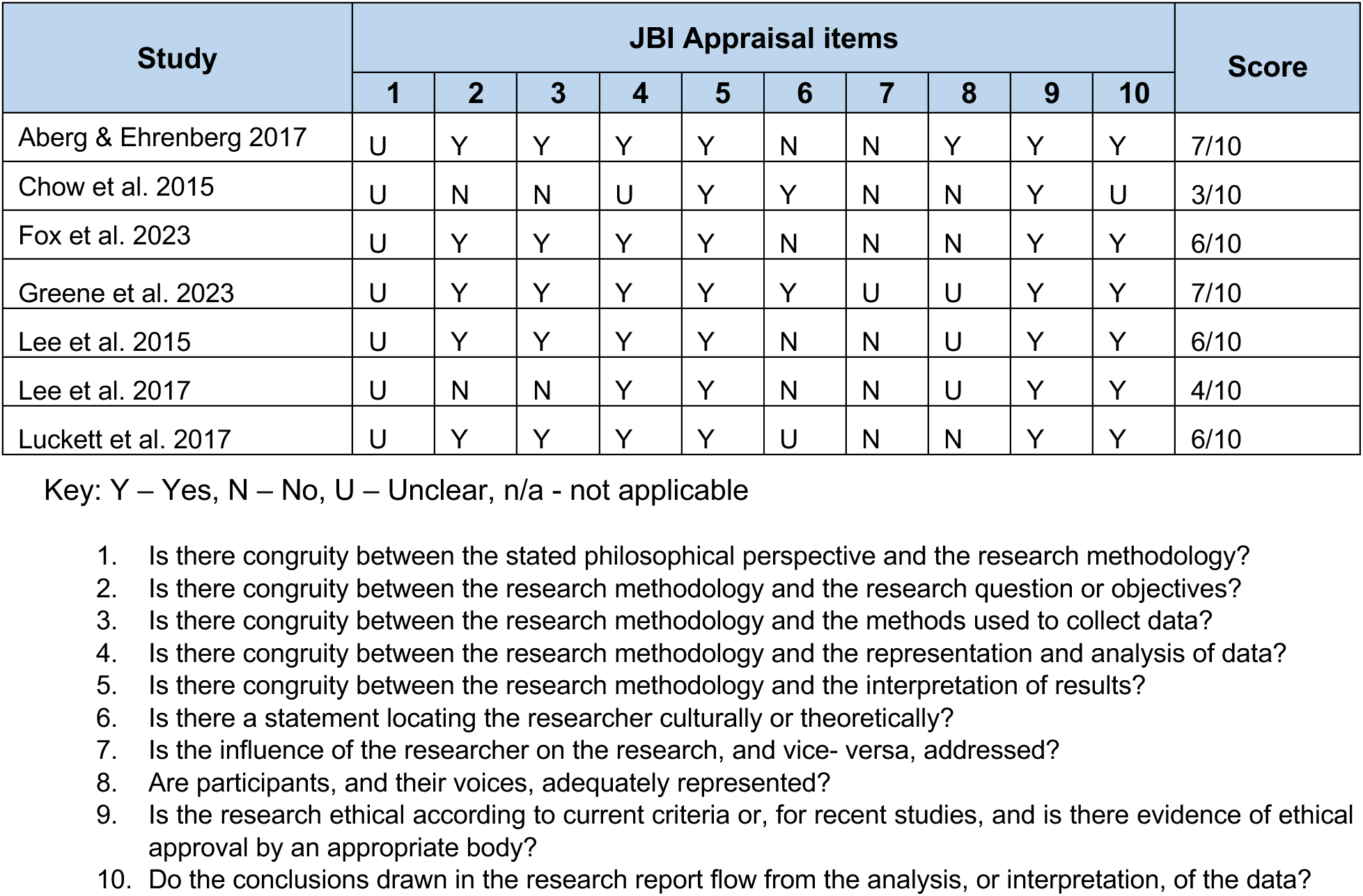
JBI critical appraisal checklist for qualitative research.

| Study | JBI Appraisal items |  |  |  |  |  |  |  |  |  | Score |
| --- | --- | --- | --- | --- | --- | --- | --- | --- | --- | --- | --- |
|  | 1 | 2 | 3 | 4 | 5 | 6 | 7 | 8 | 9 | 10 |  |
| Aberg & Ehrenberg 2017 | U | Y | Y | Y | Y | N | N | Y | Y | Y | 7/10 |
| Chow et al. 2015 | U | N | N | U | Y | Y | N | N | Y | U | 3/10 |
| Fox et al. 2023 | U | Y | Y | Y | Y | N | N | N | Y | Y | 6/10 |
| Greene et al. 2023 | U | Y | Y | Y | Y | Y | U | U | Y | Y | 7/10 |
| Lee et al. 2015 | U | Y | Y | Y | Y | N | N | U | Y | Y | 6/10 |
| Lee et al. 2017 | U | N | N | Y | Y | N | N | U | Y | Y | 4/10 |
| Luckett et al. 2017 | U | Y | Y | Y | Y | U | N | N | Y | Y | 6/10 |

Key: Y – Yes, N – No, U – Unclear, n/a - not applicable
1. Is there congruity between the stated philosophical perspective and the research methodology? 2. Is there congruity between the research methodology and the research question or objectives? 3. Is there congruity between the research methodology and the methods used to collect data? 4. Is there congruity between the research methodology and the representation and analysis of data? 5. Is there congruity between the research methodology and the interpretation of results? 6. Is there a statement locating the researcher culturally or theoretically? 7. Is the influence of the researcher on the research, and vice- versa, addressed? 8. Are participants, and their voices, adequately represented? 9. Is the research ethical according to current criteria or, for recent studies, and is there evidence of ethical approval by an appropriate body? 10. Do the conclusions drawn in the research report flow from the analysis, or interpretation, of the data?

### 6.4 Information available on request

The protocol is available online: https://doi.org/10.17605/OSF.IO/3SD4F

Search strategies and list of excluded studies is presented below in the Appendices.

## ADDITIONAL INFORMATION

### 7.1 Conflicts of interest

The authors declare they have no conflicts of interest to report.

## Acknowledgements

The authors would like to thank Tom Cushion, Helen Howson, Leo Lewis and Robert Hall for their time, expertise, and contributions during stakeholder meetings in guiding the focus of the review and interpretation of findings. We would also like to thank Alexander Bridgman for supporting our stakeholder meeting.

## APPENDICES

## APPENDIX 1: Search strategies

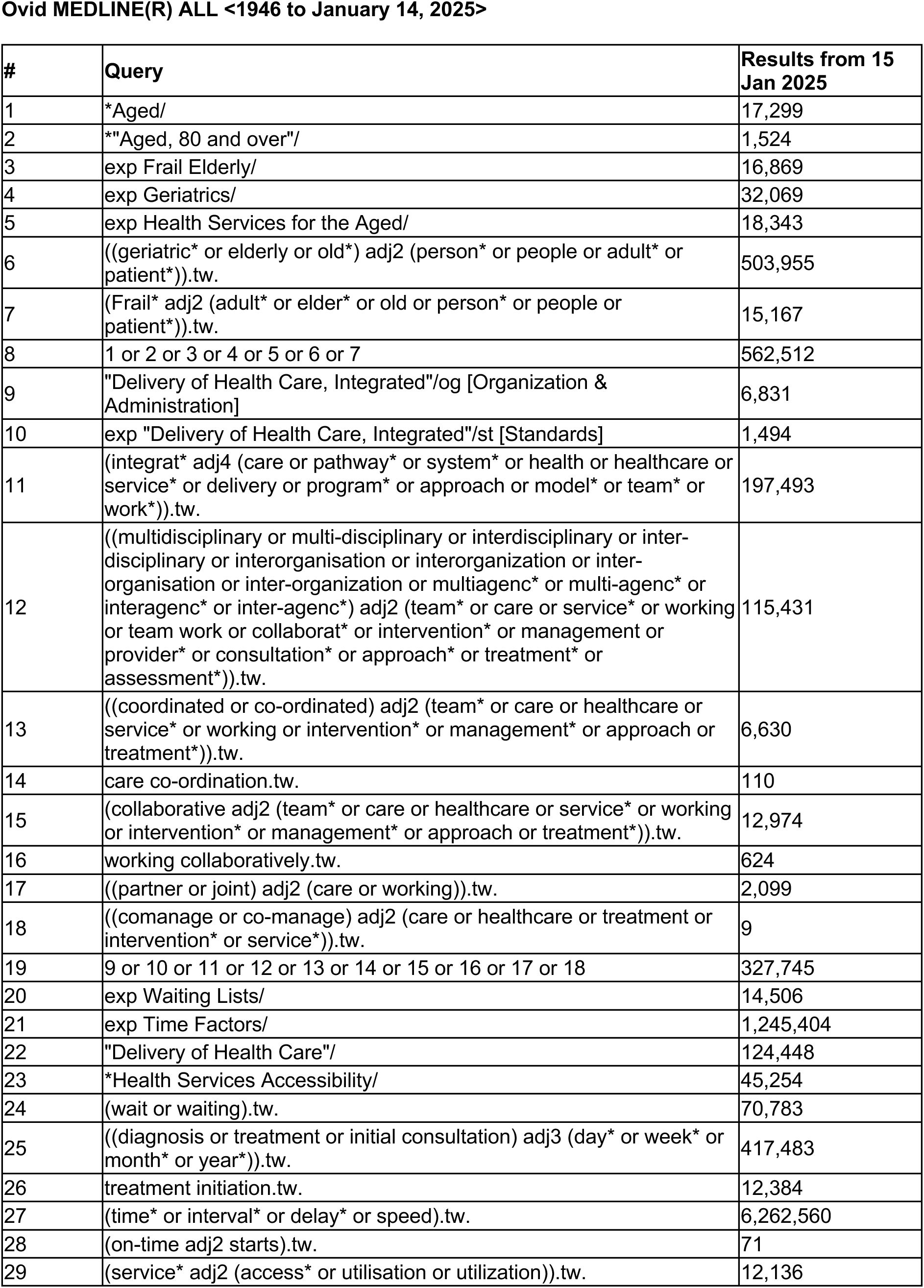
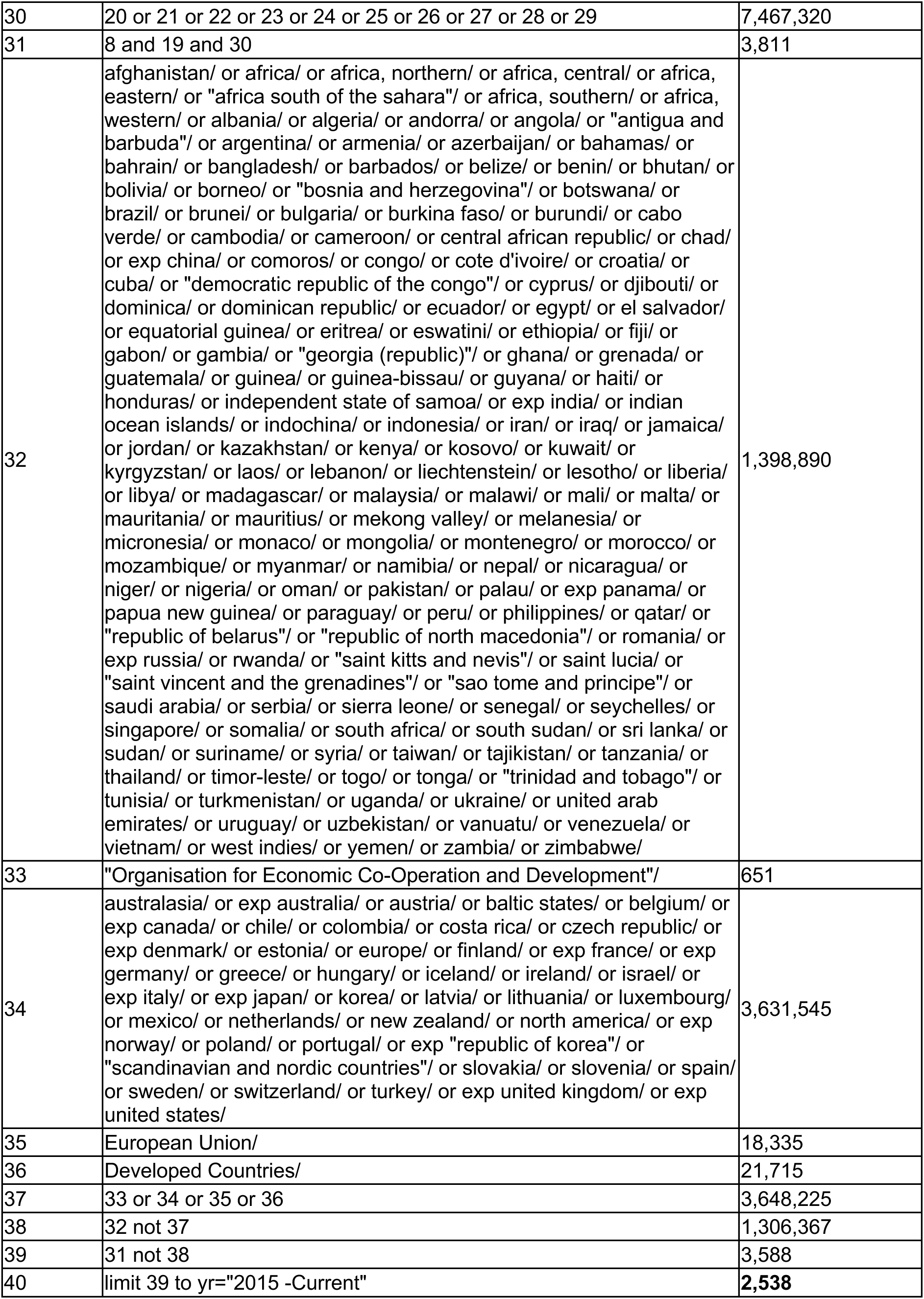
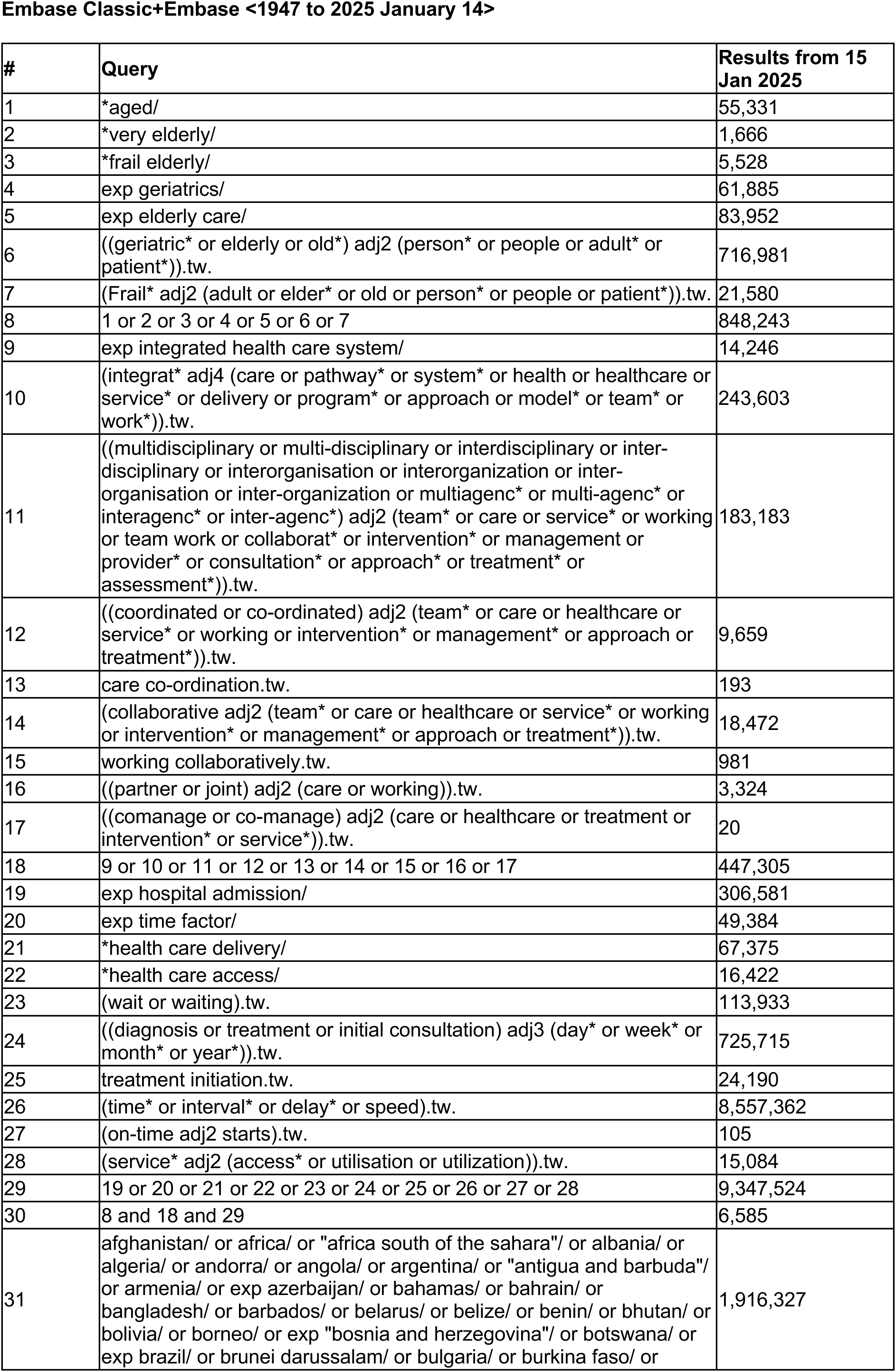
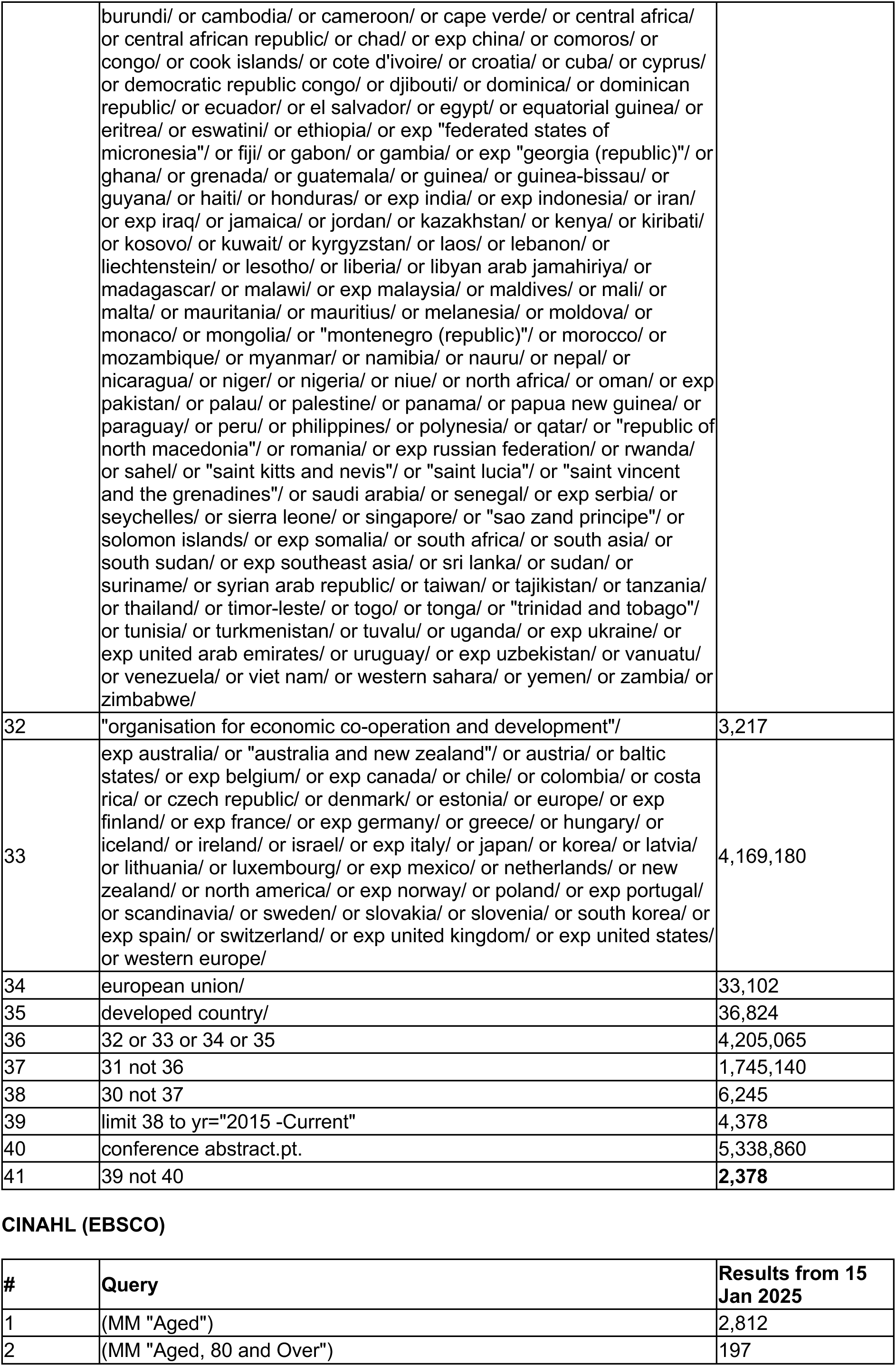
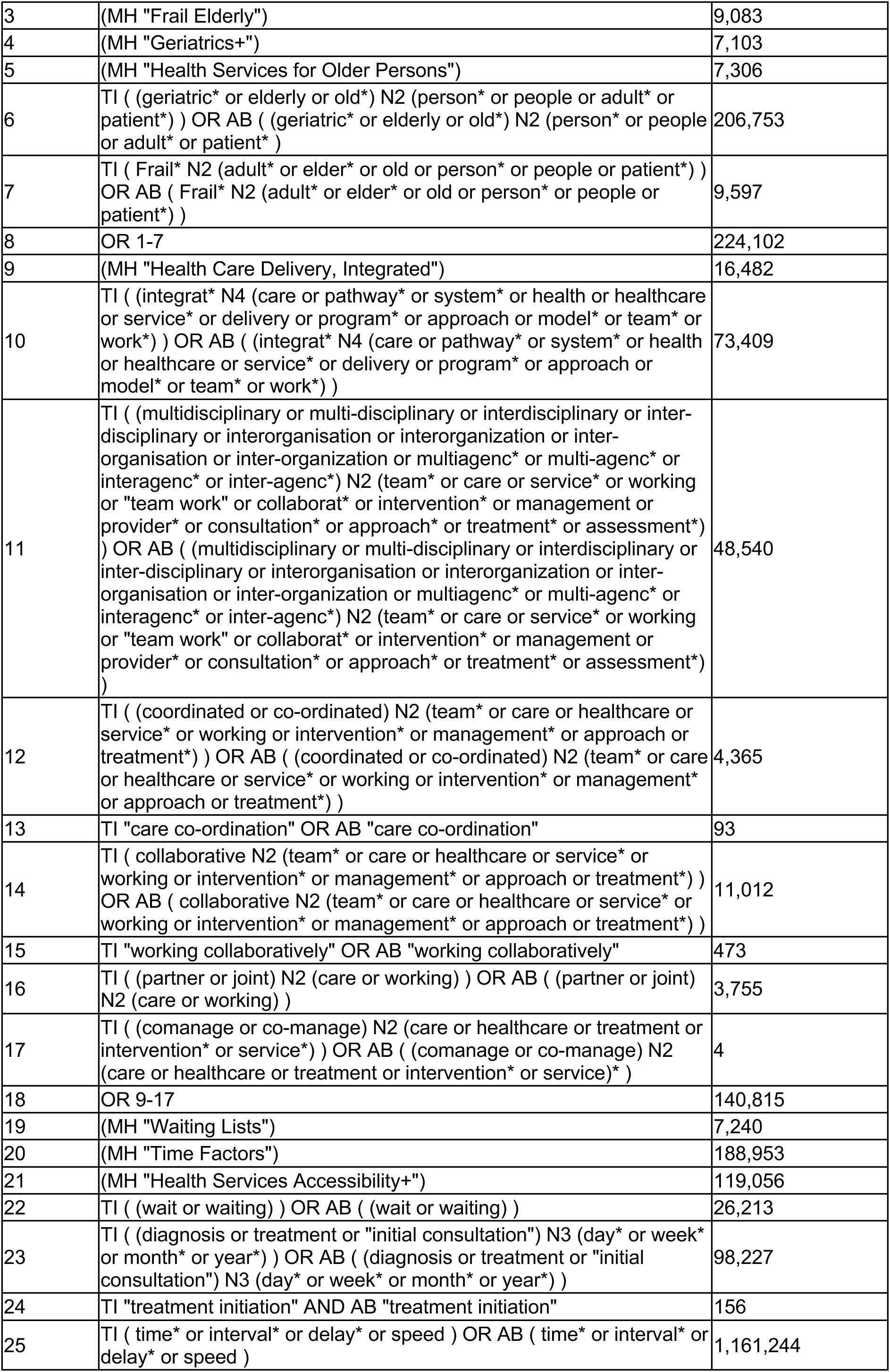
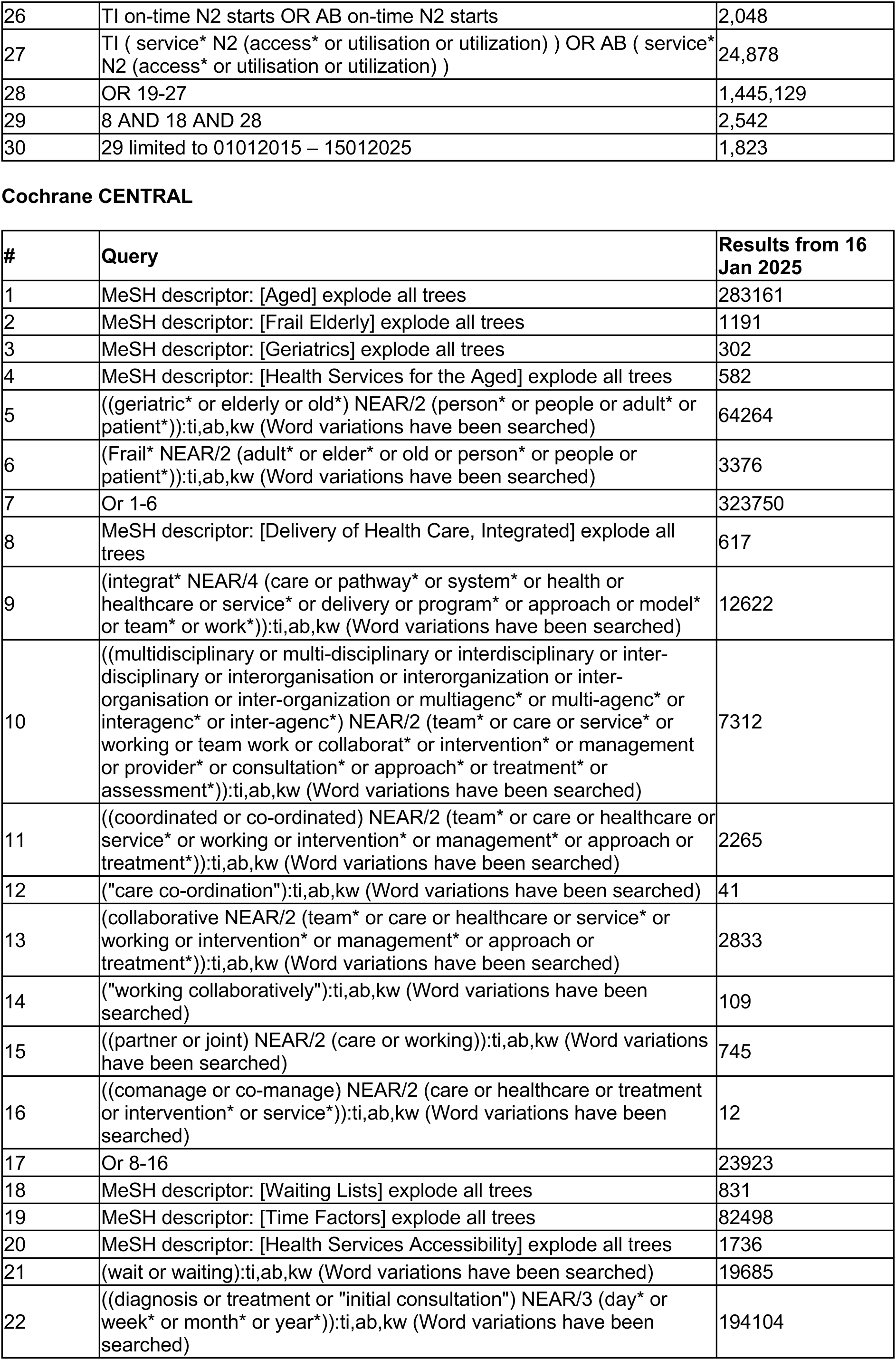
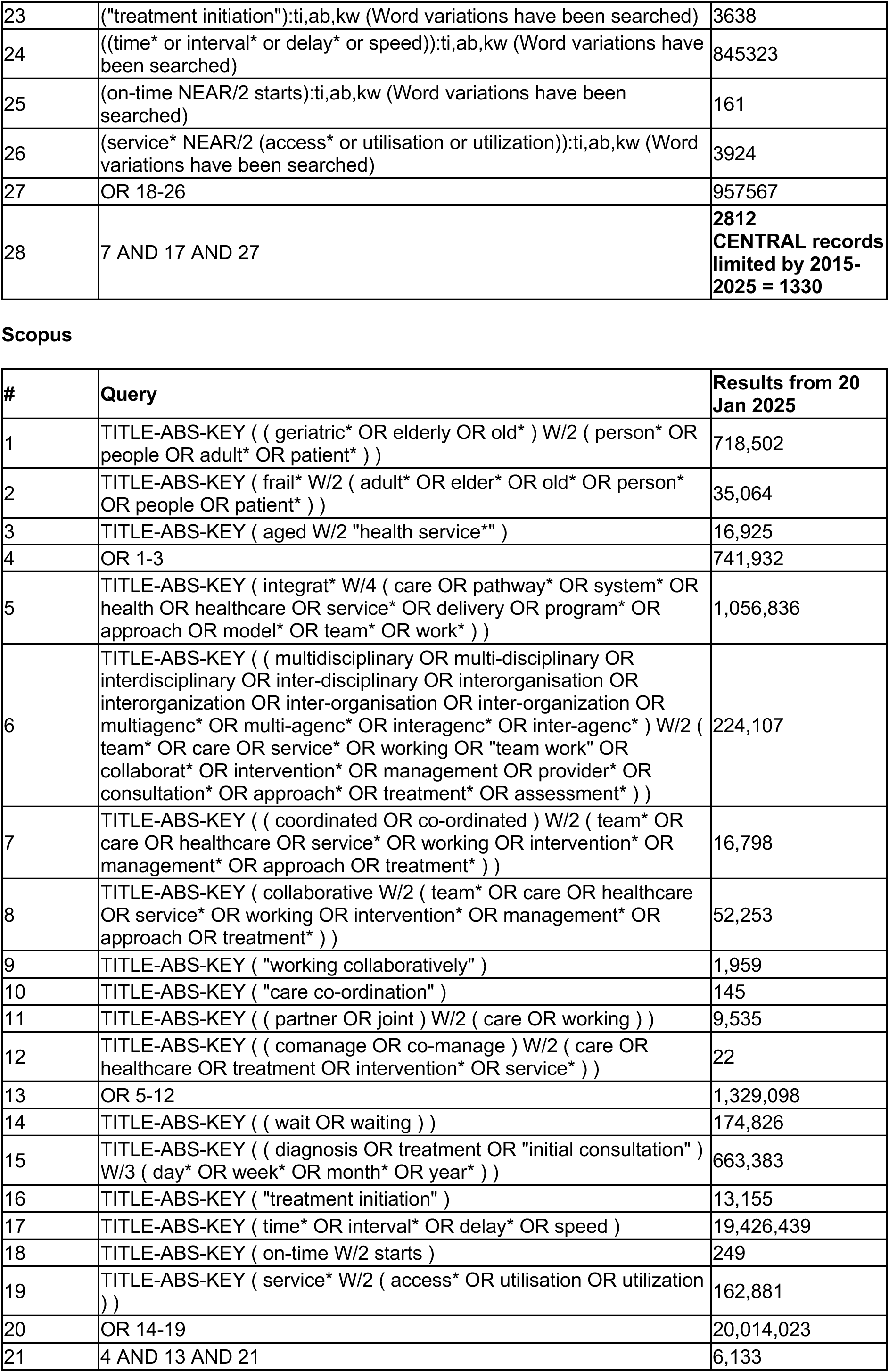
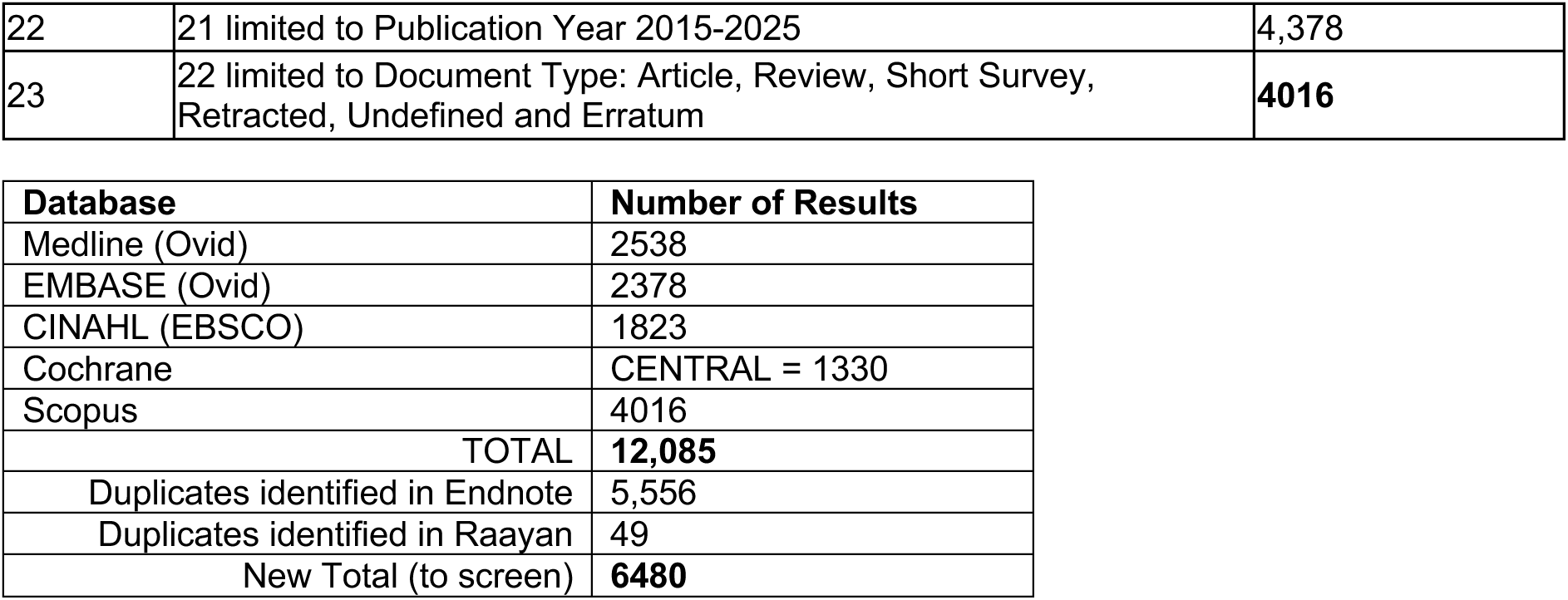

## APPENDIX 2: List of grey literature sources

| Source |
| --- |
| NHS England<br>https://www.england.nhs.uk/ |
| NHS Wales<br>https://www.wales.nhs.uk/ |
| NHS Scotland<br>https://www.careers.nhs.scot/careers/find-your-career/returning-to-practice/ |
| Health and Social Care Northern Ireland<br>https://www.hscni.net/ |
| Welsh Government<br>https://gov.wales/ |
| UK Government<br>https://www.gov.uk/ |
| Scottish Government<br>https://www.gov.scot/ |
| Northern Ireland Executive<br>https://www.northernireland.gov.uk/ |
| National Institute for Health and Care Excellence (NICE)<br>https://www.nice.org.uk/ |
| Public Health Wales<br>https://phw.nhs.wales/ |
| UK Health Security Agency<br>https://researchportal.ukhsa.gov.uk/ |
| Office for Health Improvement & Disparities<br>https://www.gov.uk/government/organisations/office-for-health-improvement-and-disparities |
| Public Health Scotland<br>https://publichealthscotland.scot/ |
| Public Health Northern Ireland<br>https://www.publichealth.hscni.net/ |
| Health Foundation<br>https://www.health.org.uk/ |
| Nuffield Trust<br>https://www.nuffieldtrust.org.uk/ |
| King's Fund<br>https://www.kingsfund.org.uk/ |
| Age UK<br>https://www.ageuk.org.uk/ |
| Age Cymru<br>https://www.agecymru.wales/ |
| Older People's Commissioner for Wales<br>https://olderpeople.wales/ |
| British Geriatric Society<br>https://www.bgs.org.uk/ |
| Social Care Institute for Excellence (SCIE)<br>https://www.scie.org.uk/ |
| Personal Social Services Research Unit (PSSRU)<br>https://www.pssru.ac.uk/ |
| Care Quality Commission<br>https://www.cqc.org.uk/ |
| Care Inspectorate Wales<br>https://careinspectorate.wales/ |
| Skills for Care<br>https://www.skillsforcare.org.uk/Recruitment-retention/Recruitment-and-retention.aspx |
| Local Government Association<br>https://www.local.gov.uk/ |
| Association of Directors of Adult Social Services (ADASS) |
| https://www.adass.org.uk/publications2 |
| Care and Support Alliance<br>https://careandsupportalliance.com/ |
| The Care Provider Alliance<br>https://careprovideralliance.org.uk/ |
| Think Local Act Personal<br>https://www.thinklocalactpersonal.org.uk/Latest/?s=337 |
| National Audit Office<br>https://www.nao.org.uk/search/type/report/ |
| The Scottish Social Services Council<br>https://www.sssc.uk.com/ |
| Social Care Wales<br>https://socialcare.wales |
| Social Work England<br>https://www.socialworkengland.org.uk/ |
| Northern Ireland Social Care Council<br>https://niscc.info/ |
| Department of Health & Social Care<br>https://www.gov.uk/government/organisations/department-of-health-and-social-care |
| British Association of Social Work<br>https://www.basw.co.uk/resources/publications-policies-and-reports |
| NHS Improvement<br>https://www.england.nhs.uk/publication/ |
| Health Education England<br>https://www.hee.nhs.uk/ |
| The National Care Forum<br>https://www.nationalcareforum.org.uk/ |
| Academy of Fabulous Stuff<br>https://fabnhsstuff.net/ |
| Royal College of Nursing<br>https://www.rcn.org.uk/ |
| NHS Confederation<br>https://www.nhsconfed.org/ |
| Bevan Commission<br>https://bevancommission.org/ |
| NESTA<br>https://www.nesta.org.uk/ |

## APPENDIX 3: List of excluded studies

| N o | Citation | Reason |
| --- | --- | --- |
| **Bibliographic database searches** |  |  |
| 1. | Adersen M, Chen, I M, Rasmussen L S, Johansen J S, Nissen M, Groenvold M and Marsaa, K. (2021). Regional and age differences in specialised palliative care for patients with pancreatic cancer. *BMC Palliative Care*. https://doi.org/10.1186/s12904-021-00870-8 | Wrong population |
| 2. | Alexander K, Hamlin P A, Tew W P, Trevino K, Tin A L, Shahrokni A, Meditz E, Boparai M, Amirnia F, Sun, S W and Korc-Grodzicki, B. (2023). Development and implementation of an interdisciplinary telemedicine clinic for older patients with cancer- Preliminary data. *Journal of the American Geriatrics Society* https://dx.doi.org/10.1111/jgs.18267 | Wrong design |
| 3. | Amjad H, Wong S K, Roth D L, Huang J, Willink A, Black B S, Johnston D, Rabins P V, Gitlin L N, Lyketsos C G et al. (2018). Health Services Utilization in Older Adults with Dementia Receiving Care Coordination: the MIND at Home Trial. *Health services research*. https://doi.org/10.1111/1475-6773.12647 | Wrong outcome |
| 4. | Beech C and Verity F (2020). Health and social work practitioners' experiences of working with risk and older people: The interconnectedness of personalities, process and policy. *Journal of Integrated Care* https://doi.org/10.1108/jica-08-2019-0036 | Wrong outcome |
| 5. | Bommireddy L, Leow T W, Gogna R, and Clark, D I. (2021). Should Femoral Shaft fractures in Patients Age Over 60 Years be Managed Using a Hip Fracture Pathway? *Injury* https://dx.doi.org/10.1016/j.injury.2020.09.059 | Wrong design |
| 6. | Choy C H, Steeds R P, Pinney J, Baig S, Turvey-Haigh L, Wahid Y, Cox H, Zaphiriou A, Srinivasan V, Wilson D et al. (2024). Extending the reach of expert amyloidosis care: A feasibility study exploring the staged implementation of a UK amyloidosis network. *Clinical Medicine, Journal of the Royal College of Physicians of London* https://doi.org/10.1016/j.clinme.2023.100004 | Wrong design |
| 7. | Clancy U, Brown M, Alio Z, Wardle K, and Pendleton, Neil (2018). Older people with hip fracture transferred to intermediate care: outcomes in an integrated health and social care model. *Future healthcare journal* https://dx.doi.org/10.7861/futurehosp.5-1-58 | Wrong design |
| 8. | Congedo M T, Nachira D, Pennisi M A, Chiappetta M, Calabrese G, Bello G et al. (2022). Risk Factors Associated with Post-Operative Complications in Multidisciplinary Treatment of Descending Necrotizing Mediastinitis. *Journal of clinical medicine* https://dx.doi.org/10.3390/jcm11216364 | Wrong outcome |
| 9. | Corcoran G, Gavaghan G, Lyons C, and Moloney, E. (2017). Timely identification of frailty & comprehensive multidisciplinary assessment on a newly established specialist geriatric ward. *International Journal of Integrated Care (IJIC)*. https://doi.org/10.5334/ijic.3668 | Conference abstract |
| 10. | Crego-Vita D, Sanchez-Perez C, Gomez-Rico J A O, De Arriba C and Clemente de Arriba C. (2017). Intracapsular hip fractures in the elderly. Do we know what is important? *Injury*. https://dx.doi.org/10.1016/j.injury.2017.01.022 | Wrong design |
| 11. | De Belvis A G, Bocci M G, Morsella A, Balducci F M, Loconsole L and Angioletti C, et al. 1(2020). Major trauma critical pathway: preliminary results from the monitoring system in the regional network and in a hub center in Rome metropolitan area. *European review for medical and pharmacological sciences*. https://doi.org/10.26355/eurrev_202007_21878 | Wrong population |
| 12. | DelaCruz J J, Giannikos C, Kakolyris A, Utzinger R C, and Karpiak, S E. (2021). Cost-Effectiveness Analysis Combining Medical and Mental Health Services for Older Adults with HIV in New York City. *Atlantic economic journal : AEJ*. https://dx.doi.org/10.1007/s11293-021-09697-3 | Wrong outcome |
| 13. | Elliott J, Koch M, McDermott M, Sacco V, and Stolee P. (2022). Developing a Regional Strategy for Older Adults Living With Frailty: Recommendations From Patients, Family Caregivers and Health Care Providers. *International Journal of Integrated Care (IJIC)*. https://doi.org/10.5334/ijic.6438 | Wrong intervention |
| 14. | Elrashidi M Y, Philpot L M, Young N P, Ramar P, Swanson K M and McKie P M et al. (2017). Effect of integrated community neurology on utilization, diagnostic testing, and | Wrong population |
|  | access. *Neurology. Clinical practice*.<br>https://dx.doi.org/10.1212/CPJ.0000000000000378 |  |
| 15. | Elston J, Grading F, Asthana S, Fox M, Dawson L, Butler D, and Byng, R. (2019). Impact of "enhanced" intermediate care located in a health & wellbeing hub at the integrated care organisation (ico) in torbay and south devon, uk. *International Journal of Integrated Care (IJIC)* http://doi.org/10.5334/ijic.s3584 | Conference abstract |
| 16. | Exley J, Abel G A, Fernandez Jose-Luis, Pitchforth E, Mendonca S, Yang M, Roland M, and McGuire, A. (2019). Impact of the Southwark and Lambeth Integrated Care Older People's Programme on hospital utilisation and costs: controlled time series and cost-consequence analysis. *BMJ open*. https://dx.doi.org/10.1136/bmjopen-2018-024220 | Wrong outcome |
| 17. | Farrell E, McCaffrey P, Toner R, and Sheeran, C. (2017). Evaluating the impact of an Acute Care at Home service on acute hospital admissions. *International Journal of Integrated Care (IJIC)*. https://doi.org/10.5334/ijic.3771 | Conference abstract |
| 18. | Gerlach L B, Mavandadi S, Maust D T, Streim J E and Oslin D W. (2018). Improving Access to Collaborative Behavioral Health Care for Rural-Dwelling Older Adults. *Psychiatric services (Washington, D.C.)* https://dx.doi.org/10.1176/appi.ps.201700026 | Wrong outcome |
| 19. | Giguere A M C, Farmanova E, Holroyd-Leduc J M, Straus S E, Urquhart R, and Carnovale V et al. (2018). Key stakeholders' views on the quality of care and services available to frail seniors in Canada. *BMC geriatrics*, https://dx.doi.org/10.1186/s12877-018-0969-y | Wrong intervention |
| 20. | Guan L, Wang C, Zhao B, Yang M, Zhu S, and Wu, X. (2022). Evaluation of Whether Emergency Physicians Should Join the Multidisciplinary Team for Older Hip Fracture Patients. *Frontiers in surgery*. https://dx.doi.org/10.3389/fsurg.2022.842978 | Not OECD |
| 21. | Guilcher S J T, Everall A, Wodchis W, DeGraaf-Dunlop J, Bar-Ziv S, Embuldeniya G, and Kuluski K. (2019). Understanding transitions of care in older adults with hip fractures: A qualitative multiple-case study in Ontario. *International Journal of Integrated Care (IJIC)*. https://doi.org/10.5334/ijic.s3283 | Conference abstract |
| 22. | Guy J, Benna M, Xia Y, Dagenet E, Ben Mrad M and Jmour O et al. (2019). Quality insurance in head and neck cancer multidisciplinary team meetings: A watchful eye on real-life experience. *Oral oncology*.<br>https://dx.doi.org/10.1016/j.oraloncology.2019.02.020 | Wrong design |
| 23. | Hall J, Dorrian C, Bradley S, Allan W, O'Keeffe R, and McGibbon F. (2016). Technology Enabled Care (TEC) provision for the care home sector in the Scottish Highlands: video conferencing in care homes. *International Journal of Integrated Care (IJIC)*. https://doi.org/10.5334/ijic.2898 | Conference abstract |
| 24. | Henderson J, Dawson S, Fuller J, O'Kane D, Gerace A, Oster C, and Cochrane, E M. (2018). Regional responses to the challenge of delivering integrated care to older people with mental health problems in rural Australia. *Aging & Mental Health*.<br>https://doi.org/10.1080/13607863.2017.1320702 | Wrong outcome |
| 25. | Hendry A, Vanhecke E, Carriazo A M, Lopez-Samaniego L, Espinosa J M, Sezgin D. (2019). Integrated Care Models for Managing and Preventing Frailty: A Systematic Review for the European Joint Action on Frailty Prevention (ADVANTAGE JA). *Translational medicine @ UniSa* | Wrong Study Design – SR to unpick |
| 26. | Heyzer L, Ramason R, De Castro M J A, Lim Chan W W L, and Loong C Y, Kwek E B K. (2022). Integrated hip fracture care pathway (IHFCP): reducing complications and improving outcomes. *Singapore medical journal*.<br>https://dx.doi.org/10.11622/smedj.2021041 | Not OECD |
| 27. | Hiltunen A M, Horhammer I, Silander K, Kaikuluoma J and Linna M. (2020). Integrating health service delivery for geriatric patients after hospital admission-A register study on the outcomes and costs. *Health Services Management Research*<br>https://dx.doi.org/10.1177/0951484819887668 | Wrong outcome |
| 28. | Janse B, Huijsman R, Looman W M and Fabbricotti I N. (2018). Formal and informal care for community-dwelling frail elderly people over time: A comparison of integrated and usual care in the Netherlands. *Health & social care in the community*.<br>https://dx.doi.org/10.1111/hsc.12516 | Wrong outcome |
| 29. | Koskas P, Pons-Peyneau C, Romdhani M, Houenou-Quenum N, Tigue-Wato A, Galleron S, and Drunat O. (2018). Effectiveness of multidisciplinary consultation for older adults with Alzheimer's disease in response to acute situations. *L'Encephale*.<br>https://dx.doi.org/10.1016/j.encep.2018.01.008 | Wrong outcome |
| 30. | Kukhareva P V, Li H, Caverly T J, Fagerlin A, Del F G, Hess R et al. (2024). Lung Cancer Screening Before and After a Multifaceted Electronic Health Record | Wrong outcome |
|  | Intervention: a Nonrandomized Controlled Trial. JAMA network open. https://doi.org/10.1001/jamanetworkopen.2024.15383 |  |
| 31. | Kuluski, Kerry and Gill, Ashlinder and McKillop, Ann and Parsons, John and Peckham, Allie and Sheridan, Nicolette and Upshur, Ross E. G. and Wong-Cornall, Cecilia (2017). The Unmet Needs of Patients and Carers within Community Based Primary Health Care. *International Journal of Integrated Care (IJIC)* https://doi.org/10.5334/ijic.3251 | Conference abstract |
| 32. | Kurpas D, Gwyther H, Szwamel K, Shaw R L, D'Avanzo B, and Holland C A, Bujnowska-Fedak M M. (2018). Patient-centred access to health care: a framework analysis of the care interface for frail older adults. *BMC geriatrics* https://dx.doi.org/10.1186/s12877-018-0960-7 | Wrong intervention |
| 33. | Kwek J L, Griva K, Kaur N, Chong K Y, Chua Z Y, and Sim G H A et al. (2022). Effectiveness and acceptability of a multidisciplinary approach in improving the care of patients with advanced chronic kidney disease: a pilot study. *International urology and nephrology*. https://doi.org/10.1007/s11255-021-02946-z | Not OECD |
| 34. | Leahy A, Barry L, Corey G, Whiston A, Purtill H and Shchetkovskyy D et al. (2023). The impact of frailty screening of older adults with multidisciplinary assessment of those at risk during emergency hospital attendance on the quality and safety of care (Solar): a randomised controlled trial. *Emergency medicine journal*. https://doi.org/10.1136/emmermed-2023-IAEM.6 | Conference abstract |
| 35. | Lee L, Hillier L M, McKinnon Wilson J, Gregg S, Fathi K, Sturdy Smith C and Smith M. (2018). Effect of Primary Care Based Memory Clinics on Referrals to and Wait-Time for Specialized Geriatric Services. https://doi.org/10.1111/jgs.15169 | Wrong study design |
| 36. | Liang W, Qin G, Yu L, and Wang Y. (2023). Reducing complications of femoral neck fracture management: a retrospective study on the application of multidisciplinary team. *BMC musculoskeletal disorders*. https://dx.doi.org/10.1186/s12891-023-06455-1 | Not OECD |
| 37. | Lynch G, Tower M and Venturato L. (2015). Identifying outcomes associated with co-managed care models for patients who have sustained a hip fracture: an integrative literature review. *International journal of orthopaedic and trauma nursing*. https://dx.doi.org/10.1016/j.ijotn.2014.07.002 | Wrong study design |
| 38. | Mann J, Quigley R, Harvey D, Tait M, Williams G, and Strivens E. (2020). OPEN ARCH: integrated care at the primary secondary interface for the community-dwelling older person with complex needs. *Australian Journal of Primary Health*. https://doi.org/10.1071/py19184 | Wrong study design |
| 39. | Mann J, Thompson F, McDermott R, Esterman A and Strivens E. (2021). Impact of an integrated community-based model of care for older people with complex conditions on hospital emergency presentations and admissions: a step-wedged cluster randomized trial. *BMC health services research*. https://dx.doi.org/10.1186/s12913-021-06668-x | Wrong outcome |
| 40. | Mas M A, Inzitari M, Sabate S, Santauegenia S J, and Miralles R. (2017). Hospital-at-home Integrated Care Programme for the management of disabling health crises in older patients: comparison with bed-based Intermediate Care. *Age and ageing*. https://dx.doi.org/10.1093/ageing/afx099 | Wrong outcome |
| 41. | Mas M A, Miralles R, Ulldemolins M J, Garcia R, Gracia S, and Picaza J M et al. (2023). Evaluating Person-Centred Integrated Care to People with Complex Chronic Conditions: Early Implementation Results of the ProPCC Programme. *International journal of integrated care*. https://dx.doi.org/10.5334/ijic.7585 | Wrong outcome |
| 42. | McHugh S, Wall O, Cronin F, Hosford O, Murphy R, and Timmons S et al. (2017). Implementing new ways of working to reduce the risk of falls among older people: an evaluation of community-based falls risk assessment clinics. *International Journal of Integrated Care (IJIC)*. https://doi.org/10.5334/ijic.3720 | Conference abstract |
| 43. | Mealy B, Nevin A, Lavan A, Jariol A L, Roll V and Connor S. et al. (2022). Home on time: multidisciplinary intervention reduces length of stay and delays in care transfers on an acute geriatric medicine ward. *Age and ageing*. https://doi.org/10.1093/ageing/afac218 | Conference abstract |
| 44. | Moran A, Nancarrow S A, and Enderby P. (2015). Mechanisms to enhance the effectiveness of allied health and social care assistants in community-based rehabilitation services: a qualitative study. *Health & Social Care in the Community*. https://doi.org/10.1111/hsc.12158 | Wrong intervention |
| 45. | Mueller-Stierlin A S, Helmbrecht M J, Herder K, Prinz S, Rosenfeld N, Walendzik J, et al. (2017). Does one size really fit all? The effectiveness of a non-diagnosis-specific integrated mental health care program in Germany in a prospective, parallel-group | Wrong outcome |
|  | controlled multi-centre trial. *BMC psychiatry*. https://doi.org/10.1186/s12888-017-1441-9 |  |
| 46 | Murtagh F E M, Okoeki M, Ukoha-Kalu B O, Khamis A, Clark J, and Boland J W et al. (2023). A non-randomised controlled study to assess the effectiveness of a new proactive multidisciplinary care intervention for older people living with frailty. *BMC geriatrics*. https://dx.doi.org/10.1186/s12877-023-03727-2 | Wrong outcome |
| 47 | Nash P, Horsnell H, O'Connor I, Burke P, Devlin V, Fitzpatrick S, and Rothwell G. (2017). National Patient Flow Improvement Programme. *International Journal of Integrated Care (IJIC)*. https://doi.org/10.5334/ijic.3498 | Conference abstract |
| 48 | Nct. (2020). SOLAR- Frailty Screening and Multidisciplinary Assessment of Older Adults in the Emergency Department https://clinicaltrials.gov/show/NCT04629690 | Duplicate |
| 49 | Nossa J M, Escobar N, Marquez D, Leal E, Cabal F, and Barreto A. (2016). Applying a multidisciplinary program for managing hip fractures in the elderly: incidence of comorbidities and their impact on surgical opportunity. *Revista Colombiana de Ortopedia y Traumatologia*. https://dx.doi.org/10.1016/j.rccot.2016.10.005 | Not in English |
| 50 | O'Caoimh R, Costello M, Judge C, Morrison L, Ghafa, Z, Ali B, and Mannion E et al. (2017). Integrating Care and Improving Flow for Frail Older Adults through the Development of an Acute Inpatient Frailty Service. *International Journal of Integrated Care (IJIC)*. https://doi.org/10.5334/ijic.3724 | Conference abstract |
| 51 | O'Connor K A, McHugh S, Dukelow T, Wall O, Murphy R, O'Keeffe A, Cronin F, Barry P, and Moriarty E. (2017). Community teams, specialist fall services and emergency department implementing an integrated regional falls risk assessment and prevention pathway. *International Journal of Integrated Care (IJIC)*. https://doi.org.uk10.5334/ijic.3686 | Conference abstract |
| 52 | Oreilly O, Courtney G, and Croke E. (2017). Integrated Care for Emergency Medical Patients - Irish Acute Medicine Programme. *International Journal of Integrated Care (IJIC)*. https://doi.org/10.5334/ijic.3166 | Conference abstract |
| 53 | Roe L, Normand C, Wren M, Browne J and O'Halloran, A M. (2017). The impact of frailty on healthcare utilisation in Ireland: evidence from the Irish longitudinal study on ageing. *BMC geriatrics*. https://dx.doi.org/10.1186/s12877-017-0579-0 | Wrong outcome |
| 54 | Roe L D, Browne J, Wren M, O'Halloran A and Normand C. (2017). Frail older people's use and experience of the Irish healthcare system: a mixed methods study to inform the design of integrated models of care. *International Journal of Integrated Care (IJIC)*. https://doi.org/10.5334/ijic.3171 | Conference abstract |
| 55 | Sampson M, Bailey M, Clark J, Evans M L, Fong R, and Hall, H. et al. (2017). A new integrated care pathway for ambulance attended severe hypoglycaemia in the East of England: The Eastern Academic Health Science Network (EAHSN) model. *Diabetes research and clinical practice*. https://dx.doi.org/10.1016/j.diabres.2017.08.017 | Wrong outcome |
| 56 | Schubert C C, Penney L S, Schwartzkopf A L, Damush T M, Preddie A, and Flemming, S. (2024). Expanding Access to Comprehensive Geriatric Evaluation via Telehealth: Development of Hybrid-Virtual Home Visits. *Journal of general internal medicine*. https://dx.doi.org/10.1007/s11606-023-08460-5 | Wrong outcome |
| 57 | Seamer P, Lloyd T, Conti S and O'Neill S. (2023). The Long-Term Impacts of an Integrated Care Programme on Hospital Utilisation among Older Adults in the South of England: A Synthetic Control Study. *International journal of integrated care*. https://dx.doi.org/10.5334/ijic.6475 | Wrong outcome |
| 58 | Shigemoto K, Sawaguchi T, Horii T, Goshima K, Iwai S, Higashikawa T, and Okuro, M. (2022). Multidisciplinary care model for geriatric patients with hip fracture in Japan: 5-year experience. *Archives of orthopaedic and trauma surgery* https://dx.doi.org/10.1007/s00402-021-03933-w | Wrong design |
| 59 | Shortt J and Ryan A. (2017). Bringing Cardiac Diagnostics to the Heart of the Community. *International Journal of Integrated Care (IJIC)*. https://doi.org/10.5334/ijic.3810 | Conference abstract |
| 60 | Sourial N, Hacker T M, Arsenault-Lapierre G, Mehta K, Kay K and Vedel, I. (2022). Interprofessional primary care: indispensable for family physicians yet invisible to older patients. *Journal of Interprofessional Care*. https://doi.org/10.1080/13561820.2022.2037529 | Wrong outcome |
| 61 | Swart E, Vasudeva E, Makhni E, Macaulay W, Bozic K, Makhni E C, and Bozic K J. (2016). Dedicated Perioperative Hip Fracture Comanagement Programs are Cost-effective in High-volume Centers: An Economic Analysis. *Clinical Orthopaedics & Related Research*. https://doi.org/10.1007/s11999-015-4494-4 | Wrong study design |
| 62 | Wu X, Tian M, Zhang J, Yang M, Gong X, and Liu Y et al. (2019). The effect of a multidisciplinary co-management program for the older hip fracture patients in Beijing: a "pre- and post-" retrospective study. *Archives of Osteoporosis* https://dx.doi.org/10.1007/s11657-019-0594-1 | Not OECD |
| 63 | Wu X N, Zhang S Y, Peng C, and Shen X. (2019). Clinical significance of multidisciplinary team collaboration for the treatment of hip fractures in the elderly. *Journal of the American Geriatrics Society*. http://doi.org/10.1111/jgs.16120 | Conference abstract |
| **Clinical trial registers** |  |  |
| 1. | CAMH - McMaster Collaborative Care Initiative for Mental Health Risk Factors in Dementia: Depression, Anxiety, and Mild Cognitive Impairment. (2016). https://clinicaltrials.gov/study/NCT02955719 | Duplicate: retrieved in database search: Dham 2022 |
| 2. | A Qualitative Study to Investigate Patients', Carers' and Providers' Perspectives on Integrated Care and Integrated Care Pathways for Frail Older People in Lambeth and Southwark, South London (2017). https://clinicaltrials.gov/study/NCT03260933 | Not found: no protocol or final report identified |
| 3. | Integrated Care for Frail Older People. (2017). https://clinicaltrials.gov/show/NCT03260933 | Same report as above – no final report identified |
| 4. | SmartCare Project: Joining up ICT and Service Processes for Quality Integrated Care in Europe. (2017). https://clinicaltrials.gov/study/NCT03112109 | Wrong outcome |
| 5. | SMARTCARE Project: Deploying Integrated Health & Social Services for Independent Living by Older People. (2017). https://clinicaltrials.gov/show/NCT03112109<br>https://pilotsmartcare.eu/outcomes/deliverables/<br>Dyrvig A. et al. (2015). D8.3 Second interim process evaluation report. https://pilotsmartcare.eu/outcomes/deliverables/<br>Stafylas, P. et al. (2016). D8.4 SMARTCARE project outcomes. https://pilotsmartcare.eu/outcomes/deliverables/ | As above – wrong outcome |
| 6. | BeyondSilos - Integrated Care Leading to a More Personalised and Coordinated Care, Improving Outcomes for Elderly Patients, Delivering More Effective Care and Support, and Provide More Cost-Efficient Health and Social Services. (2017). https://clinicaltrials.gov/study/NCT03111004<br>https://doi.org/10.2196/20938<br>Piera-Jimenez J, Daugbjerg S. et al (2020). BeyondSilos, a Telehealth-Enhanced Integrated Care Model in the Domiciliary Setting for Older Patients: Observational Prospective Cohort Study for Effectiveness and Cost-Effectiveness Assessments. *JMIR Medical Informatics*. https://doi.org/10.2196/20938 | Wrong outcome |
| 7. | Integrated Care With GP Participation for Older Persons in the Ambulatory Care Hub: a Prospective Cohort Study of Clinical and Process Outcomes. (2022). https://clinicaltrials.gov/study/NCT05527223 | Not found: no protocol or final report identified |
| 8. | Nurse-led Integrated Care of Complex Patients Facilitated by Telemonitoring: The Safe, Managed, and Responsive Transitions (SMaRT) Study. (2022). https://clinicaltrials.gov/study/NCT05543720 | Still collecting data |
| 9. | Protocol for Inpatient Nursing Frailty Assessment (INFA): Comprehensive Geriatric Assessment and Multidisciplinary Intervention for Frail Hospitalised Older Adults. (2024). https://clinicaltrials.gov/study/NCT06469723 | Still collecting data |
| 10. | The Role of the Geriatric Team in Facilitating the Emergency Department (ED) Workflow for Elderly Patients: Effects on Rate of Hospitalization and on Community Health Management: A Multicentric Randomized Controlled Intervention Study. (2024). https://clinicaltrials.gov/study/NCT06788210 | Not found: no protocol or final report identified |
| 11. | INSPIRE: Feasibility of a Community-based Integrated Care Model for Older Adults Living at Home. (2022). https://clinicaltrials.gov/study/NCT05302310<br>https://doi.org/10.1186/s12877-022-03552-z<br>Yip O, Dhaini S et al. (2022) Health and social care of home-dwelling frail older adults in Switzerland: a mixed methods study. *BMC Geriatrics*. https://doi.org/10.1186/s12877-022-03552-z | Wrong outcome |
| 12. | Transforming Primary Care for Older Canadians Living with Frailty. (2017). https://clinicaltrials.gov/show/NCT03442426 | Two published protocols but no final report |

| Grey literature searches |  |  |
| --- | --- | --- |
| 1. | Bevan Commission. (2016). Improving health and healthcare: Barriers and enablers for change. Swansea: Bevan Commission.<br>https://bevancommission.org/improving-health-and-healthcare-barriers-and-enablers-of-change/ | No explicit methods |
| 2. | Bevan Commission. (2025). Why wait? Building on proven initiatives to reduce waits in Wales. Swansea: Bevan Commission.<br>https://bevancommission.org/why-wait/ | No explicit methods |
| 3. | Gentry T, Jopling K, and Reeves C. (2023). Fixing the foundations: Why it's time to rethink how we support older people with health problems to stay well at home. London: Age UK.<br>https://www.ageuk.org.uk/siteassets/documents/reports-and-publications/reports-and-briefings/health--wellbeing/fixing-the-foundations/FTF-feb-2023.pdf | No explicit methods |
| 4. | Health Education England. (2016). Care Navigation: A Competency Framework. London: Health Education England.<br>https://www.hee.nhs.uk/sites/default/files/documents/Care%20Navigation%20Competency%20Framework_Final.pdf | No explicit methods |
| 5. | Hopper, A. (2021). Getting it Right First Time. Geriatric Medicine GIRFT Programme National Specialty Report.<br>https://gettingitrightfirsttime.co.uk/wp-content/uploads/2021/09/Geriatric-Medicine-Sept21h.pdf | No explicit methods |
| 6. | NHS England. (2016). New Care Models: Vanguard - developing a blueprint for the future of NHS and care services.<br>https://www.england.nhs.uk/wp-content/uploads/2015/11/new_care_models.pdf | No explicit methods |
| 7. | NHS RightCare. (2019). Frailty Toolkit: Optimising a frailty system. London: NHS England. https://www.england.nhs.uk/rightcare/wp-content/uploads/sites/40/2019/07/frailty-toolkit-june-2019-v1.pdf | No explicit methods |
| 8. | Royal College of General Practitioners. (2016). Integrated care for older people with frailty Innovative approaches in practice.<br>https://www.bgs.org.uk/sites/default/files/content/resources/files/2018-10-09/RCGP-Integrated-care-for-older-people-with-frailty-2016.pdf | No explicit methods |
| 9. | Age Cymru. (2024). Why are we still waiting? Delays in social care in Wales.<br>https://www.agecymru.wales/siteassets/documents/why-are-we-still-waiting/why-are-we-still-waiting.pdf | Wrong design and wrong intervention |
| 10. | Bliss A, Williamson S, and Alayo, L. (2024). The state of integrated care systems 2023/24: tackling today while building for tomorrow. London: NHS Confederation.<br>https://www.nhsconfed.org/publications/state-integrated-care-systems-202324 | Wrong design |
| 11. | Cairney P, Boswell J, Bliss A, Mahmood H, and Raine, J. (2024). Unlocking prevention in integrated care systems. London: NHS Confederation.<br>https://www.nhsconfed.org/publications/unlocking-prevention-integrated-care-systems | Wrong outcome |
| 12. | Carnes Chichlowska S, Burholt V, Beech C, and Dobbs C. (2015). The Integrated Care Evaluation Framework. A Realistic Evaluation of integrated health and social care services in Wales. Welsh Government.<br>https://www.gov.wales/sites/default/files/statistics-and-research/2019-07/150514-integrated-care-evaluation-framework-en.pdf | Wrong outcome |
| 13. | NHS Confederation. (2022). The state of integrated care systems 2021/22. London: NHS Confederation.<br>https://www.nhsconfed.org/publications/state-integrated-care-systems-2021-22 | Wrong outcome |
| 14. | NHS Confederation. (2023). The state of integrated care systems 2022/23: Riding the storm. London: NHS Confederation.<br>https://www.nhsconfed.org/publications/state-integrated-care-systems-2022-23 | Wrong outcome |
| 15. | NHS Confederation. (2023). The evolution of provider collaboration. London: NHS Confederation.<br>https://www.nhsconfed.org/publications/evolution-provider-collaboration | Wrong design and intervention |
| 16. | Older People's Commissioner for Wales. (2017). GP Services in Wales: The Perspective of Older People: older people's experiences of accessing and using GP services in Wales.<br>https://olderpeople.wales/wp-content/uploads/2022/03/GP-Services-in-Wales-The-Perspective-of-Older-People.pdf | Wrong design and intervention |
| 17. | Robertson R, Blythe N, and Jefferies D. (2023). Tackling health inequalities on NHS waiting lists: Learning from local case studies. London: The King's Fund.<br>https://www.kingsfund.org.uk/insight-and-analysis/reports/health-inequalities-nhs-waiting-lists | Wrong intervention? |
| 18. | Alderwick H, Robertson R, Appleby J et al. (2015). Better value in the NHS The role of changes in clinical practice. Kings Fund. | No explicit methods |
|  | https://www.kingsfund.org.uk/insight-and-analysis/reports/better-value-nhs |  |
| 19 | Baird B, Reeve H, Ross S et al. (2018). Innovative models of general practice. Kings Fund. https://www.kingsfund.org.uk/insight-and-analysis/reports/innovative-models-general-practice | Wrong study design |
| 20 | Naylor C, Alderwick H, and Honeyman M. (2015). Acute hospitals and integrated care: From hospitals to health systems. London: The King's Fund. https://www.kingsfund.org.uk/publications/acute-hospitals-integrated-care | No explicit methods |
| 21 | Jackson P, McCaffrey R, Swindlehurst M and Tomkinson A. (2018). Foundations for Integrated Care: A Position Paper for the pilot in the West. https://www.gov.im/media/1366687/foundations-for-integrated-care-24th-dec-18.pdf | Wrong intervention |
| 22 | Page B, Sugavanam T, Fitzpatrick R et al. (2024). Floundering or Flourishing? Early Insights from the Inception of Integrated Care Systems in England. *International Journal of Integrated Care*. https://doi.org/10.5334/ijic.7738 | Wrong Intervention |
| 23 | Czypionka T, Kraus M, Reiss M, et al. (2020). The patient at the centre: Evidence from 17 European integrated care programmes for persons with complex needs. *BMC Health Services Research*, 20, Article 1102. https://doi.org/10.1186/s12913-020-05917-9 | Wrong outcome |
| 24 | Calafiore M, Chazard E, Averlant L, et al (2021). Wide variations among GPs in the level of interest in integrated care for frail, elderly adults and their association with data on consulting and prescribing: an analysis of the PAERPA project. *British Journal of General Practice*. https://doi.org/10.3399/BJGP.2021.0626 | Wrong outcome |
| 25 | Transforming Cancer Services Team. (2019). A guide to reducing variation and improving outcomes in cancer rehabilitation in London. London: NHS England. https://transformationpartners.nhs.uk/wp-content/uploads/2019/07/A-guide-to-reducing-variation-and-improving-outcomes-in-cancer-rehabilitation-in-London.pdf | Wrong outcome |
| **Studies identified from systematic/scoping reviews (unpicking)** |  |  |
| 1. | De Lange J, Deusing E, Francke A L et al. (2018). Factors facilitating dementia case management: Results of online focus groups. *Dementia*. https://doi.org/10.1177/1471301216634959 | Wrong outcome |
| 2. | Lind K B, Soerensen C A, Salamon S A et al. (2015). Impact of clinical pharmacist intervention on length of stay in an acute admission unit: a cluster randomised study. *European Journal of Hospital Pharmacy*. https://doi.org/10.1136/ejhpharm-2015-000767 | Wrong intervention |
| 3. | Quigley R, Russell S, Harvey D and Mann J. (2021). OPEN ARCH integrated care model: experiences of older Australians and their carers *Australian Journal of Primary Health* https://doi.org/10.1071/PY20203 | Wrong outcome |

## Footnotes

1 Hazard ratio can be defined as a measure of how often the outcome event happens (early treatment initiation) in the intervention group compared to how often it happens in the control group, over time National Cancer Institute. (2025). NCI Dictionaries - Hazard Ratio. USA: National Cancer Institute. Available at: https://www.cancer.gov/publications/dictionaries/cancer-terms/def/hazard-ratio [Accessed 21/03/2025].

2 Grading was based on guidance by Public Health Agency of Canada (2014). For more information see section 5.8.

4 The preliminary literature review is available on request.

* *This section has been completed by the Centre for Health Economics & Medicines Evaluation (CHEME), Bangor University*

## Notes

### Competing Interest Statement

The authors have declared no competing interest.

